# The gut-microbiome in adult Attention-deficit/hyperactivity disorder - A Meta-analysis

**DOI:** 10.1101/2023.12.18.23300126

**Authors:** Babette Jakobi, Priscilla Vlaming, Danique Mulder, Marta Ribases, Vanesa Richarte, Josep Antoni Ramos-Quiroga, Indira Tendolkar, Philip van Eijndhoven, Janna N. Vrijsen, Jan Buitelaar, Barbara Franke, Martine Hoogman, Mirjam Bloemendaal, Alejandro Arias-Vasquez

## Abstract

Attention-deficit/hyperactivity disorder (ADHD) is a common neurodevelopmental condition that persists into adulthood in the majority of individuals. While the gut-microbiome seems to be relevant for ADHD, the few publications on gut-microbial alterations in ADHD are inconsistent, in the investigated phenotypes, sequencing method/region, preprocessing, statistical approaches, and findings. To identify gut-microbiome alterations in adult ADHD, robust across studies and statistical approaches, we harmonized bioinformatic pipelines and analyses of raw 16S rRNA sequencing data from four adult ADHD case-control studies (N_ADHD_=312, N_NoADHD_=305). We investigated diversity and differential abundance of selected genera (logistic regression and ANOVA-like Differential Expression tool), corrected for age and sex, and meta-analyzed the study results. Converging results were investigated for association with hyperactive/impulsive and inattentive symptoms across all participants. Beta diversity was associated with ADHD diagnosis but showed significant heterogeneity between cohorts, despite harmonized analyses. Several genera were robustly associated with adult ADHD; e.g., *Ruminococcus_torques_group* (LogOdds=0.17, p_fdr_=4.42×10^−2^), which was more abundant in adults with ADHD, and *Eubacterium_xylanophilum_group* (LogOdds= −0.12, *p*_fdr_=6.9 x 10^−3^), which was less abundant in ADHD. *Ruminococcus_torques_group* was further associated with hyperactivity/impulsivity symptoms and *Eisenbergiella* with inattention and hyperactivity/impulsivity (p_fdr_<0.05). The literature points towards a role of these genera in inflammatory processes. Irreproducible results in the field of gut-microbiota research, due to between study heterogeneity and small sample sizes, stress the need for meta-analytic approaches and large sample sizes. While we robustly identified genera associated with adult ADHD, that might overall be considered beneficial or risk-conferring, functional studies are needed to shed light on these properties.

## Introduction

Attention-deficit/hyperactivity disorder (ADHD) is a common neurodevelopmental condition ^1^, characterized by symptoms of inattention and hyperactivity/impulsivity (American Psychiatric Association 2013). The clinical presentation of ADHD is quite heterogeneous; symptoms and impairments persist into adulthood in the majority of affected individuals, and other psychiatric and somatic problems often accompany ADHD ^2^. The etiology of ADHD is likely multifactorial, combining genetic and environmental risk-factors or protective influences ^3^. Studies have suggested potential roles of e.g. immune- and inflammatory processes and alterations in dopaminergic and serotonergic neurotransmission, resulting in altered brain development and functioning, for the emergence of ADHD symptoms ^4^.

The gut-microbiome is involved in early brain-development as well as every-day brain functioning; it can modulate the bioavailability of key-signaling molecules (e.g. neurotransmitters or nutrients relevant for energy homeostasis) by influencing the metabolism and the integrity of intestinal- and-blood-brain barriers ^5^. Through the regulation of the intestinal barrier, but also through the production of short-chain-fatty-acids (SCFA) and the release of cytokines, it further plays an important role in immune and inflammatory responses (for a review see ^5^). These pathways might influence ADHD symptoms and pathophysiology ^6, 7^. Studies showing associations of microbial diversity and composition with altered neurodevelopment (for a review, see ^8^), psychiatric disorders (for a review, see ^9^) and common metabolic comorbidities of ADHD ^10, 11^ have fueled hypotheses about the potential role of the gut-microbiome alterations for ADHD. The few published studies investigating gut-microbiome alterations in ADHD, however, report inconsistent results. Most authors reported no differences between individuals with and without ADHD or conflicting results for gut-microbiome diversity (^12–16^). Recent systematic reviews showed that differential abundance of some taxa between individuals with and without ADHD was reported in all published studies ^13^. However, the results converged at the genus-level (marking the highest resolution of taxonomic assignment from 16S sequencing) in maximally two out of eight published studies in children and adolescents (see Supplementary Table 1 ^17, 18^). The three studies published on adults so far showed no overlap in results, despite a substantial overlap in samples and wet-lab procedures between Aarts et al. (2017) and Szopinska-Tokov et al. (2020) ^19–21^. The scarcity of consistent results renders biological interpretations of gut-microbiome alterations as well as consideration of the abundance of particular taxa as potential biomarkers or treatment targets for ADHD difficult.

Inconsistencies in reported results might be explained by a variety of factors; ^22^ provide an overview on this topic. Most relevant may be the expected small effect sizes of microbiome alterations due to high inter- and intrapersonal variability, in conjunction with the statistical testing of a high number of features and the small sample sizes of gut-microbiome studies published to date (mean n_ADHD_ = 39, mean n_noADHD_ = 49 ^13^); those might lead to high false positive rates and at the same time low detection of true effects. Heterogeneity in terms of age (children, adolescents, adults), sex (some studies only focused on males), and ethnic origin (Asian or European) might render summarizing and comparing the results more difficult. Accounting for common confounders (e.g. age, sex, diet), focusing on one developmental stage, and meta-analysis across studies might help increase the robustness of findings. Importantly, methodological choices can strongly influence observed microbial diversity and composition, e.g. in the technical variation of wet-lab procedures and processing of microbiome data. While most studies use 16S rRNA sequencing, different sequencing methods (extraction, storing, platforms, protocols) and the choice of the regions of the 16S gene can have a substantial influence on the identified features (for a review, see ^23^). The lack of consensus on preprocessing pipelines and statistical analysis tools or approaches further contributes to the scattered and irreproducible gut-microbiome associations with ADHD across the field. 16S microbiome data comes with particular properties, that should be accounted for: 1) Compositionality-bias (stemming from the fact that the count of a feature does not carry information about its absolute abundance) results from the sequencing and the restriction to the library size. Transformations such as the center-log-ratio (CLR) scale the counts to a reference and can successfully eliminate compositionality bias of between-sample comparisons ^24^. 2) Zero-inflation and un-identifiable sources of zeros (e.g. sampling bias, sequencing bias, true zero) are particularly problematic for differential abundance analysis. Bias-related zeros increase the number of (uninformative) tests, and the resulting distribution has to be accounted for by the statistical approach. A recent paper ^25^ reviewed approaches to deal with these issues, showing that exclusion of features that are observed in less than 10% of the samples from further analysis (prevalence threshold) increased cross-method comparability and reduced the statistical testing burden, false discovery rates, and zero inflation bias while maintaining sufficient information content for downstream statistical analysis. Attempts to account for these biases in the statistical approaches, however, vary profoundly between research groups, resulting in incomparable results across studies. The comparison of results across tools is recommended, where converging results are most likely to reflect true findings. Next to logistic regression in case-control studies, ANOVA-like Differential Expression (ALDEx2 ^26^) is a promising approach to analyze differential abundance; this method produced the most comparable results across statistical approaches and studies while preserving a low false discovery rate ^25^.

In this study, we aimed to investigate gut-microbiome alterations in adult ADHD and identify genera that might be risk-conferring for the condition. To assure robustness of the results, we applied four strategies: 1) We harmonized the bioinformatic pipelines for sequencing data of four case-control cohorts of adults with and without ADHD (N=617); 2) we focused on one developmental stage, i.e. adulthood; 3) we investigated diversity and differential abundance across tools and indices, correcting for common confounders per study; 4) we meta-analyzed the results across studies. Converging results across tools and studies were analyzed for associations with the clinical representations of ADHD such as hyperactive/impulsive and inattentive symptoms.

## Materials and Methods

### Cohort description

We requested data from all ADHD case-control studies that included 16S fecal gut-microbiome samples, identified in two recent systematic reviews ^13, 16^, see Supplementary Section 1.1, Table 1 and Supplementary Figure 15. We received clinical information and raw sequencing data from four adult cohorts (comprising the three articles including adults published to date: the NeuroIMAGE cohort ^20, 21^ and the Mental-Cat cohort from the Vall d’Hebron Research Institute in Barcelona (VHIR) ^19^; and unpublished data from the MIND-Set cohort ^27^ and our own cohort IMpACT2-NL ^28^.

**Table 1.** Demographic description and characteristics of the included studies.

| **Cohort**<br>Recruitment period<br>Recruitment Country | **IMpACT2-NL**<br>2017-2020<br>The Netherlands |  | **NeuroIMAGE**<br>2009- 2012<br>The Netherlands |  | **MIND-Set**<br>2016-2021<br>The Netherlands |  | **Mental-Cat**<br>2016-2018<br>Spain |  |
| --- | --- | --- | --- | --- | --- | --- | --- | --- |
| Diagnostic group | Control | ADHD | Control | ADHD | Control | ADHD | Control | ADHD |
| Number of subjects | 78 | 78 | 38 | 29 | 90 | 104 | 100 | 100 |
| Sex, proportion female participants | 0.51 | 0.57 | 0.47 | 0.31 | 0.54 | 0.42 | 0.53 | 0.49 |
| Age mean years (SD) | 34.49 (13.08) | 33.86 (10.31) | 21.84 (2.54) | 22.21 (3.05) | 38.55 (16.87) | 37.38 (12.55) | 30.3 (8.47) | 33.17 (11.41) |
| BMI mean (SD) | 24.85 (4.25) | 24.86 (4.49) | 22.84* (3.05) | 24.39 (3.79) | 23.85** (4.46) | 28.01*** (15.19) | 22.13 (2.91) | 24.67 (4.25) |
| Smoking, proportion non-smokers | 0.90 | 0.65 | NA | NA | 0.93 | 0.76 | NA | NA |
| ADHD medication proportion currently using ADHD medication | 0 | 0.51 | 0 | 0.42 | 0 | 0.84<sup>1</sup> | 0 | Exclusion criterium |
| Inattentive symptoms mean(SD) | 0.87 (1.26) | 5.65 (2.17) | 41.74 (10.67) | 58.45 (12.12) | 2.31* (1.98) | 8.86**** (2.81) | NA | 17.86 (4.33) |
| Hyperactive/impulsive symptoms mean(SD) | 0.79 (1.10) | 7.38 (1.83) | 45.42 (9.29) | 66.86 (11.32) | 2.51 (1.66) | 8.09 (2.27) | NA | 13.26 (6.08) |
| Instrument symptom assessment<sup>2</sup> | DIVA adult self-report |  | CAARS-DSM-IV adult self-report |  | CAARS-S.S short adult self-report |  | ADHD-RS adult self-report |  |
| Sequencing Region | V4 |  | V1/V2 |  | V4 |  | V3/V4 |  |
| Sequencing platform | Illumina NovaSeq 6000 |  | Illumina HiSeq PE300 |  | Illumina NovaSeq 6000 |  | Illumina MiSeq 300-nt |  |
| Antibiotic treatment, proportion using antibiotics | 0.01<sup>3</sup> |  | No information |  | 0 |  | Exclusion criterium |  |
| Psychiatric comorbidity, proportion of ADHD + additional current psychiatric diagnoses | Exclusion criterium<sup>4</sup> |  | Exclusion criterium<sup>5</sup> |  | 0.83 |  | Exclusion criterium |  |

Footnote: \* 1 missing value, \*\* 27 missing values, \*\*\*15 missing values, \*\*\*\*11 missing values.
<sup>1</sup> 65.5 % of MIND-Set participants with an ADHD diagnosis received additional pharmacological treatment targeting the central nervous system (mostly antidepressants), with a maximum of 6 different drugs prescribed to an individual.
<sup>2</sup> Symptom assessment tools are described in more detail in Supplementary Chapter 1.1.
<sup>3</sup> IMpACT2-NL assessed frequency of antibiotics use (frequent, sometimes, rarely or never; 96.4% of participants answered never or rarely, one control participant reported frequent antibiotics usage.
<sup>4</sup> Diagnoses of neurodevelopmental and current psychiatric disorders were excluded.
<sup>5</sup> Diagnoses of autism were excluded.

The NeuroIMAGE cohort comprises adolescents and young adults with ADHD, their family members as well as unrelated healthy controls; only adult participants were included in this study ^13, 29^. The Mental-Cat cohort consists of medication-naive adults with and without ADHD. The MIND-Set cohort includes adults with ADHD exhibiting a high level of psychiatric comorbidity and psychopharmacological treatment, as well as healthy individuals. For detailed information about individual studies, recruitment and inclusion/exclusion criteria, see Supplementary Chapter 1.1. Information about fecal sample collection, storing, and sequencing are provided in Supplementary Chapter 2. Our report follows STORMS guidelines for human microbiome research, whenever possible ^30^, see the Supplementary Table 14.

Participants with gut-related diseases (irritable bowel disease) and those with an unclear ADHD diagnosis were excluded, and overlapping samples between MIND-Set and IMpACT2-NL were removed from the MIND-Set cohort, resulting in a sample of 312 adults with and 305 adults without ADHD (56 exclusions, see Supplementary Figure 1). Table 1 provides a demographic overview of the included sample.

### Microbiome preprocessing

Preprocessing was harmonized and performed per study using QIIME2 pipeline defaults ^31^, Figure 1 summarizes the preprocessing steps. For all studies, the raw forward and reverse reads were demultiplexed and then denoised using DADA2 ^32^, where the primers were trimmed off, sequencing errors and erroneous read combinations were removed and clusters of representative sequences (amplicon sequence variants, ASVs) were identified. We assured high sequencing quality by truncating the reads displaying a signal drop below a median phred-score of 30 (marking 99.9% base-call accuracy ^33^) towards the end of the reads resulting in a truncation from basepair 260 in NeuroIMAGE and VHIR. For IMpACT and MIND-Set no truncation was applied (no signal drop). The sequencing data for NeuroIMAGE was delivered in four batches and merged after denoising. The ASVs were aligned to their phylogenic tree (fasttree2 ^34^). Taxonomy was assigned using a naive bayes classifier, pre-trained on a SILVA reference database of the full-length 16S gene (version 138, 99% OTUs full-length sequences, https://docs.qiime2.org/2022.2/data-resources/), to assure coherent classification irrespective of the sequencing region of the study. The resulting feature table with taxonomic assignments and the phylogenic tree were imported in R (version 4.2.1 ^35^). We removed non-bacterial ASVs and screened for read depth (plateau in the rarefaction curve) and summarized sequencing data after each step, see Supplementary Chapter 2. We then investigated potential differences in alpha and beta diversity as well as composition per study, correcting for age and sex. Other common confounders were assessed post-hoc (ADHD medication, diet) or not included due to (excess) missing information (body-mass-index (BMI), smoking, general medication, anti-/probiotics) or high co-linearity with ADHD-control grouping (smoking, psychopharmacological treatment, comorbidities).

**Figure 1.**
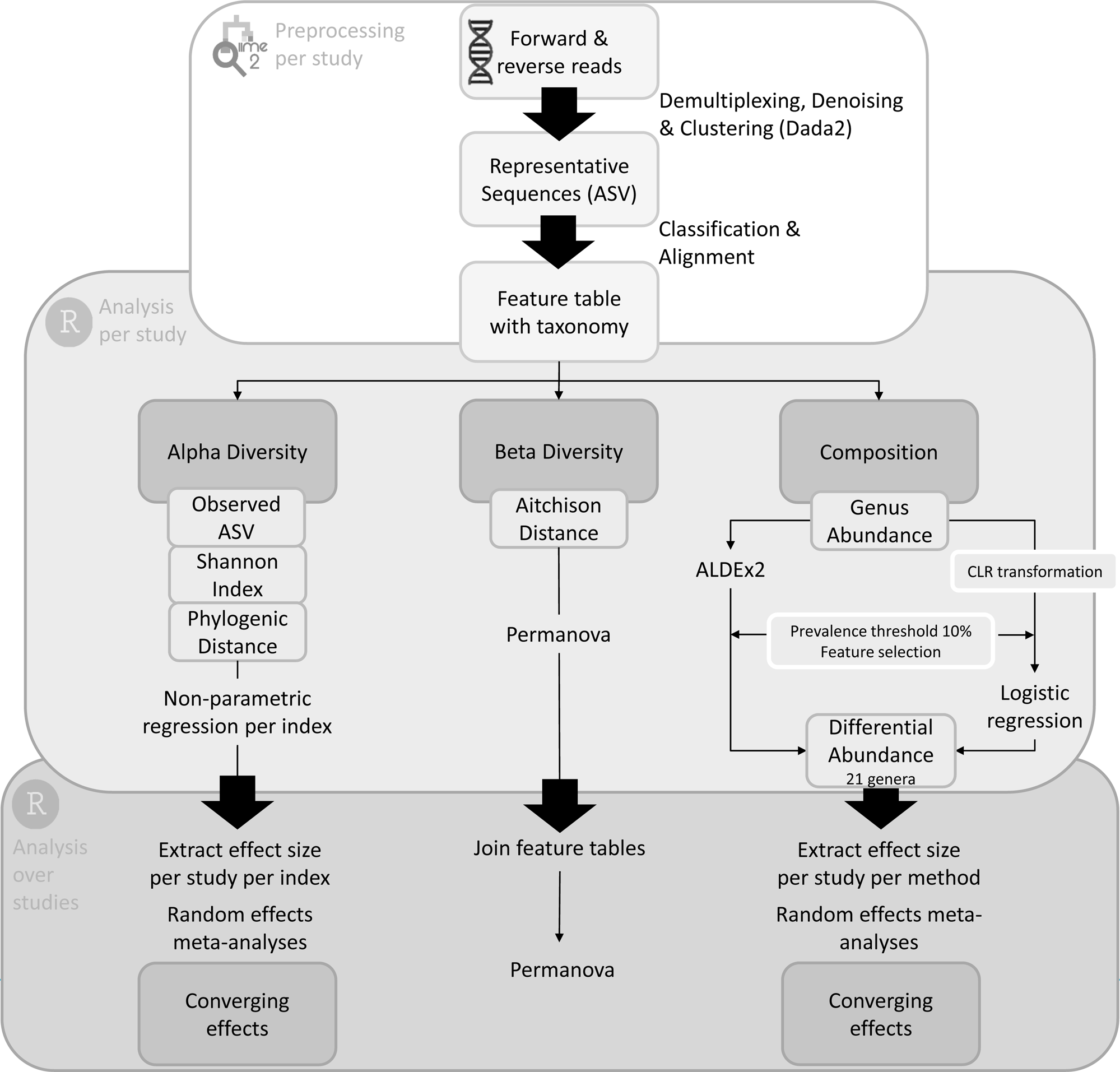
Workflow diagram of bioinformatic pipelines for preprocessing and statistical analyses. Section 1 (top) describes the preprocessing per study, section 2 (middle) describes the statistical analysis of diversity and composition per study and section 3 (bottom) describes the statistical analysis over all studies. Abbreviations: ASV: amplicon sequence variants, ALDEx2: ANOVA-like differential expression, CLR: center log ratio.

### Alpha Diversity Analysis

We estimated three indices of alpha diversity, considering 1) number of observed ASVs, 2) abundance of ASVs (Shannon index), and 3) phylogenic relationships between ASVs (phylogenic distance) (*microbiome* package,^36^). We applied rank-based nonparametric regression analysis (*Rfit,* ^37^) for each study (alpha diversity ∼ ADHD diagnosis). We further extracted the standardized correlation coefficient R as effect size measure per study and meta analyzed over the four cohorts with a random-effects models, which estimates study heterogeneity (*metafor* package ^38^).

### Beta Diversity Analysis

Beta diversity (the similarity of the microbiome between samples) was investigated per study, applying the Permanova (*adonis2)* with 999 permutations in order to estimate Aitchison distance by ADHD diagnosis (*vegan* package ^39^). We additionally investigated beta diversity differences over all studies in a mega-analytic fashion by combining all feature tables and including the cohort as a nominal dummy variable. We visualized the effect of ADHD diagnosis on beta diversity using Canonical Analysis of Principal coordinates (CAP) plots, supervised for group effects (*phyloseq* package ^40^).

### Composition

To investigate microbial composition, we applied a prevalence threshold of 10% (improves comparability over statistical tools, reduces the number of statistical tests and zero-biases ^25^) and aggregated the data to the genus level, see Supplementary Table 2 for counts per study. We applied CLR transformations to the count data to account for zero-inflation and compositionality biases ^24^.

#### Feature selection

To test only genera with potential informative value for the outcome of interest (ADHD diagnosis), we performed feature selection using randomized lasso stability selection in each cohort (*monaLisa* package, ^41^). In a random subsample of n/2, ADHD diagnosis was regressed against all genera in a lasso-penalized regression (repeated for 999 subsamples). The selection probability was calculated as the number of permutations in which a genus was selected (i.e., β≠0) divided by the total number of permutations. Due to small individual sample sizes and high interindividual variability in the gut-microbiome, the selection probabilities per genus are expected to be small. We applied a lenient threshold of 10% stability selection probability within each study, to assure that genera with potentially small within-study relevance will be picked up, as they might accumulate across studies. We disregarded all genera from further analysis whose selection probability stability paths showed low to no informative value, see Supplementary Chapter 5.1. Selected genera, prevalent across cohorts were introduced to differential abundance analysis.

#### Differential abundance analysis

We applied differential abundance analysis per study, using logistic regressions associating ADHD diagnosis with the CLR-transformed abundance. We meta-analyzed over the standardized effect size (log odds ratio) of all four studies for each genus. To account for potential inconsistencies across statistical tools, we additionally performed differential abundance analysis with ALDEx2. We applied ALDEx2 per study before prevalence thresholding and feature selection, as the correction for feature variation is estimated most accurately taking all features into account. We subsequently estimated standardized effect sizes (standardized correlation coefficient R and variance) and performed meta-analysis on only the prevalent, feature selected genera. We applied a significance threshold of *p* < 0.05, where *p-values* were false discovery rate (fdr) corrected.

#### Associations with symptoms

To investigate if compositional differences in the gut-microbiome of adults with ADHD were associated with the symptoms of hyperactivity/impulsivity or inattention, we employed rank-based regression (*Rfit*) on the mean centered number of symptoms (see supplementary chapter 1.1) and the (CLR-transformed) abundance of those genera, that were robustly associated with ADHD diagnosis abundance, corrected for age and sex. We extracted the standardized effect size measure R (*metafor*) for each association per study and meta analyzed (N = 505). We applied a significance threshold of *p* < 0.05, *p-values* were false discovery rate (fdr) corrected.

## Results

### Alpha Diversity

We found no significant differences of alpha diversity between adults with and without ADHD, neither on the individual study level nor in the meta-analysis, in terms of observed ASV, Shannon index, or Faith’s phylogenic diversity, see Supplementary Chapter 3.

### Beta Diversity

At the individual study level, three out of four studies showed differences in beta diversity between people with and without ADHD (Supplementary Chapter 4). The Permanova over all studies showed a significant association of ADHD diagnosis with beta diversity (*p* = 4.7E^−2^, F=1.77) explaining 0.2% of variance, despite pronounced differences between the cohorts (explaining ca. 25%, *p* = 1.0E^−2^, F=67.94, Table 2). Figure 2 shows a separation of individuals with (blue) and without ADHD (red) as well as separation of MIND-Set and IMpACT2-NL, using the same sequencing technique and region, from the other two cohorts, which applied different wet-lab techniques.

**Figure 2.**
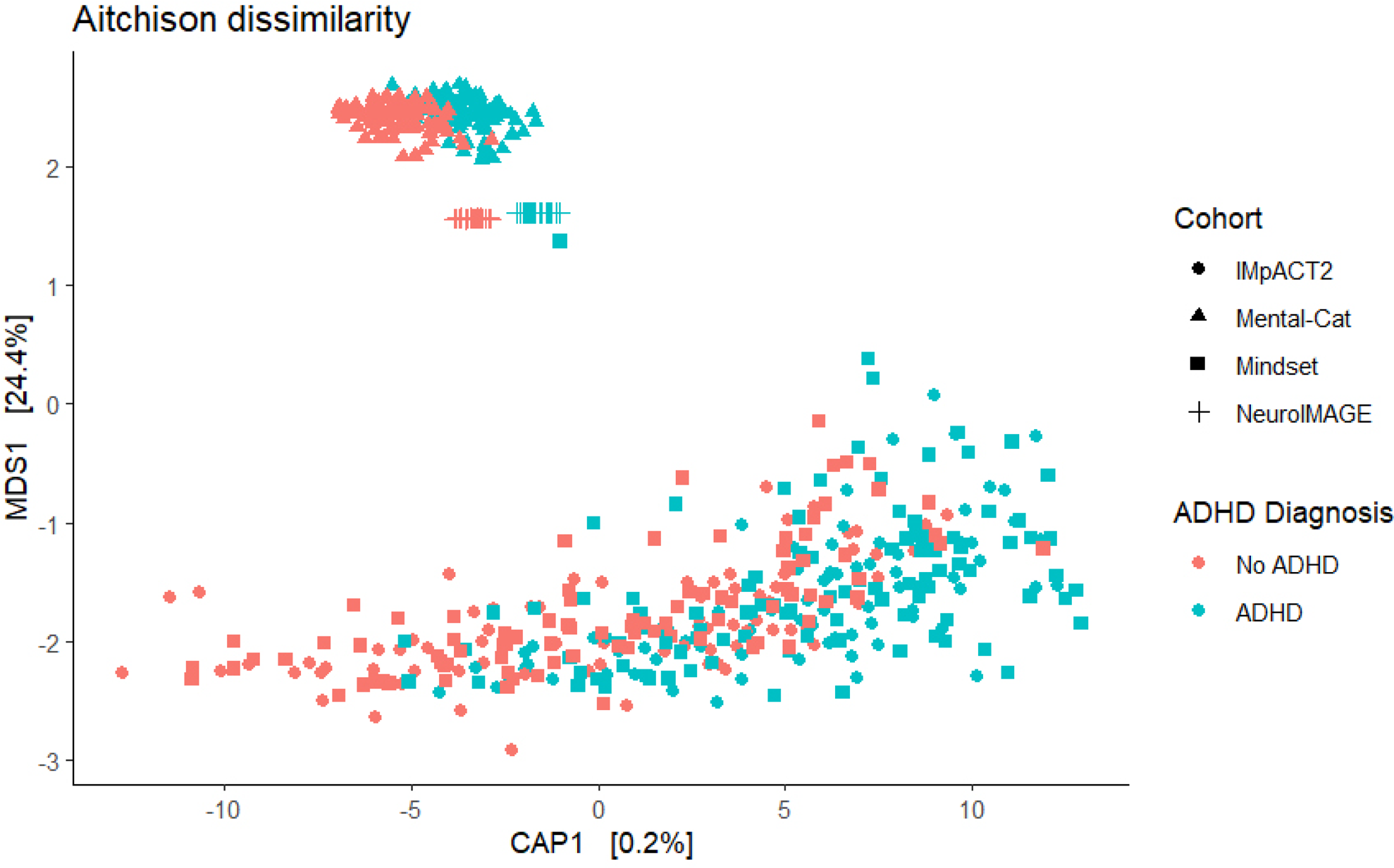
Beta-diversity of gut microbial communities in 312 people with ADHD (marked in blue) and 305 without ADHD (marked in red) over four cohorts. The CAP plot is supervised for differences in Aitchison dissimilarity between diagnostic groups (ADHD diagnosis), the cohort is marked by shapes (dot for IMpACT2-NL, triangle for MIND-Set, square for NeuroIMAGE and + for Mental-Cat).

**Table 2.** Beta Diversity results from Permanova with 999 permutations over the samples of all four studies combined.

| | Sum of Squares | $R^2$ | F | p-value |
| --- | --- | --- | --- | --- |
| ADHD diagnosis | 12890 | 0.002 | 1.77 | 4.7E-02 |
| Age | 23665 | 0.003 | 3.24 | 1.0E-03 |
| Sex | 11763 | 0.002 | 1.61 | 6.4E-02 |
| Cohort | 1486900 | 0.244 | 67.94 | 1.0E-03 |
| Residual | 4450154 | 0.731 |  |  |
| Total | 6087500 | 1 |  |  |

Table contains the sum of squares, the proportion of variance in beta diversity explained by the regressors diagnosis, age, sex, and effect of the study ( $R^2$ ), as well as test statistics F and permutation p-value.

### Composition

#### Feature selection

A total of 27 genera were selected over all four studies in the randomized Lasso stability selection, 20 of which exceeded the prevalence threshold of 10% in all four cohorts; see Supplementary Chapter 5 for the selected features, selection probability, and the stability paths per study.

#### Differential abundance analysis

Logistic regression-based meta-analyses of the 20 selected genera identified 5 significantly different genera between adults with and without ADHD (fdr-corrected): *Ruminococcus_torques_group* (*p*_fdr_=4.4E^−2^, Log odds ratio (LOR) = 0.17), *Eubacterium_xylanophilum_group* (*p*_fdr_=6.9E^−3^, LOR = −0.12), *Eubacterium_ruminantium_group* (*p*_fdr_=4.4E^−2^, LOR = −0.06), *Eisenbergiella* (*p*_fdr_=2.0E^−2^, LOR = 0.14), and *Clostridia_UCG_014* (*p*_fdr_=4.4E^−2^, LOR = −0.07). Figure 3 displays the effect size, direction of effects, and significance level for each genus.

**Figure 3.**
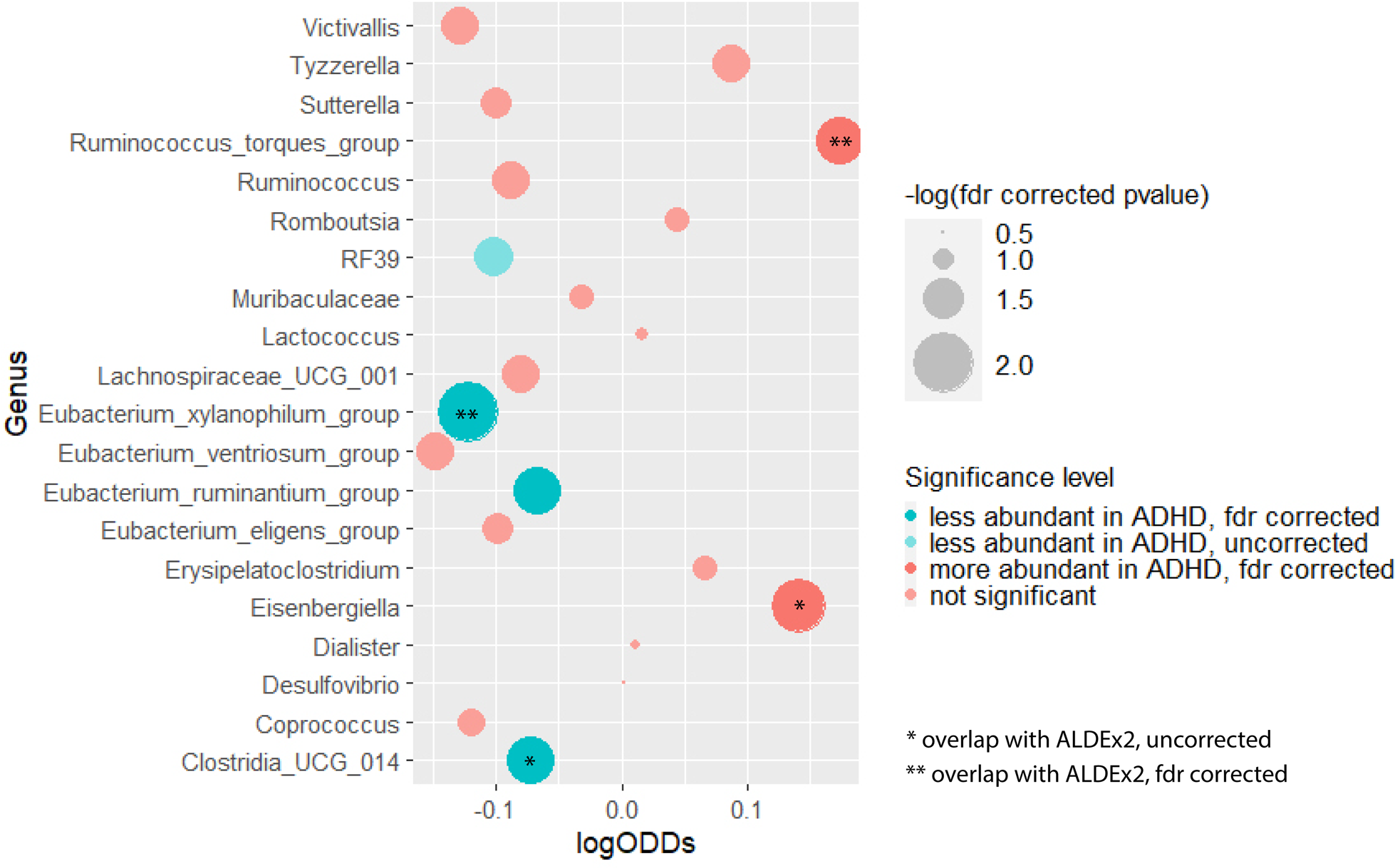
Results from the meta-analyses on all 21 genera (y-axis) from logistic regression, displaying significance level (bubble size), effect size (x-axis) and direction of effects (red for higher, blue for lower abundance in ADHD). Converging results with ALDEx2 are marked with *(uncorrected), **(fdr corrected).

By repeating the meta-analyses on the ALDEx2-based effects of the selected genera, we identified two fdr-corrected significantly different genera, both overlapping with the results from logistic regression; *Ruminococcus_torques_group* (*p*_fdr_=1.9E^−2^, *r*=0.13) was more abundant, while *Eubacterium_xylanophilum_group* (*p*_fdr_=1.9E^−2^, *r*=-0.12) was less abundant in people with ADHD compared to those without ADHD. At an uncorrected significance level, ALDEx2 identified four out of the five genera identified in logistic regression (along with *Eisenbergiella* and *Clostridia_UCG_014*), which were subsequently introduced to post-hoc symptom associations (see below). The significant results showed small effect sizes, with no significant influence of study heterogeneity. Forest plots of significant results, and results tables of the meta-analyses are described in Supplementary Chapter 6.1 and 6.2. We further visualized relative abundance and the number of non-zero samples for the 20 selected genera, see Supplementary Chapter 6.3.

#### Post-hoc associations with ADHD symptoms

More inattention symptoms were significantly associated with higher abundance of *Eisenbergiella* (*p*_fdr_=1.1E^−3^, *r*=0.16), and more hyperactivity/impulsivity symptoms were significantly associated with higher abundance of *Eisenbergiella* (*p*_fdr_=4.2E^−3^, *r*=0.14) and *Ruminococcus_torques_group* (*p*_fdr_=1.2E^−2^, *r*=0.13). The results of the meta-analyses are available in the Supplementary Chapter 7.1.

## Discussion

### Study summary and results

In study, we aimed to identify robust gut-microbiome alterations in adult ADHD by harmonizing the bioinformatic pipelines of four case-control cohorts (N=617), investigating diversity and differential abundance across tools and indices, correcting for common confounders per study, meta-analyzing and interpreting converging results across tools and studies. While alpha diversity was not significantly different between adults with and without ADHD, we found differences in beta diversity and identified robust microbiome-compositional alterations in adult ADHD and associations with the severity of inattention and hyperactivity/impulsivity symptoms.

Interestingly, beta diversity was additionally influenced by the individual cohort setup, despite harmonized preprocessing. The clustering of two studies with overlapping wet-lab procedures and separation of two studies who applied different wet-lab strategies underlines the impact on detected composition and supports the need for meta-analytic approaches and harmonized wet-lab protocols to account for influences of collection, storing and sequencing. While diversity is an unspecific measure for disease-related processes, indicating only global tendencies of potentially disrupted ecosystems, compositional differences can add information on individual features involved and help identifying relevant functional pathways.

On a compositional level, *Eubacterium_xylanophilum_group* was significantly less abundant in adults with ADHD, while *Ruminococcus_torques_group* was more abundant and associated with more hyperactivity/impulsivity symptoms, converging over meta-analyses. Additionally, both methods identified higher abundance of *Eisenbergiella*, associated with hyperactivity/impulsivity and inattention symptoms, and lower abundance of *Clostridia_UCG_014* in adults with ADHD.

### Functional properties

*The Ruminococcus_torques_group* has earlier been found enriched in children with ASD ^42^, Irritable Bowel Syndrome ^43^, Crohn’s disease ^44^, and influenza-like illness ^45^; it was also found associated with the degradation of the intestinal mucosal layer ^46^. *Ruminococcus_torques* species were further associated with a western diet (high energy, low nutrition) and neuroinflammation; it might also be related to reduction in striatal dopamine in Parkinson’s Disease (for a review, see ^47^). These studies suggest a role of *Ruminococcus_torques_group* in gut-barrier functioning and pro-inflammatory processes, which have previously been implicated in the etiology of psychiatric, neurodevelopmental, and neurological disorders ^48, 49^.

*Eisenbergiella* has also been found enriched in individuals with neuropsychiatric disorders, such as ASD, depression, and Parkinson’s disease ^50–52^, in chickens and mice infected with pathogens ^53, 54^, and in children with an allergy to cow’s milk ^55^. It was found reduced in rats after immunosuppressive/anti-inflammatory treatment ^56^, suggesting general associations with pro-inflammatory processes and immune activation. Higher abundance of *Eisenbergiella* has also been linked to a high energy diet (rich in carbohydrates, fat, and protein) ^57^ and to metabolic disorders, such as gestational diabetes mellitus ^58^.

*Eubacterium_xylanophilum_group* seems to play an anti-inflammatory role. It is considered a producer of SCFAs and was found enriched after intervention with polyphenols in piglets ^59^. Metabolism of polyphenols into SCFAs by the gut-microbiome is discussed as a potential mechanism through which polyphenols might unfold their anti-inflammatory properties ^60^. *Eubacterium_xylanophilum_group* was also found enriched in mice with colorectal cancer after supplementation with sodium butyrate, associated with SCFA production and an improved immune response ^61^. Associations of *Eubacterium_xylanophilum_group* with beneficial effects on immune functioning might be relevant for ADHD, potentially supporting favorable health outcomes, but these interpretations are highly speculative, as human and functional studies are lacking.

In summary, the idea that *Ruminococcus_torques_group* and *Eisenbergiella* could play a role in the pathophysiology of adult ADHD is supported by the described associations with other brain disorders characterized by altered monoamine neurotransmission (e.g. ASD, depression, Parkinson’s disease) and by the observed influences on inflammation and immune functioning. Immune activation, for example caused by increased intestinal permeability, is considered as a potential mechanism causing and maintaining psychiatric and somatic symptoms, reflecting in shared genetic risk of immune and psychiatric disorders^62^

Notably, none of these genera had been reported in any previous studies of gut microbiome composition in ADHD (see table ST1), potentially due to the differences in origin, developmental stage and power issues of the published studies.

### Strengths and Limitations

The results of this study have to be viewed in the light of its strengths and limitations. Even though we provide a comparably big sample size, the expected small effect sizes and high inter- and intra-individual variability in gut-microbial signatures require replication of these findings in even larger samples. Collecting data from different studies comes with limitations of the included individual studies (quality, technical variation). We reduced and accounted for effects of individual study differences by harmonizing pipelines and using random effects meta-analyses. However, different sequencing region, for example, might still limit the detection of the same genera across studies. Inconsistently detected genera would not be considered or introduced to meta-analysis.

All included datasets were based on 16S sequencing, which picks up on low abundant features, but provides comparably low resolution. The taxonomic identification at this resolution does not provide information about functional properties of the identified taxa. Discussions of potential disorder-related mechanistic pathways (e.g. a potential role of inflammatory processes in ADHD) or functional properties of genus-level data are based on a narrative summary of associations with other phenotypes in the literature, and are therefore necessarily speculative and unspecific to ADHD. The gut-microbiome is sensitive to environmental, behavioral, and dietary changes. For people with psychiatric disorders, disorganized thoughts and behaviors or impulsive food choices, for example, might be responsible for the observed difference in abundance of particular genera. Similarly, the decrease of *Eisenbergiella* after immune-suppressive treatment supports the idea that - rather than a cause - increases in abundance could be seen as an epiphenomenon or consequence of inflammation. Through the association studies performed here, we therefore cannot give insights on causality or consequences of the differential microbial abundance in ADHD.

While we corrected for age and sex effects on the gut-microbiome, diet information was only provided for IMpACT2-NL and MIND-Set. After post-hoc analyses, correcting for diet on an individual study level in MIND-Set and IMpACT2-NL, all significant associations remained significant, see Supplementary Chapter 7.2. However, diet was significantly associated with ADHD diagnosis, confirming the relevance of diet as either behavioral epiphenomenon of or influence on ADHD. Dietary habits should be assessed thoroughly using standardized tools and accounted for in each study. Another important confounder is medication. Drugs can impact the gut-microbiome; in turn, the gut-microbiome might also impact the metabolism of medications and modulate the treatment response (for an overview, see ^63^). However, in psychiatric case-control cohorts, pharmacological treatment is highly colinear with the diagnostic group, and issues like multi-medication resulting from high comorbidity and treatment adherence further complicate this picture. Significant contributions of the medication naïve sample (Mental-Cat) as well as the highly medicated sample (MIND-Set), point towards low sensitivity of these results for ADHD medication. Post-hoc analyses of medication effects was performed on cases only in IMpACT2-NL and NeuroIMAGE. Eisenbergiella was more abundant in participants with ADHD in the NeuroIMAGE sample, but not on IMpACT and no other association of genus abundance with current current use of ADHD medication was found, see Supplementary Chapter 7.3. The interplay of gut-microbiota with psychopharmacological treatment should be investigated in future studies.

### Conclusion and future directions

In summary, we identified alterations of the gut-microbiome in adult ADHD that were robust to statistical approach and study heterogeneity. The ADHD-associated genera suggested potential relevance of inflammatory and immune processes for ADHD symptoms. However, more human and functional studies are needed to support potential interpretations.

To exhaust the potential of 16S sequencing studies and increase comparability we emphasize the need for standardized pre-processing and statistical pipelines, large samples, and deep phenotyping. To extend this work to the lifespan of ADHD, future studies should integrate studies in children, adolescents, and adults in meta-regression or in longitudinal designs. These approaches could account for natural changes in the gut-microbiome in the different developmental stages and help to provide insight in potential influences of the gut-microbiome in early life on later neurodevelopment within ADHD (e.g. to identify potential remission profiles). To distinguish disorder-specific, symptom-/trait-specific, and transdiagnostic effects, studies across psychiatric categories and in population-based cohorts are needed. Additionally, the combination of 16S studies with metagenomic sequencing, clustering of sequences based on their involvement in functional mechanistic pathways as well as wet-lab culturing studies investigating functions of the bacteria are needed to gain mechanistic insights. Intervention studies, targeting the reduction of genera enriched in ADHD (and/or the enrichment of reduced genera), could be implemented to infer causal relationships between ADHD symptoms and gut-microbial alterations; such studies could evaluate the potential of the gut-microbiome as a biomarker as well as for treatment support. Investigating the gut-microbial changes in pharmacological randomized control trials could help disentangle disorder-related effects from medication-related effects, additionally providing a perspective to improve treatment response.

## Data Availability

All data produced in the present study are available upon reasonable request to the authors

## Acknowledgements

NeuroIMAGE: This project was supported by grants from National Institutes of Health (grant R01MH62873 to SV Faraone) for initial sample recruitment, and from NWO Large Investment (grant 1750102007010 to JK Buitelaar), NWO Brain & Cognition (grant 433-09-242 to JK Buitelaar), and grants from Radboud University Medical Center, University Medical Center Groningen, Accare, and VU University Amsterdam for subsequent assessment waves. NeuroIMAGE also received funding from the European Community’s Seventh Framework Programme (FP7/2007 – 2013) under grant agreements n° 602805 (Aggressotype), n° 278948 (TACTICS), and n° 602450 (IMAGEMEND), and from the European Community’s Horizon 2020 Programme (H2020/2014 – 2020) under grant agreements n° 643051 (MiND), n° 667302 (CoCA), and n° 728018 (Eat2beNICE).

IMpACT: We acknowledge funding from the Netherlands Organization for Scientific Research (NWO), i.e. the Veni Innovation Program (grant 016-196-115 to MH) and the Vici Innovation Program (grant 016–130-669 to BF). The work was also supported by the European College of Neuropsychopharmacology (ECNP) Network “ADHD Across the Lifespan”.

BJ and BF were also supported by funding from the European Community’s Horizon 2020 Programme (H2020/2014 – 2020) under grant agreement n° 847879 (PRIME).

## References

1. Faraone SV. ADHD. Nature Reviews Disease Primers 2015; 15027.

2. Kittel-Schneider S, Arteaga-Henriquez G, Vasquez AA, Asherson P, Banaschewski T, Brikell I et al. Non-mental diseases associated with ADHD across the lifespan: Fidgety Philipp and Pippi Longstocking at risk of multimorbidity? Neuroscience & Biobehavioral Reviews 2021.

3. Franke B, Michelini G, Asherson P, Banaschewski T, Bilbow A, Buitelaar JK et al. Live fast, die young? A review on the developmental trajectories of ADHD across the lifespan. European Neuropsychopharmacology 2018; 28(10): 1059–1088.

4. Núñez-Jaramillo L, Herrera-Solís A, Herrera-Morales WV. ADHD: reviewing the causes and evaluating solutions. Journal of personalized medicine 2021; 11(3): 166.

5. Warner BB. The contribution of the gut microbiome to neurodevelopment and neuropsychiatric disorders. Pediatric Research 2019; 85(2): 216–224.

6. Bull-Larsen S, Mohajeri MH. The potential influence of the bacterial microbiome on the development and progression of ADHD. Nutrients 2019; 11(11): 2805.

7. Dam SA, Mostert JC, Szopinska-Tokov JW, Bloemendaal M, Amato M, Arias-Vasquez A. The role of the gut-brain axis in attention-deficit/hyperactivity disorder. Gastroenterology Clinics 2019; 48(3): 407–431.

8. Liu F, Li J, Wu F, Zheng H, Peng Q, Zhou H. Altered composition and function of intestinal microbiota in autism spectrum disorders: a systematic review. Translational psychiatry 2019; 9(1): 1–13.

9. Nikolova VL, Hall MR, Hall LJ, Cleare AJ, Stone JM, Young AH. Perturbations in gut microbiota composition in psychiatric disorders: a review and meta-analysis. JAMA psychiatry 2021; 78(12): 1343–1354.

10. Karlsson F, Tremaroli V, Nielsen J, Bäckhed F. Assessing the human gut microbiota in metabolic diseases. Diabetes 2013; 62(10): 3341–3349.

11. Ding J-H, Jin Z, Yang X-X, Lou J, Shan W-X, Hu Y-X et al. Role of gut microbiota via the gut-liver-brain axis in digestive diseases. World Journal of Gastroenterology 2020; 26(40): 6141.

12. Checa-Ros A, Jeréz-Calero A, Molina-Carballo A, Campoy C, Muñoz-Hoyos A. Current evidence on the role of the gut microbiome in ADHD pathophysiology and therapeutic implications. Nutrients 2021; 13(1): 249.

13. Shirvani-Rad S, Ejtahed H-S, Ettehad Marvasti F, Taghavi M, Sharifi F, Arzaghi SM et al. The Role of Gut Microbiota-Brain Axis in Pathophysiology of ADHD: A Systematic Review. Journal of Attention Disorders 2022: 10870547211073474.

14. Sukmajaya AC, Lusida MI, Setiawati Y. Systematic review of gut microbiota and attention-deficit hyperactivity disorder (ADHD). Annals of general psychiatry 2021; 20(1): 1–12.

15. Bundgaard-Nielsen C, Knudsen J, Leutscher PD, Lauritsen MB, Nyegaard M, Hagstrøm S et al. Gut microbiota profiles of autism spectrum disorder and attention deficit/hyperactivity disorder: A systematic literature review. Gut Microbes 2020; 11(5): 1172–1187.

16. Gkougka D, Mitropoulos K, Tzanakaki G, Panagouli E, Psaltopoulou T, Thomaidis L et al. Gut microbiome and attention deficit/hyperactivity disorder: a systematic review. Pediatric Research 2022: 1–13.

17. Wan L, Ge W-R, Zhang S, Sun Y-L, Wang B, Yang G. Case-control study of the effects of gut microbiota composition on neurotransmitter metabolic pathways in children with attention deficit hyperactivity disorder. Frontiers in Neuroscience 2020; 14: 127.

18. Jiang H-y, Zhou Y-y, Zhou G-l, Li Y-c, Yuan J, Li X-h, et al. Gut microbiota profiles in treatment-naïve children with attention deficit hyperactivity disorder. Behavioural brain research 2018; 347: 408–413.

19. Richarte V, Sánchez-Mora C, Corrales M, Fadeuilhe C, Vilar-Ribó L, Arribas L et al. Gut microbiota signature in treatment-naive attention-deficit/hyperactivity disorder. Translational psychiatry 2021; 11(1): 1–7.

20. Aarts E, Ederveen TH, Naaijen J, Zwiers MP, Boekhorst J, Timmerman HM et al. Gut microbiome in ADHD and its relation to neural reward anticipation. PloS one 2017; 12(9): e0183509.

21. Szopinska-Tokov J, Dam S, Naaijen J, Konstanti P, Rommelse N, Belzer C, et al. Correction: Szopinska-Tokov, et al. Investigating the Gut Microbiota Composition of Individuals with Attention-Deficit/Hyperactivity Disorder and Association with Symptoms. Microorganisms 2020, 8, 406. Microorganisms 2021; 9(7): 1358.

22. Hiergeist A, Gessner J, Gessner A. Current limitations for the assessment of the role of the gut microbiome for attention deficit hyperactivity disorder (ADHD). Frontiers in Psychiatry 2020; 11: 623.

23. Abellan-Schneyder I, Matchado MS, Reitmeier S, Sommer A, Sewald Z, Baumbach J et al. Primer, pipelines, parameters: issues in 16S rRNA gene sequencing. Msphere 2021; 6(1): e01202–01220.

24. Gloor GB, Macklaim JM, Pawlowsky-Glahn V, Egozcue JJ. Microbiome datasets are compositional: and this is not optional. Frontiers in microbiology 2017; 8: 2224.

25. Nearing JT, Douglas GM, Hayes MG, MacDonald J, Desai DK, Allward N et al. Microbiome differential abundance methods produce different results across 38 datasets. Nature communications 2022; 13(1): 1–16.

26. Gloor GB, Reid G. Compositional analysis: a valid approach to analyze microbiome high-throughput sequencing data. Canadian journal of microbiology 2016; 62(8): 692–703.

27. van Eijndhoven P, Collard R, Vrijsen J, Geurts DE, Vasquez AA, Schellekens A et al. Measuring Integrated Novel Dimensions in Neurodevelopmental and Stress-Related Mental Disorders (MIND-SET): protocol for a cross-sectional comorbidity study from a Research Domain Criteria perspective. JMIRx Med 2022; 3(1): e31269.

28. Jakobi B, Arias-Vasquez A, Hermans E, Vlaming P, Buitelaar JK, Franke B et al. Neural Correlates of Reactive Aggression in Adult ADHD. Frontiers in psychiatry 2022: 1019.

29. von Rhein D, Mennes M, van Ewijk H, Groenman AP, Zwiers MP, Oosterlaan J et al. The NeuroIMAGE study: a prospective phenotypic, cognitive, genetic and MRI study in children with attention-deficit/hyperactivity disorder. Design and descriptives. European child & adolescent psychiatry 2015; 24(3): 265–281.

30. Mirzayi C, Renson A, Zohra F, Elsafoury S, Geistlinger L, Kasselman LJ et al. Reporting guidelines for human microbiome research: the STORMS checklist. Nature medicine 2021; 27(11): 1885–1892.

31. Bolyen E, Rideout JR, Dillon MR, Bokulich NA, Abnet CC, Al-Ghalith GA et al. Reproducible, interactive, scalable and extensible microbiome data science using QIIME 2. Nature biotechnology 2019; 37(8): 852–857.

32. Callahan BJ, McMurdie PJ, Rosen MJ, Han AW, Johnson AJA, Holmes SP. DADA2: High-resolution sample inference from Illumina amplicon data. Nature methods 2016; 13(7): 581–583.

33. Ewing B, Green P. Base-calling of automated sequencer traces using phred. II. Error probabilities. Genome research 1998; 8(3): 186–194.

34. Price MN, Dehal PS, Arkin AP. FastTree 2–approximately maximum-likelihood trees for large alignments. PloS one 2010; 5(3): e9490.

35. R Core Team R, Team RC. R: a language and environment for statistical computing. R Foundation for Statistical Computing; 2020. 2021.

36. Lahti L, Shetty S. Tools for microbiome analysis in R. Microbiome package version 1.7. 21. Bioconductor Available at: http://microbiomegithubcom/microbiome[Google Scholar] 2017.

37. Kloke JD, McKean JW. Rfit: rank-based estimation for linear models. R J 2012; 4(2): 57.

38. Balduzzi S, Rücker G, Schwarzer G. How to perform a meta-analysis with R: a practical tutorial. BMJ Ment Health 2019; 22(4): 153–160.

39. Oksanen J, Blanchet F, Friendly M. vegan: community ecology package. R package version 2.4–4.2. 2017.

40. McMurdie PJ, Holmes S. phyloseq: an R package for reproducible interactive analysis and graphics of microbiome census data. PloS one 2013; 8(4): e61217.

41. Meinshausen N, Bühlmann P. Stability selection. Journal of the Royal Statistical Society: Series B (Statistical Methodology) 2010; 72(4): 417–473.

42. Wang L, Christophersen CT, Sorich MJ, Gerber JP, Angley MT, Conlon MA. Increased abundance of Sutterella spp. and Ruminococcus torques in feces of children with autism spectrum disorder. Molecular autism 2013; 4(1): 1–4.

43. Malinen E, Krogius-Kurikka L, Lyra A, Nikkilä J, Jääskeläinen A, Rinttilä T et al. Association of symptoms with gastrointestinal microbiota in irritable bowel syndrome. World journal of gastroenterology: WJG 2010; 16(36): 4532.

44. Joossens M, Huys G, Cnockaert M, De Preter V, Verbeke K, Rutgeerts P et al. Dysbiosis of the faecal microbiota in patients with Crohn’s disease and their unaffected relatives. Gut 2011; 60(5): 631–637.

45. Fuentes S, den Hartog G, Nanlohy NM, Wijnands L, Ferreira JA, Nicolaie MA, et al. Associations of faecal microbiota with influenza-like illness in participants aged 60 years or older: an observational study. The Lancet Healthy Longevity 2021; 2(1): e13–e23.

46. Hoskins LC, Agustines M, McKee WB, Boulding ET, Kriaris M, Niedermeyer G. Mucin degradation in human colon ecosystems. Isolation and properties of fecal strains that degrade ABH blood group antigens and oligosaccharides from mucin glycoproteins. The Journal of clinical investigation 1985; 75(3): 944–953.

47. Hamamah S, Hajnal A, Covasa M. Impact of Nutrition, Microbiota Transplant and Weight Loss Surgery on Dopaminergic Alterations in Parkinson’s Disease and Obesity. International journal of molecular sciences 2022; 23(14): 7503.

48. Pellegrini C, Fornai M, D’Antongiovanni V, Antonioli L, Bernardini N, Derkinderen P. The intestinal barrier in disorders of the central nervous system. The Lancet Gastroenterology & Hepatology 2022.

49. Bauer ME, Teixeira AL. Inflammation in psychiatric disorders: what comes first? Annals of the New York Academy of Sciences 2019; 1437(1): 57–67.

50. Ye F, Gao X, Wang Z, Cao S, Liang G, He D et al. Comparison of gut microbiota in autism spectrum disorders and neurotypical boys in China: a case-control study. Synthetic and Systems Biotechnology 2021; 6(2): 120–126.

51. Comparison of Metabolites and Gut Microbes between Patients with Parkinson’s Disease and Healthy Individuals—A Pilot Clinical Observational Study (STROBE Compliant). Proceedings of the Healthcare 2022. MDPI.

52. Fontana A, Manchia M, Panebianco C, Paribello P, Arzedi C, Cossu E et al. Exploring the role of gut microbiota in major depressive disorder and in treatment resistance to antidepressants. Biomedicines 2020; 8(9): 311.

53. Bao J, Zheng H, Wang Y, Zheng X, He L, Qi W et al. Echinococcus granulosus infection results in an increase in Eisenbergiella and Parabacteroides genera in the gut of mice. Frontiers in Microbiology 2018; 9: 2890.

54. Li H-Y, Zhang H-L, Zhao F-J, Wang S-Q, Wang Z-X, Wei Z-Y. Modulation of gut microbiota, short-chain fatty acid production, and inflammatory cytokine expression in the cecum of porcine Deltacoronavirus-infected chicks. Frontiers in microbiology 2020; 11: 897.

55. Mauras A, Wopereis H, Yeop I, Esber N, Delannoy J, Labellie C et al. Gut microbiota from infant with cow’s milk allergy promotes clinical and immune features of atopy in a murine model. Allergy 2019; 74(9): 1790.

56. Zhang J, Feng D, Law HK-W, Wu Y, Zhu G-h, Huang W-y, et al. Integrative Analysis of Gut Microbiota and Fecal Metabolites in Rats after Prednisone Treatment. Microbiology spectrum 2021; 9(3): e00650–00621.

57. Surono IS, Jalal F, Bahri S, Romulo A, Kusumo PD, Manalu E et al. Differences in immune status and fecal SCFA between Indonesian stunted children and children with normal nutritional status. PloS one 2021; 16(7): e0254300.

58. Ma S, You Y, Huang L, Long S, Zhang J, Guo C et al. Alterations in gut microbiota of gestational diabetes patients during the first trimester of pregnancy. Frontiers in cellular and infection microbiology 2020; 10: 58.

59. Hu R, Wu S, Li B, Tan J, Yan J, Wang Y et al. Dietary ferulic acid and vanillic acid on inflammation, gut barrier function and growth performance in lipopolysaccharide-challenged piglets. Animal Nutrition 2022; 8(1): 144–152.

60. Diotallevi C, Fava F, Gobbetti M, Tuohy K. Healthy dietary patterns to reduce obesity-related metabolic disease: polyphenol-microbiome interactions unifying health effects across geography. Current Opinion in Clinical Nutrition & Metabolic Care 2020; 23(6): 437–444.

61. Ma X, Zhou Z, Zhang X, Fan M, Hong Y, Feng Y et al. Sodium butyrate modulates gut microbiota and immune response in colorectal cancer liver metastatic mice. Cell Biology and Toxicology 2020; 36(5): 509–515.

62. Rudzki L, Szulc A. “Immune gate” of psychopathology—The role of gut derived immune activation in major psychiatric disorders. Frontiers in psychiatry 2018; 9: 205.

63. Maier L, Typas A. Systematically investigating the impact of medication on the gut microbiome. Current opinion in microbiology 2017; 39: 128–135.

