## Supplementary Material for "The gut-microbiome in adult Attention-deficit/hyperactivity disorder - A Meta-analysis"

1 Study identification and description

We identified potentially relevant studies by screening recent systematic reviews [13, 14] and through literature search for original case-control studies including fecal microbiome samples from participants with and without ADHD diagnoses. The flowchart in Figure 15 shows the identified studies and an example of the systematic search for 1^st^ of January 2022, yielding the same results. Supplementary Table 1 summarizes the studies that were contacted with requests for the raw data and summarizes their main results.

Supplementary Table 1: Summary of gut-microbiome studies in ADHD

| Authors | Year | Country | Sequencing | Sample size  ADHD:Control | Age | Alpha diversity | Beta diversity | Genus level differential abundance |
| --- | --- | --- | --- | --- | --- | --- | --- | --- |
| **Published case-control comparison studies** | | | | | | | | |
| Richarte et al. | 2021 | Spain | 16S V3/V4 | 100:100 | Adults | No difference | No difference | *Dialister, Megamonas* up *Gracilibacter, Anaerotaenia* down |
| Szopinska-Tokov et al. | 2020 | Netherlands | 16S V1/V2 | 41:48 | Adolescents - adults | No difference | No difference | *Intestinibacter* up, C*oprococcus_2, Prevotella_9* down |
| Aarts et al. | 2017 | Netherlands | 16S V1/V2 | 19:77 | Adolescents - adults | No difference | NA | *Bifidobacterium* up |
| Wan et al. | 2020 | China | Shotgun metagenomics | 17:17 | Children | No difference | NA | ***Faecalibacterium* down**, *Veillonellaceae* down, *Odoribacter* up, *Enterococcus* up |
| Wang et al. | 2020 | Taiwan | 16S V3/V4 | 30:30 | Children | Simpson down, Shannon up, Chao2 up | NA | *Lactobacillus* up, *Fusobacterium* up |
| Prehn-Kristensen et al. | 2018 | Germany | 16S V1/V2 | 14:17 | Children | Shannon down | Difference | *Prevotella, Parabacteroides* down, *Neisseria* up |
| Jiang et al. | 2018 | China | 16S V3/V4 | 51:32 | Children | No difference | No difference | ***Faecalibacterium* down**, *Lachnoclostridium, Dialister* down |
| Signals across cohorts (meta-analytic approach) | | | | | | | | |
| Wang et al. | 2022 | Netherlands, Germany, China, Taiwan | 16S (V1-V2, V3-V4, shotgun metagenomics | 2 to 5 studies per index (alpha diversity)  2 to 3 studies per genus (differential abundance | Across age groups | No difference | Not assessed | *Blautia* (across three individual studies) |
| Non-peer reviewed reports/preprints | | | | | | | | |
| Li et al. | 2020 | China | metagenomics | 98:109 | Children | Lower observed features, Shannon no difference | No difference | No genus level results but species belonging to *Bacteroides, Prevotella* and *Bifidobacterium* genus |
| Fan et al. | 2019 | China | 16S V3/V4 | 49:24 | Children | lower across indices in inattention |  | No overall case-control differences but *Prevotella, Intestinimonas, Marvinbryantia, Eggerthella* (hyperactivity), 14 genera (inattention), *Megamonas, Corprococcus_2, Paraprevotella* (combined) |
| Akram et al. | 2017 | NA | 16S V3/V4 | 14:20 | Young adults | NA | NA | *Phascolarctobacterium, Paraprevotella, Veillonella, Odoribacter* |

*Summary of results of previous studies on the fecal gut-microbiome samples in people with and without ADHD, including description of the study population and summary of the main findings, synthesized from recent systematic reviews on the gut-microbiome in ADHD [13, 14]. Data of the grey-shaded studies was included in this meta-analysis, data of all studies was requested.. Results of the non-peer-reviewed studies were not interpreted in the manuscript.*

1.1 Recruitment and inclusion criteria per study

The Mental-Cat cohort consists of 100 treatment naïve adults with and 100 adults without ADHD. Participants with ADHD were recruited from an outpatient program in Catalonia, Spain, by a clinical group from the Hospital Universitari Vall d’Hebron of Barcelona, Spain. The study was approved by the Clinical Research Ethics Committee (CREC) of Hospital Universitari Vall d’Hebron.  Exclusion criteria: intelligence quotient < 70, current or past psychiatric, neurologic or systemic disorders, other developmental disorders, current or past neurologic, metabolic, cardiac, liver, kidney, or respiratory conditions, chronic pharmacological treatment of any kind, birth weight ≤ 1.5 kg, treatment with probiotics or antibiotics prior to fecal sample collection. For the control group, participants with ADHD or other psychiatric disorders (current or past) were excluded. Assessment of **ADHD symptoms** was conducted with the Adult Self-Report Scale A.S.R. S v1.1 in the participants with ADHD only [64]. This scale consists of 18 questions about frequency of symptoms of adult ADHD in the scale of never, rarely, sometimes, often and very often. 7 of these questions are scored with 1 when answered with often or very often, the remaining 11 questions also if the answer is sometimes. See [65] for further information.

The IMpACT2-NL cohort consists of 83 adults with and 79 adults without ADHD, of which 77 with and 79 without ADHD provided fecal samples. Participants were recruited via local newspapers, advertisements in sports clubs in and around Nijmegen, The Netherlands, and via patient organizations. Participants received monetary compensation and gave written informed consent. The study was approved by the local medical ethical committee (Central Commission for Human Rights Research (CCMO)). Exclusion criteria for all participants comprised: younger than 18 or older than 60 years, neurological disorders, psychosis or substance abuse in the last 6 months, current major depression, psycho-pharmaceutical therapy (other than stimulants for participants with ADHD), impairments of hearing, seeing and sensorimotor abilities and no knowledge of the Dutch language. Exclusion criteria in the control group comprised no current or previous diagnoses of ADHD, and no first-degree family members with ADHD. Assessment of **ADHD symptoms** was conducted with the Diagnostic Interview for Adult ADHD (DIVA 2.0 [66]], based on the Diagnostic and Statistical Manual of Mental Disorders (DSM), assessing the presence or absence of 9 symptoms of inattention and 9 symptoms of hyperactivity/impulsivity providing every-day examples for each symptom. See [28] for further information.

The **MIND-Set** cohort comprises ca 650 individuals with a psychiatric disorder and 150 neurotypical controls (by 2021). Oh those, 104 individuals with a diagnosis of ADHD and 90 neurotypical individuals provided fecal samples. Participants were recruited at the inpatient and outpatient unit of the psychiatric department of the Radboud University Medical Center (Radboudumc), Nijmegen, the Netherlands, provided written informed consent and received monetary compensation. The study was approved by the local medical ethical committee (Commissie Mensgebonden Onderzoek Arnhem-Nijmegen). Individuals of at least 18 years were included in the neurotypical control group if they had no psychiatric diagnoses, or in the psychiatric patient group if they had a diagnosis of a stress-related (mood disorder, anxiety disorder, or substance use disorder) or neurodevelopmental disorder (ASD or ADHD). In this analyses, only participants with ADHD were included, presenting with a high level of comorbidity (84% had another psychiatric diagnosis), and often pharmacological treatment with several medications (most frequently ADHD medication and antidepressants). Exclusion criteria for the psychiatric patient group were diseases of the central nervous system, (permanent) sensorimotor or (neuro)cognitive impairments, current psychosis, IQ <70, no knowledge of the Dutch language. **ADHD symptoms** were assessed with the CAARS-S:S, a short self-report rating scale covering the frequency of symptoms of Inattention/ Memory Problems (five items), Hyperactivity/Restlessness (five items) and Impulsivity/Emotional Lability (five items), scored on a 4-point scale (0 = never, 1 = sometimes, 2 = often, and 3 = very often). We used the mean of Impulsivity/Emotional Lability and Hyperactivity/Restlessness symptoms as a measure of Hyperactivity/Impulsivity symptoms.

The **NeuroIMAGE** cohort is a Dutch multicenter study that recruited families with members with ADHD and families without members with ADHD in Nijmegen and Amsterdam, The Netherlands. The study was approved by the regional ethics committee (Centrale Commissie Mensgebonden Onderzoek: CMO Regio Arnhem Nijmegen; 2008/163; ABR: NL23894.091.08). A small subset of the study-arm in Nijmegen provided fecal samples: 41 participants with ADHD, 48 healthy controls as well as 15 participants with so-called subthreshold ADHD (e.g. control participants with high symptom scores or individuals with an ADHD diagnosis who scored low on symptom scales). For more information on the definition of ADHD in NeuroIMAGE, see [67]. Inclusion criteria were: An age of 5 to 30 years, European Caucasian descent, IQ ≥70, no diagnosis of ASD, epilepsy, neurological disorders and learning disabilities. Only adult participants with unquestionable ADHD status were included in this study. **ADHD symptom** scores were derived from the K-SADS and the different versions of Conners’ rating scale (in adults the CAARS:L, the long version of the self-report), including the DSM-5 subscales for Inattentive and Hyperactive/Impulsive symptoms, for a detailed description of the calculation of symptom scores, see [67].

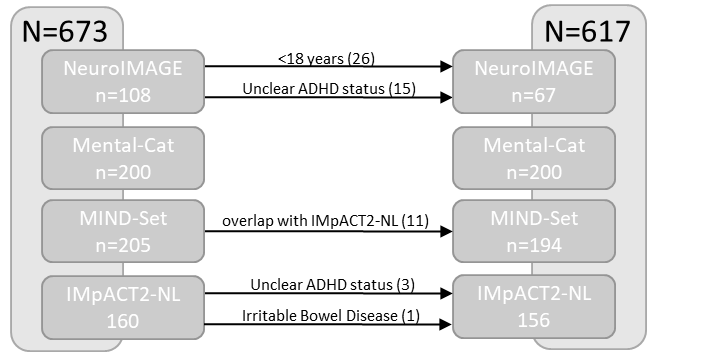

*Supplementary Figure 1. Flowchart of reasons for exclusions of participants from this study.*

2 Sample collection, storing and sequencing

For detailed information on the sample collection, storing, and sequencing of the NeuroIMAGE and VHIR cohorts please refer to the original papers [19, 20].

The fecal samples of the MIND-Set and IMpACT study were collected, stored, and processed according to our lab-routine (as described in Bloemendaal et al, 2022). Participants were instructed to collect their fecal samples at home using a validated protocol by OMNIgene•GUT kit (DNAGenotek, Ottawa, CA) and send them back to our laboratory. Samples received in good order were aliquoted into 1.5 ml Eppendorf tubes and stored in -80˚C. Further processing was done by Baseclear B.V by aliquoting 150 mg feces, isolating and purifying DNA using a bead-beating procedure, using ZymoBIOMICS DNA 96 MagBead kit in conjunction with Kingfisher. To identify bacterial DNA, the V4 region of 16S ribosomal RNA (rRNA) gene was targeted. Amplicons were built using the primers 515-F: TCGTCGGCAGCGTCAGATGT-GTATAAGACAGGTGYCAGCMGCCGCGGTAA, 806Rb: GTCTCGTGGGCTCGGAGATGTGTATAAGACAGAGG-GACTACNVGGGTWTCTAAT) using the Phusion High-Fidelity PCR Master Mix with an HF Buffer (Thermo Scientific). Illumina indexes were then ligated to the amplicons. The libraries were sequenced on the Illumina Novaseq 6000 platform with the Novaseq 6000 SP reagent kit v1 with 500 cycles (paired-end, 250 bp)(Baseclear B.V., Leiden, the Netherlands). Both a PCR negative control (water control for PCR and library preparation) was included sample to assess contamination introduced during the PCR step as well as a positive control (ZymoBIOMICS Gut Microbiome Stdrd (ZY-D6331)) to assess correct detection of a sample with a known microbial distribution. Finally, reads were demultiplexed, filtered, and adapter sequences and control signals removed.

Supplementary Table 2: Sequencing data summary

|  | IMPACT2-NL | MIND-Set | VHIR | NeuroIMAGE |
| --- | --- | --- | --- | --- |
| **Quality** | | | | |
| average Phred score across all basepairs (forward and reverse) in Median percentile | 37 | 37 | 35.62 | 33.69 |
| **Denoising stats** | | | | |
| Input sequences | 776796.6 | 762115.9 | 75640.5 | 91257.3 |
| Filtered sequences | 693940.1 | 672774.9 | 48995 | 61604.4 |
| Percentage of input passed filter | 89.33 % | 88.28 % | 64.77 % | 67.50 % |
| Denoised sequences | 690254 | 667342.3 | 47970.7 | 56613.5 |
| Merged sequences | 678190 | 652028.4 | 43971.0 | 38715.7 |
| Percentage of input merged | 87.30 % | 85.55 % | 58.13 % | 42.42 % |
| Non-chimeric sequences | 633693.7 | 585221.7 | 39889.2 | 16667.1 |
| Percentage of input non-chimeric | 81.58 % | 76.78 % | 52.74 % | 18.26 % |
| **ASV level** | | | | |
| ASV count | 14408 | 21699 | 6535 | 20459 |
| Mean sequence length | 238.72 | 231.31 | 506.38 | 362.7 |
| Mean frequency per sample | 641512.5 | 601319.9 | 41252.5 | 19016 |
| Median frequency per feature | 23 | 12 | 58 | 50 |
| **Resulting datasets** | | | | |
| Bacteria count | 10422 | 14000 | 6466 | 11406 |
| Genera count | 613 | 564 | 334 | 234 |
| Prevalent genera count (10%) | 234 | 226 | 159 | 116 |

*Summary of sequencing data before and after each preprocessing step: denoising, clustering, genus aggregation and prevalence filtering.*

2.2 Rarefaction curves

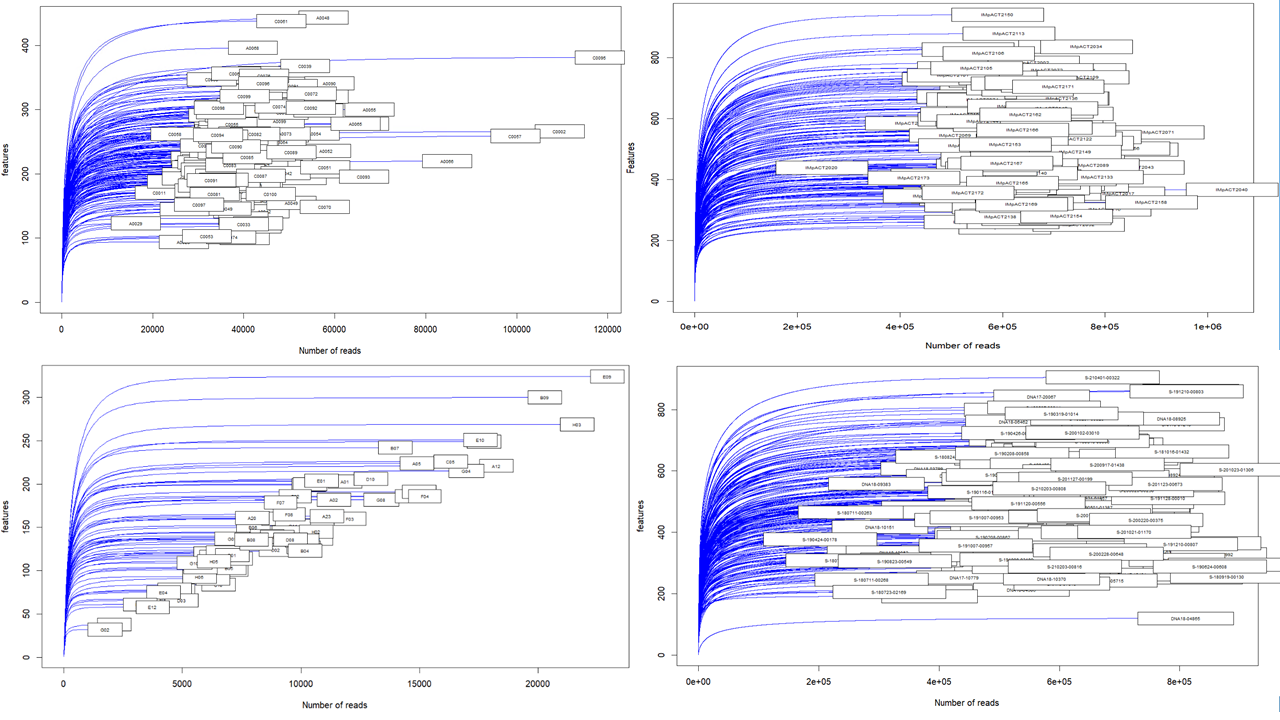

*Supplementary Figure 2. Rarefaction curve of each study (IMpACT2-NL top left, Mental-Cat top right, NeuroIMAGE bottom left, MIND-Set bottom right).*

3 Alpha diversity

3.1 Meta-analyses results of alpha diversity

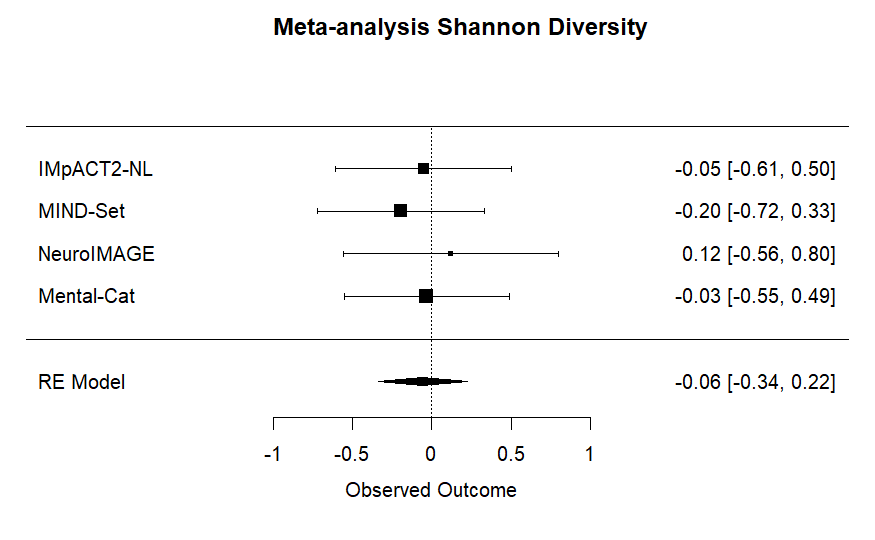

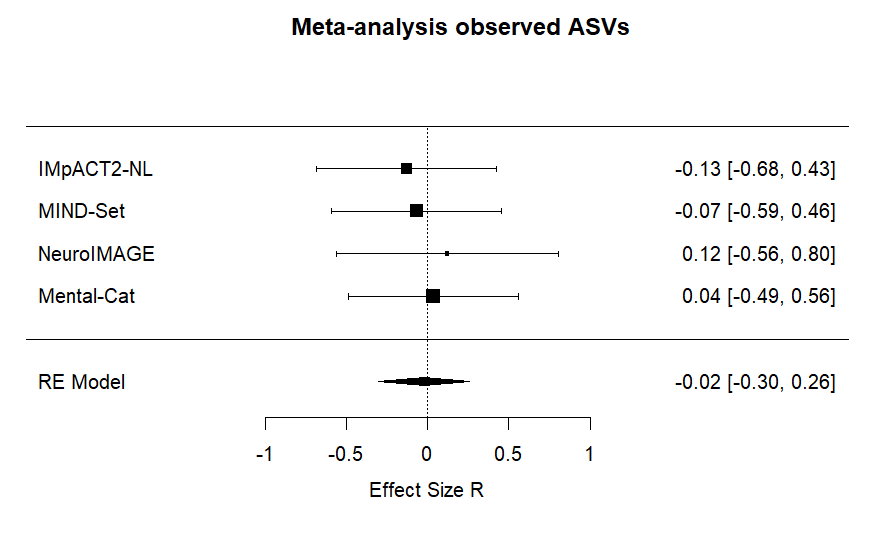

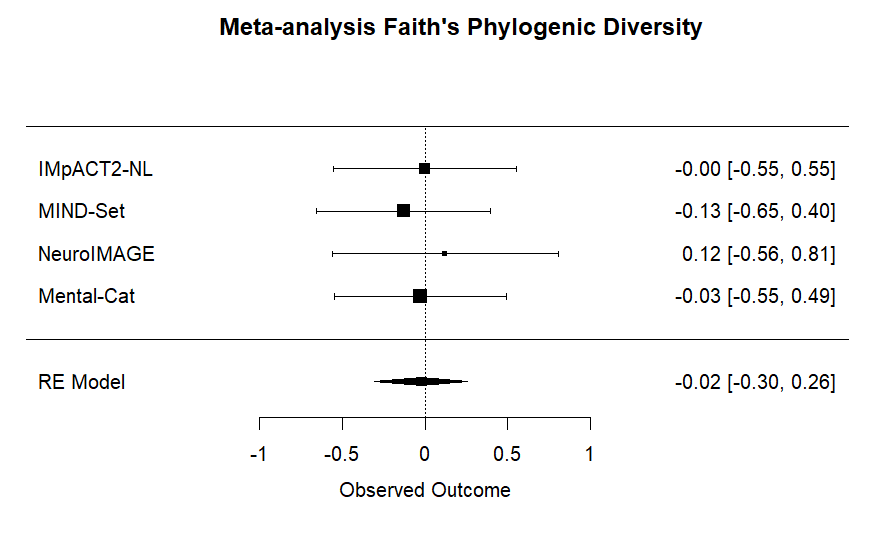

*Supplementary Figure 3. Forest plots of the alpha diversity meta-analyses of observed ASVs (top left), Shannon index (top right), and phylogenic distance (bottom).*

Supplementary Table 3: Results of the alpha diversity meta-analyses

|  | **Observed ASVs** | **Shannon index** | **Phylogenic distance** |
| --- | --- | --- | --- |
| Estimate | -0.02 | -0.06 | -0.02 |
| Standard Error | 0.14 | 0.14 | 0.14 |
| z-value | -0.15 | -0.41 | -0.17 |
| p-value | 0.87 | 0.68 | 0.86 |
| CI lower | -0.30 | -0.33 | -0.30 |
| CI upper | 0.21 | 0.22 | 0.25 |
| p-value heterogeneity | 0.94 | 0.91 | 0.95 |
| Q-value heterogeneity | 0.39 | 0.54 | 0.33 |

3.2 Individual study results of alpha diversity
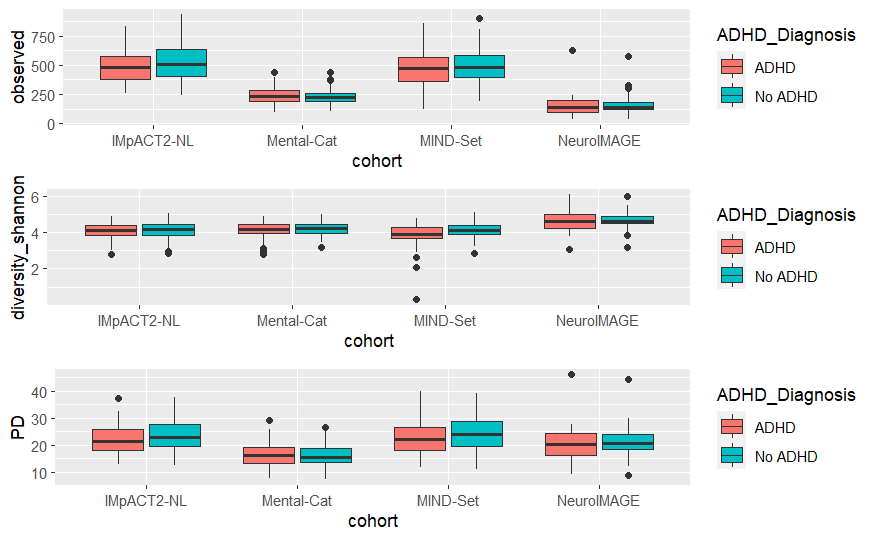

*Supplementary Figure 4. Alpha Diversity per study of observed ASVs (top), Shannon index (middle) and phylogenic diversity (bottom). Participants with ADHD are marked in red, without ADHD are marked in blue.*

Supplementary Table 4: Individual study results of alpha diversity

|  |  | **Observed ASV** | | | **Shannon index** | | | **Phylogenic Diversity** | | |
| --- | --- | --- | --- | --- | --- | --- | --- | --- | --- | --- |
| Cohort |  | ADHD | age | gender | ADHD | age | gender | ADHD | age | gender |
| **IMpACT2** | Estimate | -37.92 | 6.26 | -50.86 | -0.05 | 0.01 | -0.22 | -1.42 | 0.18 | -1.72 |
|  | Std.Error | 23.36 | 1.00 | 23.44 | 0.07 | 0.00 | 0.07 | 0.79 | 0.03 | 0.79 |
|  | T value | -1.62 | 6.27 | -2.17 | -0.66 | 4.47 | -3.02 | -1.80 | 5.45 | -2.17 |
|  | P value | 1.07E-01 | 3.51E-09 | 3.16E-02 | 5.12E-01 | 1.50E-05 | 2.99E-03 | 7.33E-02 | 2.01E-07 | 3.13E-02 |
| **MIND-Set** | Estimate | -21.08 | 1.93 | -9.50 | -0.19 | 0.00 | -0.03 | -1.57 | 0.04 | -0.40 |
|  | Std.Error | 22.16 | 0.76 | 22.42 | 0.07 | 0.00 | 0.07 | 0.95 | 0.03 | 0.96 |
|  | T value | -0.95 | 2.54 | -0.42 | -2.76 | 0.35 | -0.49 | -1.65 | 1.32 | -0.42 |
|  | P value | 3.43^E-01^ | 1.20E-02 | 6.72E-01 | 6.38E-03 | 7.26E-01 | 6.24E-01 | 1.01E-01 | 1.88E-01 | 6.78E-01 |
| **NeuroIMAGE** | Estimate | -13.71 | -4.35 | 2.59 | -0.11 | -0.02 | 0.02 | -0.55 | -0.83 | 0.20 |
|  | Std.Error | 15.71 | 15.85 | 2.81 | 0.12 | 0.12 | 0.02 | 1.44 | 1.45 | 0.26 |
|  | T value | -0.87 | -0.27 | 0.92 | -0.90 | -0.18 | 0.84 | -0.38 | -0.57 | 0.79 |
|  | P value | 3.86E-01 | 7.84E-01 | 3.60E-01 | 3.73E-01 | 8.55E-01 | 4.02E-01 | 7.03E-01 | 5.71E-01 | 4.34E-01 |
| **Mental-Cat** | Estimate | 4.37 | -0.95 | 1.79 | -0.02 | -0.01 | 0.01 | 0.00 | -0.70 | 0.10 |
|  | Std.Error | 8.65 | 8.57 | 0.43 | 0.05 | 0.05 | 0.00 | 0.59 | 0.58 | 0.03 |
|  | T value | 0.50 | -0.11 | 4.18 | -0.47 | -0.27 | 3.13 | -0.01 | -1.21 | 3.47 |
|  | P value | 6.14E-01 | 9.12E-01 | 4.37E-05 | 6.41E-01 | 7.91E-01 | 1.99E-03 | 9.95E-01 | 2.29E-01 | 6.30E-04 |

4 Beta diversity

4.1 Individual study results of beta diversity: CAP Plots
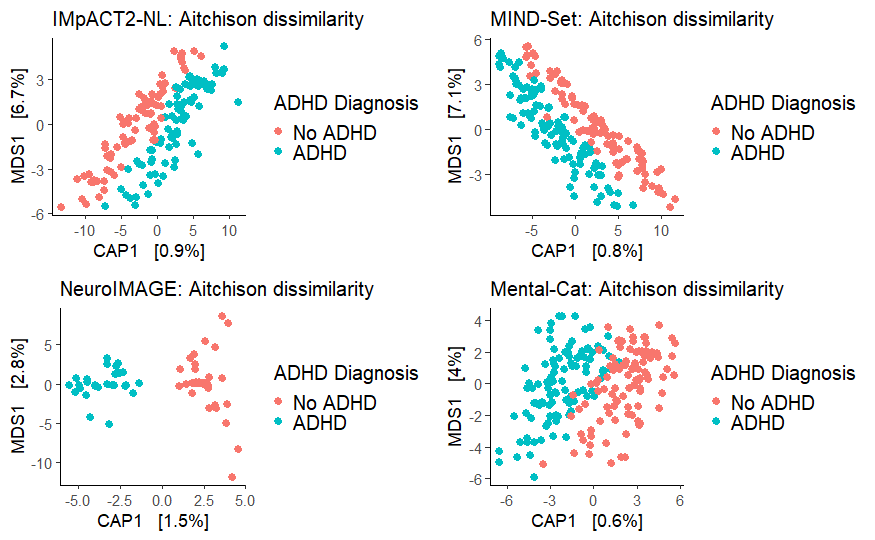

*Supplementary Figure 5. CAP plots supervised for effects of ADHD diagnosis per study.*

Supplementary Table 5. Individual study results of beta diversity permanova

| Cohort | Regressors | Sum Of Squares | R^2^ | F | *P* value |
| --- | --- | --- | --- | --- | --- |
| IMpACT2 | ADHD diagnosis | 12637.89 | 0.01 | 1.41 | 8.00E-03 |
|  | Age | 22960.25 | 0.02 | 2.56 | 1.00E-03 |
|  | Sex | 13255.37 | 0.01 | 1.48 | 3.00E-03 |
| MIND-Set | ADHD diagnosis | 13463.10 | 0.01 | 1.50 | 5.00E-03 |
|  | Age | 17776.96 | 0.01 | 1.99 | 1.00E-03 |
|  | Sex | 10074.13 | 0.01 | 1.13 | 1.19E-01 |
| NeuroIMAGE | ADHD diagnosis | 5176.29 | 0.01 | 0.95 | 9.61E-01 |
|  | Age | 5612.08 | 0.02 | 1.03 | 2.80E-01 |
|  | Sex | 5456.44 | 0.02 | 1.00 | 4.61E-01 |
| Mental-Cat | ADHD diagnosis | 5282.24 | 0.01 | 1.19 | 1.10E-02 |
|  | Age | 7021.62 | 0.01 | 1.45 | 2.00E-03 |
|  | Sex | 6565.05 | 0.01 | 1.36 | 4.00E-03 |

5 Feature selection

Supplementary Table 6. Summary of the selected genera across studies

| Genus | Selection probability |
| --- | --- |
| *Tyzzerella* | 0.4992 |
| *Eubacterium_eligens_group* | 0.4581 |
| *Romboutsia* | 0.3682 |
| *RF39* | 0.3384 |
| *Victivallis* | 0.3398 |
| *Lachnospiraceae UCG 001* | 0.2562 |
| *Ruminococcus torques group* | 0.2222 |
| *Eubacterium ventriosum group* | 0.2059 |
| *Ruminococcus* | 0.1962 |
| *Sutterella* | 0.1849 |
| *Eubacterium xylanophilum group* | 0.1523 |
| *Eisenbergiella* | 0.1303 |
| *Erysipelatoclostridium* | 0.1294 |
| *Dialister* | 0.1289 |
| *Clostridia UCG 014* | 0.1186 |
| *Eubacterium ruminantium group* | 0.1123 |
| *Coprococcus* | 0.1098 |
| *Desulfovibrio* | 0.1059 |
| *Lactococcus* | 0.1052 |
| *Muribaculaceae* | 0.1041 |

*Feature selection results over all studies. Selection probability of all selected genera are shown across studies.*

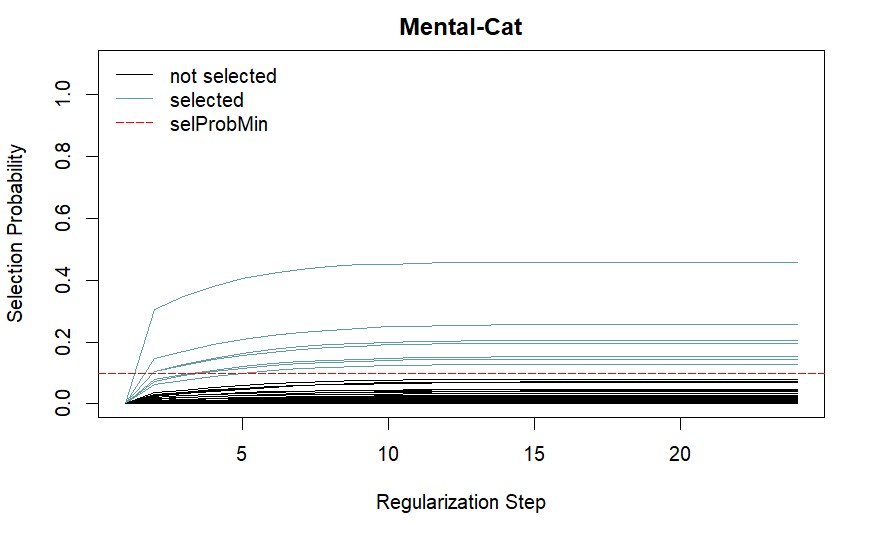

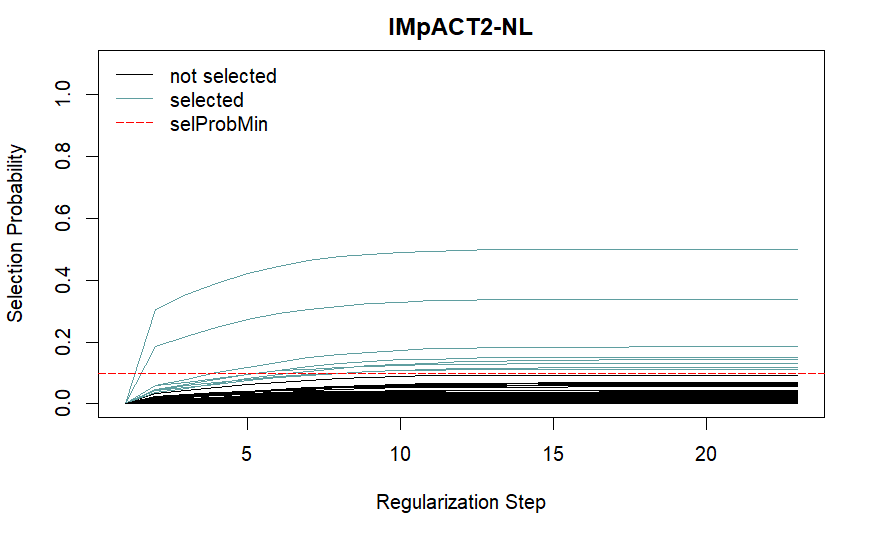

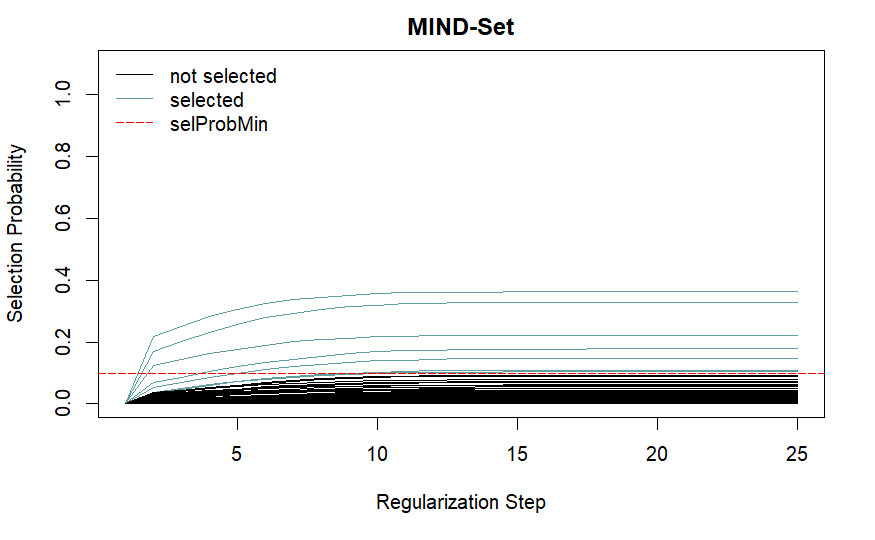

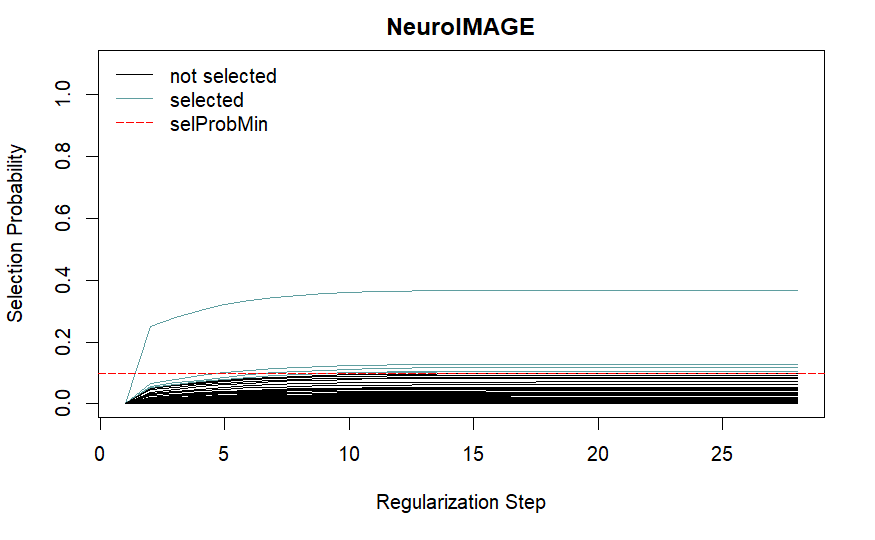
 *Supplementary Figure 6. Stability path plots of the feature selection for Mental-Cat (top-left),IMpACT2-NL (top-right), MIND-Set (bottom-left) and NeuroIMAGE (bottom-right). Randomized Lasso stability selection starts with a probability of 0 for all genera. With each regularization step (penalized regression on a different subsample), the selection probability is updated. The selection probability threshold of 10% (marked in red) was chosen in order to pick up signals deviating visibly from the center of mass around 0 across all four studies. Stability paths of genera with selection probability of 10% or higher are marked in blue. Unselected genera are marked in black.*

6 Differential abundance analysis

6.1 Logistic regression Meta-analyses

Supplementary Table 7. Results of the logistic regression meta-analyses

| Genus | uncorrected p-value Logistic Regression | fdr corrected p-value Logistic Regression | Effect Size R Logistic Regression | Standard Error of R Logistic Regression | I2 Logistic Regression | pvalue test for heterogeneity Logistic Regression | Q value test for heterogeneity Logistic Regression |
| --- | --- | --- | --- | --- | --- | --- | --- |
| *Eubacterium xylanophilum group* | 3.31E-04 | 6.95E-03 | -0.12 | 0.03 | 0.04 | 3.04E-01 | 3.63 |
| *Eisenbergiella* | 1.91E-03 | 2.00E-02 | 0.14 | 0.05 | 0.00 | 4.91E-01 | 2.41 |
| *Clostridia UCG 014* | 1.05E-02 | 4.42E-02 | -0.07 | 0.03 | 0.23 | 2.15E-01 | 4.47 |
| *Eubacterium ruminantium group* | 9.68E-03 | 4.42E-02 | -0.07 | 0.03 | 0.01 | 1.77E-01 | 4.93 |
| *Ruminococcus torques group* | 1.05E-02 | 4.42E-02 | 0.17 | 0.07 | 7.54 | 2.64E-01 | 3.98 |
| *Eubacterium brachy group* | 3.22E-02 | 1.03E-01 | 0.11 | 0.05 | 0.00 | 6.83E-01 | 1.50 |
| *RF39* | 3.44E-02 | 1.03E-01 | -0.10 | 0.05 | 51.08 | 1.45E-01 | 5.40 |
| *Eubacterium ventriosum group* | 5.69E-02 | 1.39E-01 | -0.15 | 0.08 | 56.72 | 7.66E-02 | 6.86 |
| *Lachnospiraceae UCG 001* | 5.95E-02 | 1.39E-01 | -0.08 | 0.04 | 33.50 | 2.38E-01 | 4.23 |
| *Ruminococcus* | 7.41E-02 | 1.41E-01 | -0.09 | 0.05 | 27.30 | 3.03E-01 | 3.64 |
| *Victivallis* | 6.87E-02 | 1.41E-01 | -0.13 | 0.07 | 66.43 | 3.14E-02 | 8.85 |
| *Tyzzerella* | 8.49E-02 | 1.49E-01 | 0.09 | 0.05 | 59.95 | 6.10E-02 | 7.37 |
| *Sutterella* | 1.51E-01 | 2.44E-01 | -0.10 | 0.07 | 73.79 | 1.32E-02 | 10.74 |
| *Eubacterium eligens group* | 1.84E-01 | 2.76E-01 | -0.10 | 0.07 | 70.75 | 2.08E-02 | 9.76 |
| *Coprococcus* | 2.55E-01 | 3.58E-01 | -0.12 | 0.10 | 59.90 | 6.42E-02 | 7.26 |
| *Muribaculaceae* | 3.19E-01 | 4.19E-01 | -0.03 | 0.03 | 41.84 | 2.12E-01 | 4.51 |
| *Erysipelatoclostridium* | 3.67E-01 | 4.31E-01 | 0.07 | 0.07 | 65.97 | 4.67E-02 | 7.97 |
| *Romboutsia* | 3.69E-01 | 4.31E-01 | 0.04 | 0.05 | 30.29 | 2.31E-01 | 4.30 |
| *Lactococcus* | 7.06E-01 | 7.80E-01 | 0.02 | 0.04 | 0.07 | 2.38E-01 | 4.23 |
| *Dialister* | 8.18E-01 | 8.59E-01 | 0.01 | 0.05 | 73.97 | 1.49E-02 | 10.48 |
| *Desulfovibrio* | 9.31E-01 | 9.31E-01 | 0.00 | 0.03 | 0.01 | 6.02E-01 | 1.86 |

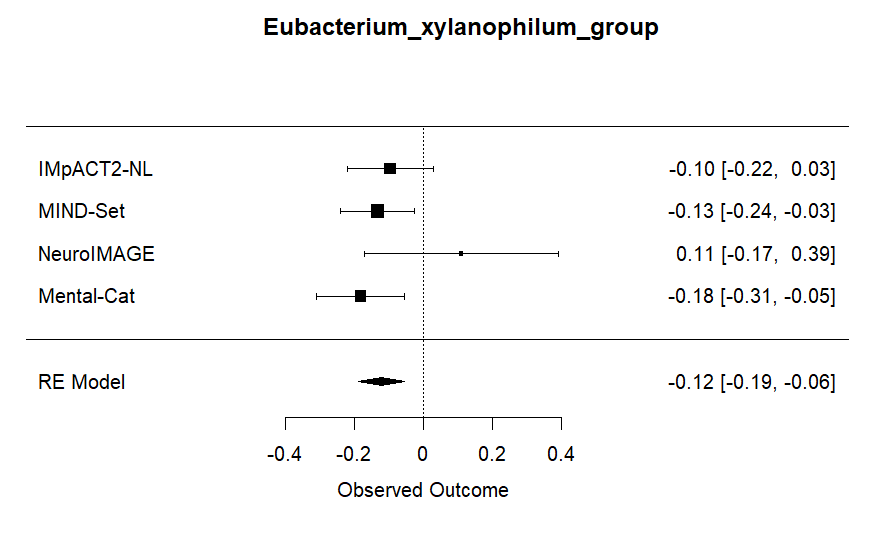

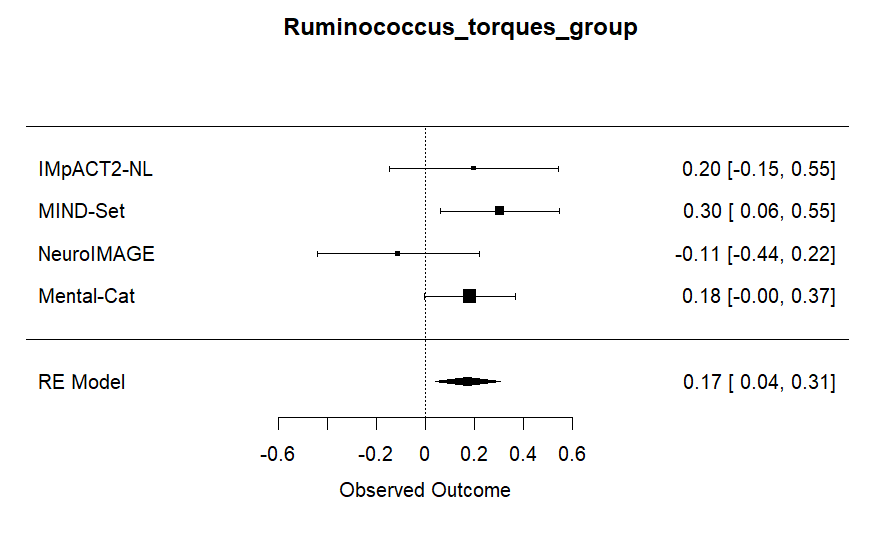

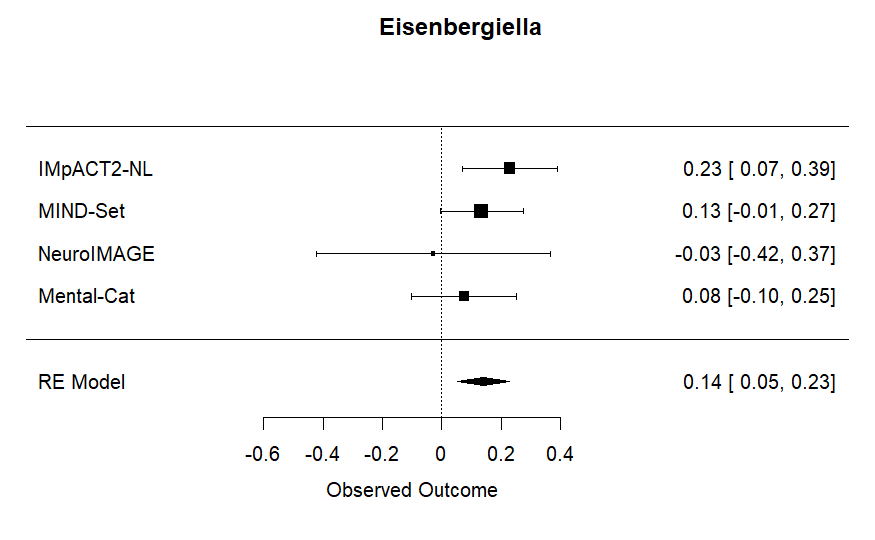

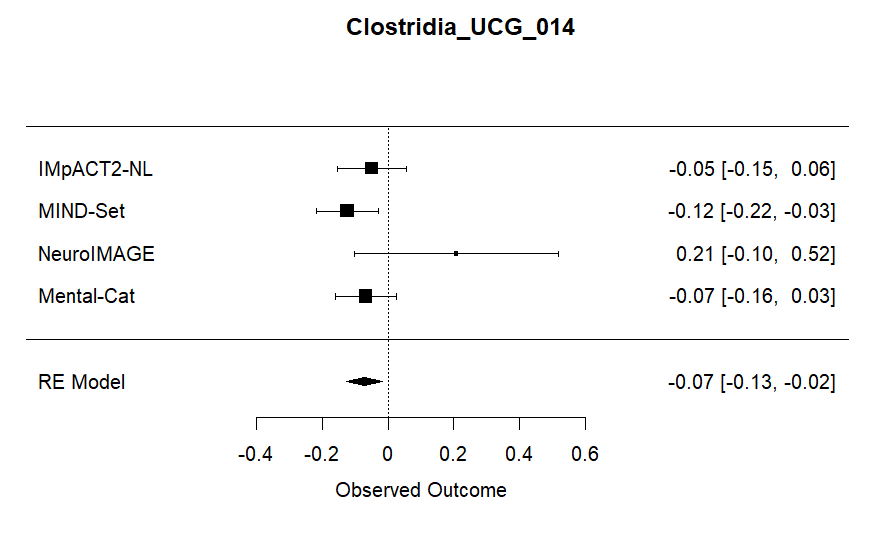

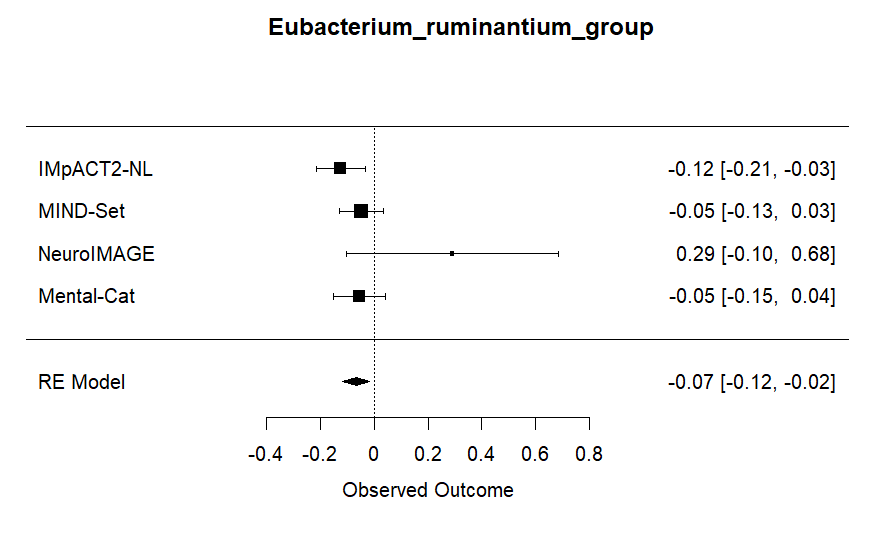

*Supplementary Figure 7. Forest plots of the genera showing significant associations with ADHD diagnosis in the meta-analyses based on logistic regression.*

6.2 ALDEx2 Meta-analyses

Supplementary Table 8. Results of the ALDEx2 meta-analyses

| Genus | uncorrected p-value ALDEx2 | Effect Size R ALDEx2 | Standard Error of R ALDEx2 | I^2^ ALDEx2 | pvalue test for heterogeneity ALDEx2 | Q value test for heterogeneity ALDEx2 | fdr corrected p-value ALDEx2 |
| --- | --- | --- | --- | --- | --- | --- | --- |
| *Eubacterium xylanophilum group* | 1.87E-03 | -0.13 | 0.04 | 0.03 | 1.77E-01 | 4.93 | 1.96E-02 |
| *Ruminococcus torques group* | 1.46E-03 | 0.13 | 0.04 | 0.00 | 8.81E-01 | 0.67 | 1.96E-02 |
| *Eisenbergiella* | 7.86E-03 | 0.11 | 0.04 | 0.00 | 5.13E-01 | 2.30 | 5.50E-02 |
| *Clostridia UCG 014* | 1.23E-02 | 0.11 | 0.04 | 15.27 | 4.05E-01 | 2.91 | 6.48E-02 |
| *Erysipelatoclostridium* | 3.22E-02 | -0.09 | 0.04 | 4.15 | 2.29E-01 | 4.32 | 1.35E-01 |
| *Eubacterium brachy group* | 1.07E-01 | 0.06 | 0.04 | 0.00 | 4.96E-01 | 2.39 | 3.48E-01 |
| *Tyzzerella* | 1.16E-01 | 0.11 | 0.07 | 63.06 | 4.27E-02 | 8.17 | 3.48E-01 |
| *Eubacterium ruminantium group* | 1.50E-01 | -0.08 | 0.06 | 45.24 | 1.22E-01 | 5.80 | 3.51E-01 |
| *Romboutsia* | 1.41E-01 | 0.07 | 0.04 | 15.71 | 3.74E-01 | 3.12 | 3.51E-01 |
| *Ruminococcus* | 1.76E-01 | -0.07 | 0.05 | 33.39 | 1.78E-01 | 4.92 | 3.69E-01 |
| *Lachnospiraceae UCG 001* | 2.11E-01 | -0.07 | 0.06 | 50.65 | 1.02E-01 | 6.21 | 4.03E-01 |
| *RF39* | 2.95E-01 | -0.09 | 0.08 | 75.75 | 9.02E-03 | 11.57 | 5.16E-01 |
| *Victivallis* | 3.73E-01 | -0.07 | 0.08 | 69.98 | 1.94E-02 | 9.90 | 6.03E-01 |
| *Coprococcus* | 4.97E-01 | -0.04 | 0.06 | 54.19 | 8.54E-02 | 6.61 | 6.03E-01 |
| *Eubacterium eligens group* | 5.46E-01 | -0.04 | 0.07 | 68.09 | 2.39E-02 | 9.45 | 6.03E-01 |
| *Eubacterium ventriosum group* | 4.34E-01 | -0.05 | 0.06 | 53.27 | 9.27E-02 | 6.42 | 6.03E-01 |
| *Lactococcus* | 4.67E-01 | 0.03 | 0.04 | 0.00 | 6.72E-01 | 1.55 | 6.03E-01 |
| *Muribaculaceae* | 5.23E-01 | -0.04 | 0.06 | 46.10 | 1.29E-01 | 5.66 | 6.03E-01 |
| *Sutterella* | 4.98E-01 | -0.06 | 0.08 | 75.62 | 7.78E-03 | 11.89 | 6.03E-01 |
| *Dialister* | 7.38E-01 | 0.02 | 0.07 | 65.95 | 2.92E-02 | 9.01 | 7.75E-01 |
| *Desulfovibrio* | 9.17E-01 | 0.00 | 0.04 | 0.00 | 7.41E-01 | 1.25 | 9.17E-01 |

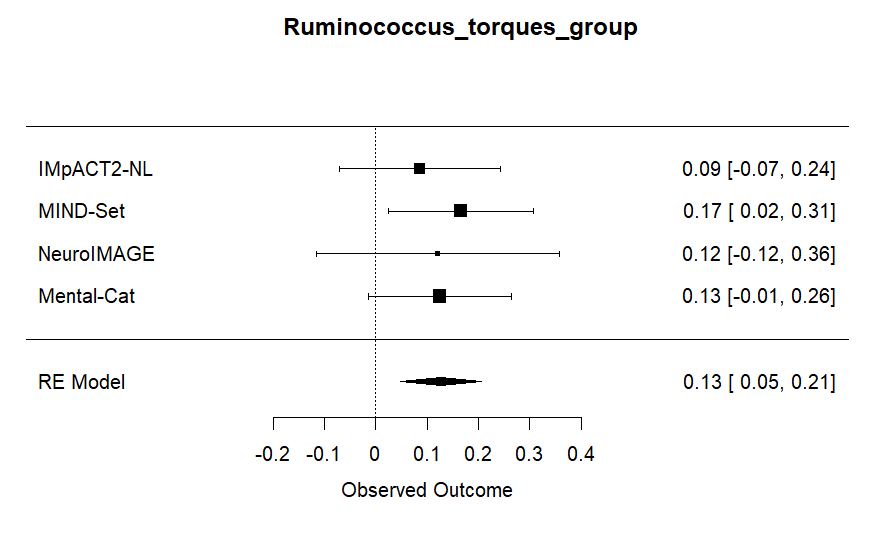

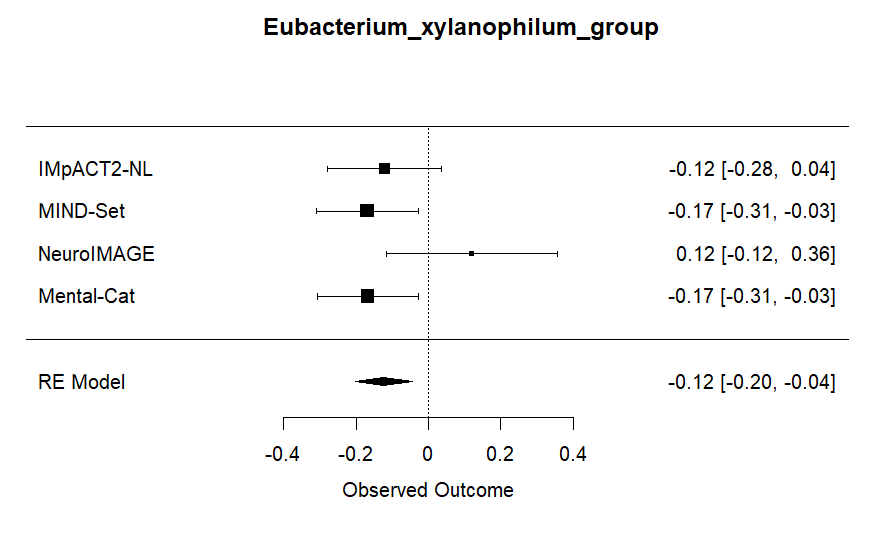

*Supplementary Figure 8.* *Forest plots of the genera showing significant associations with ADHD diagnosis in the meta-analyses based on ALDEx2.*

6.3 Relative Abundance plots

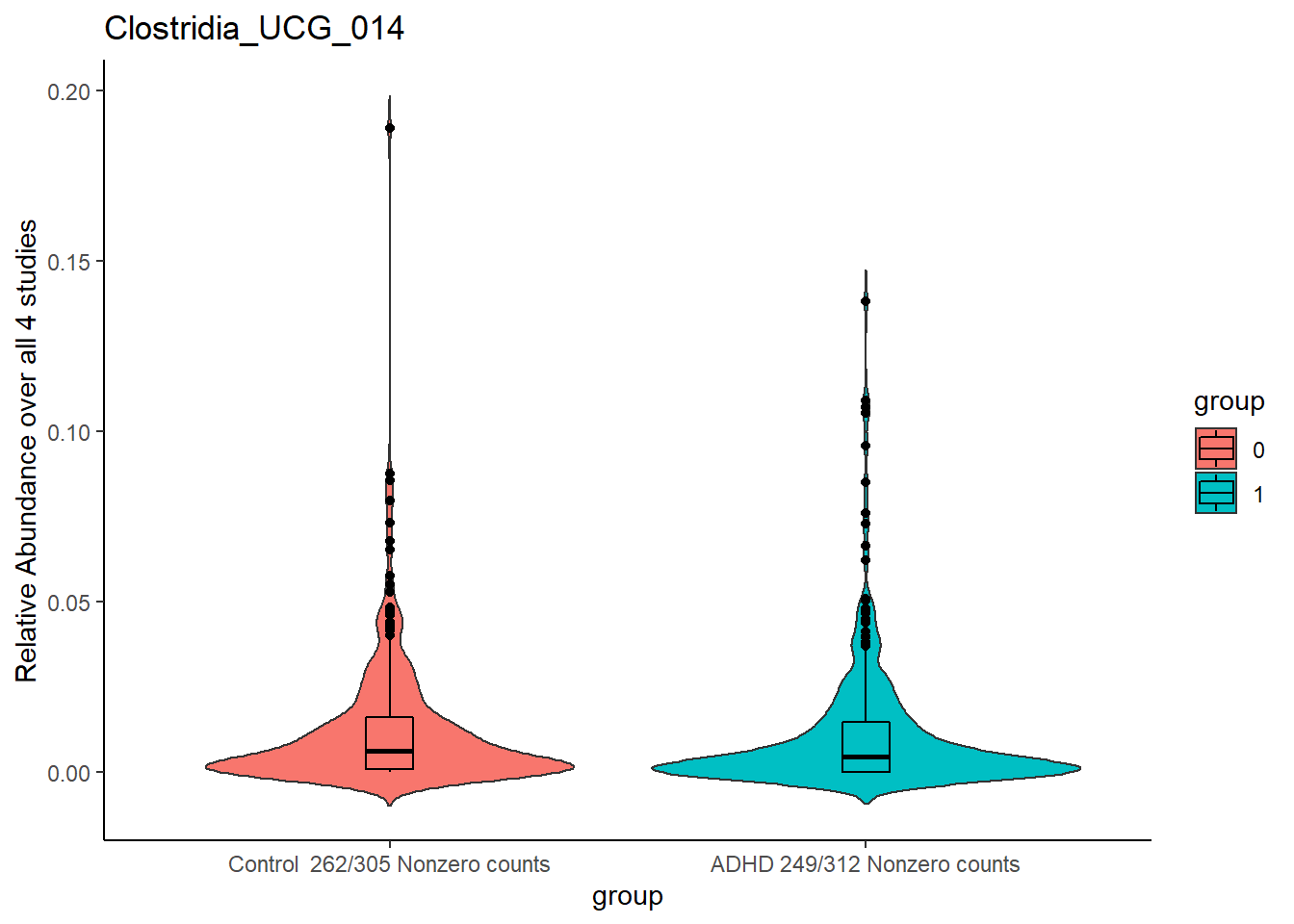

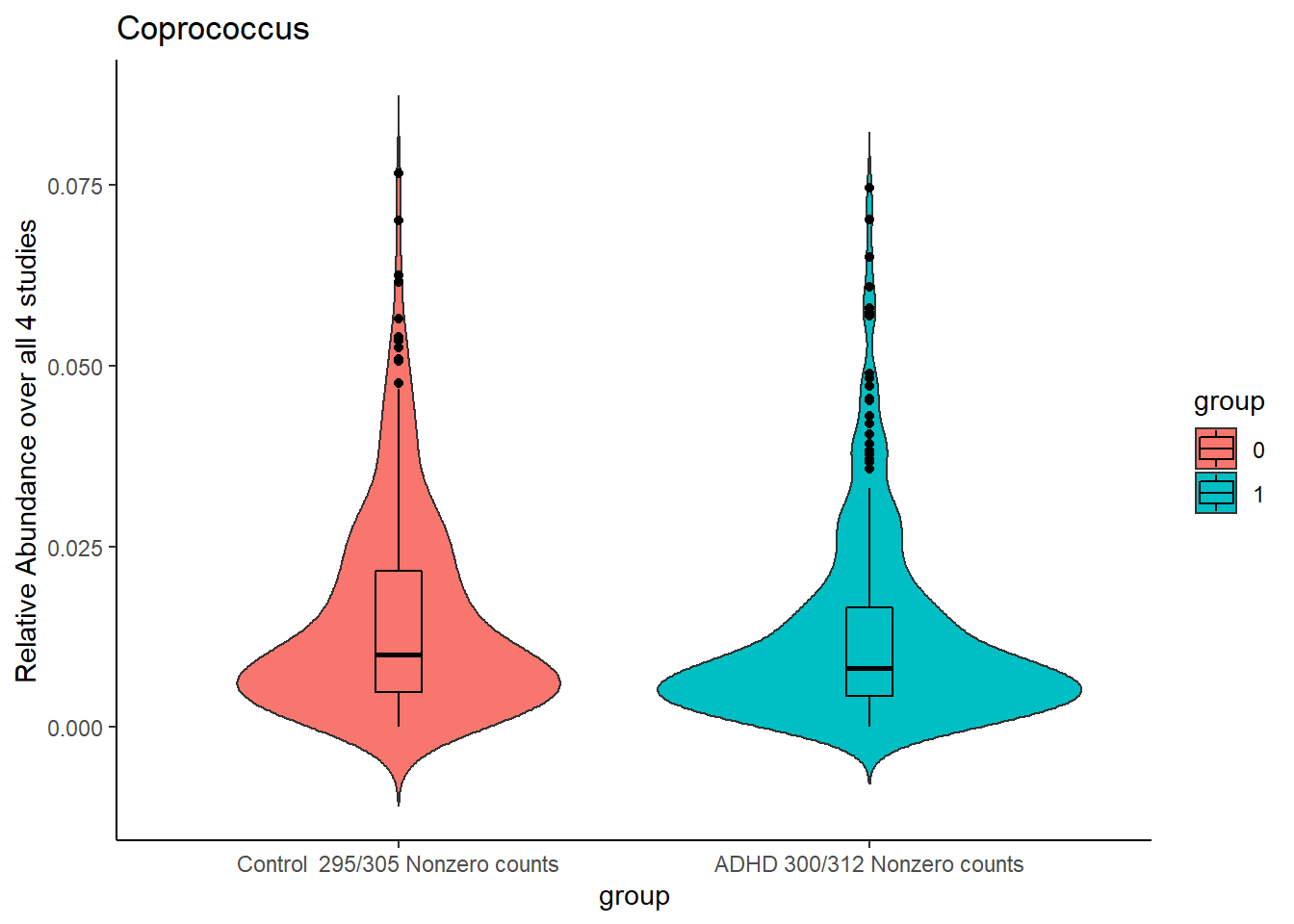

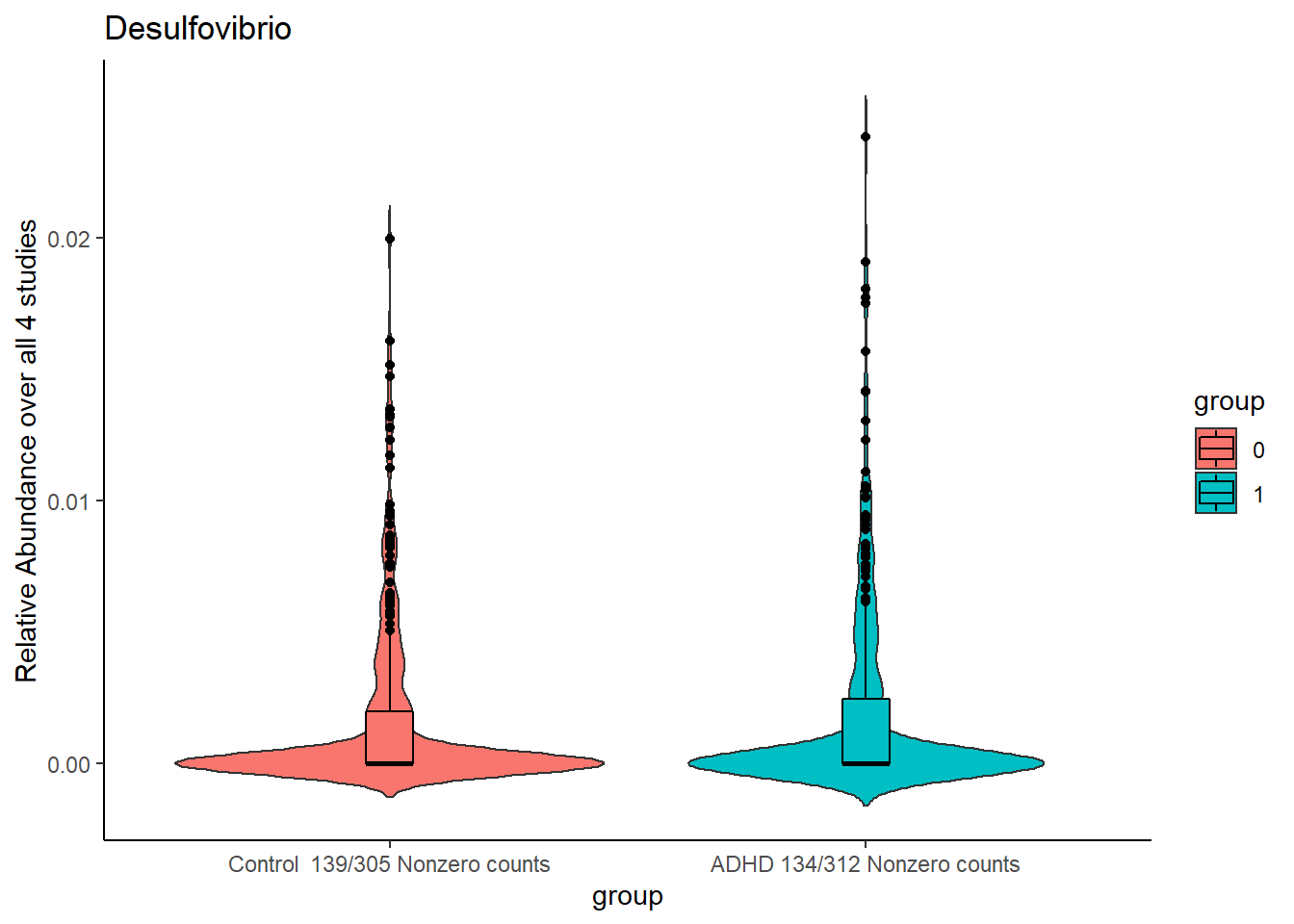

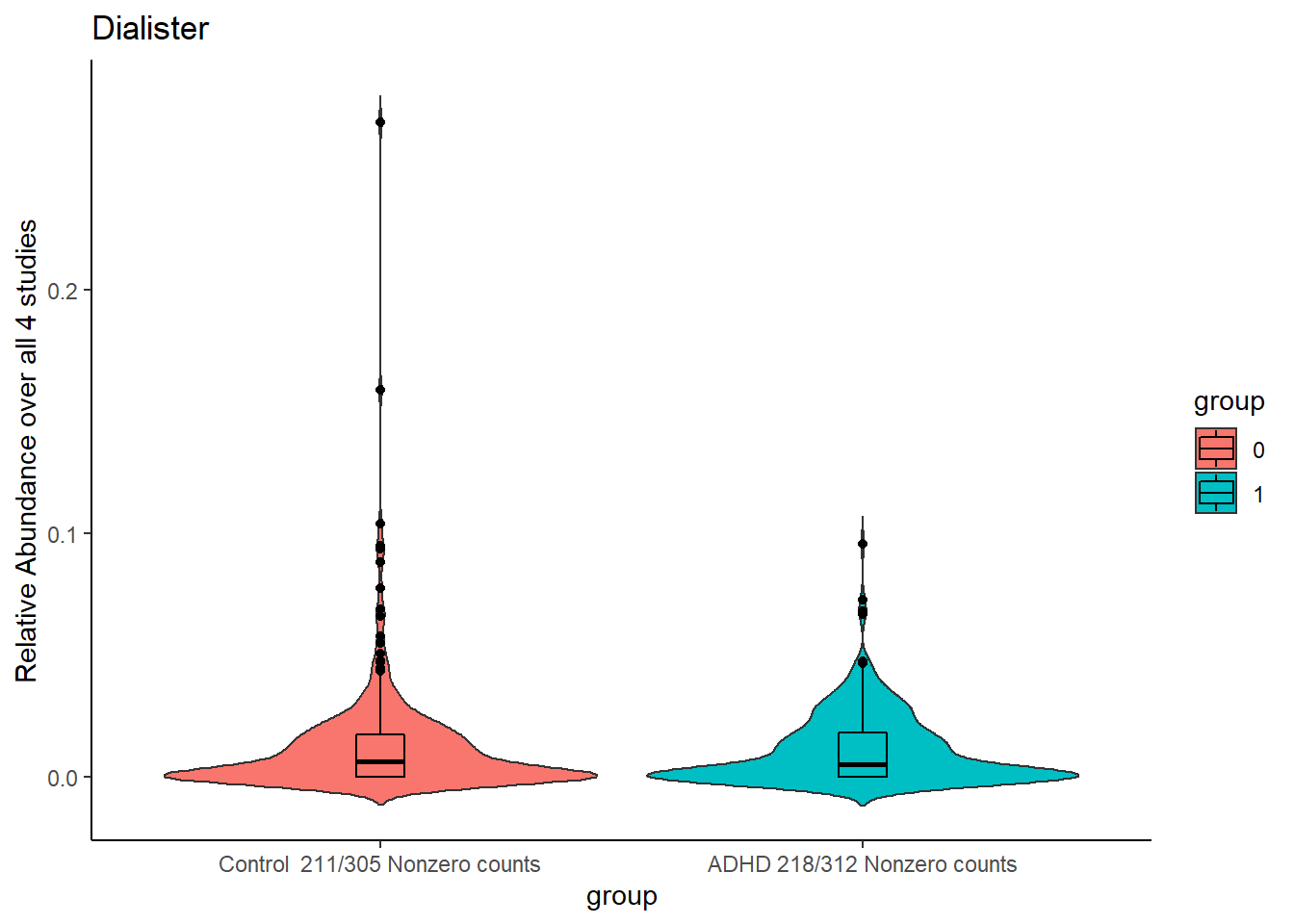

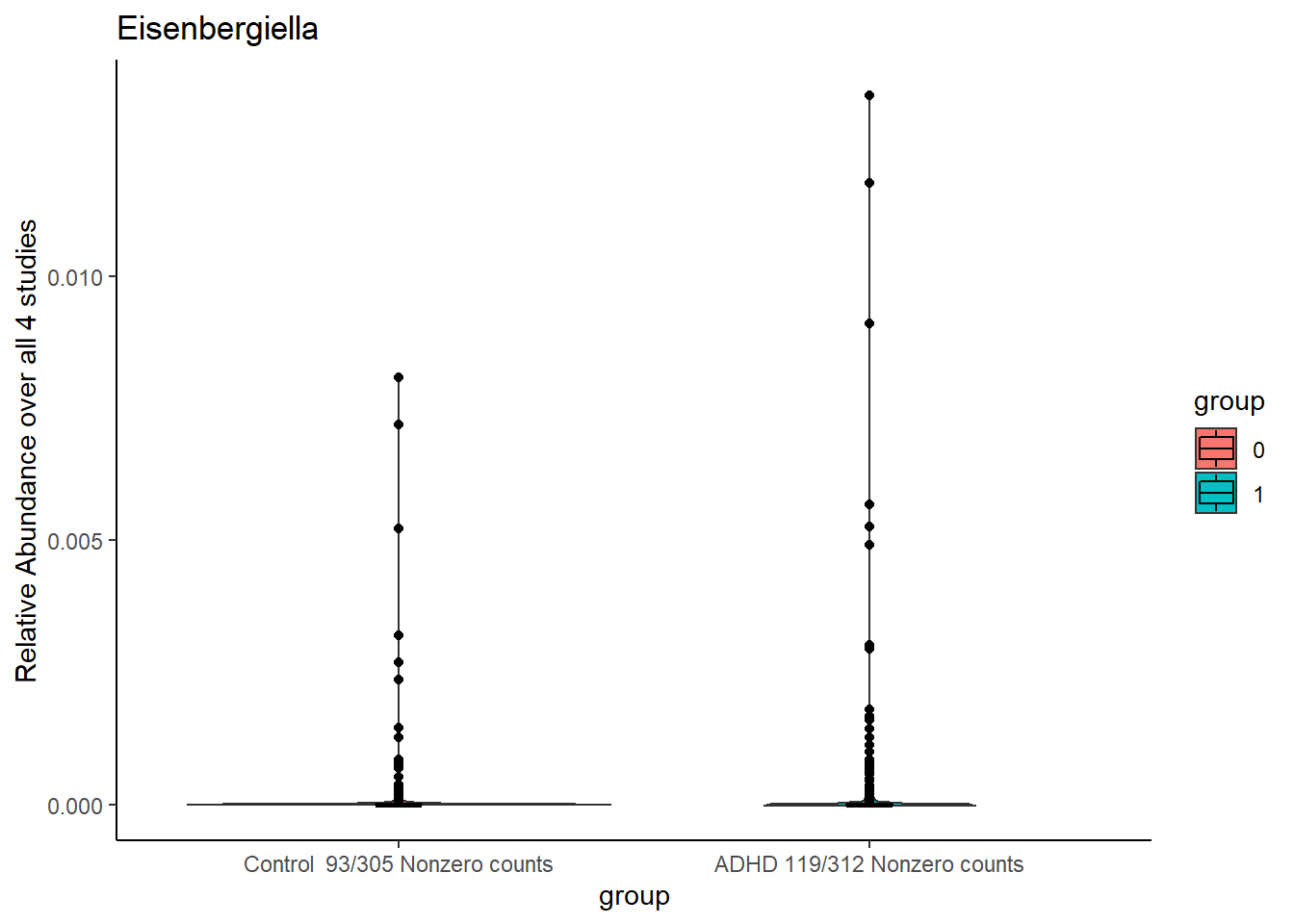

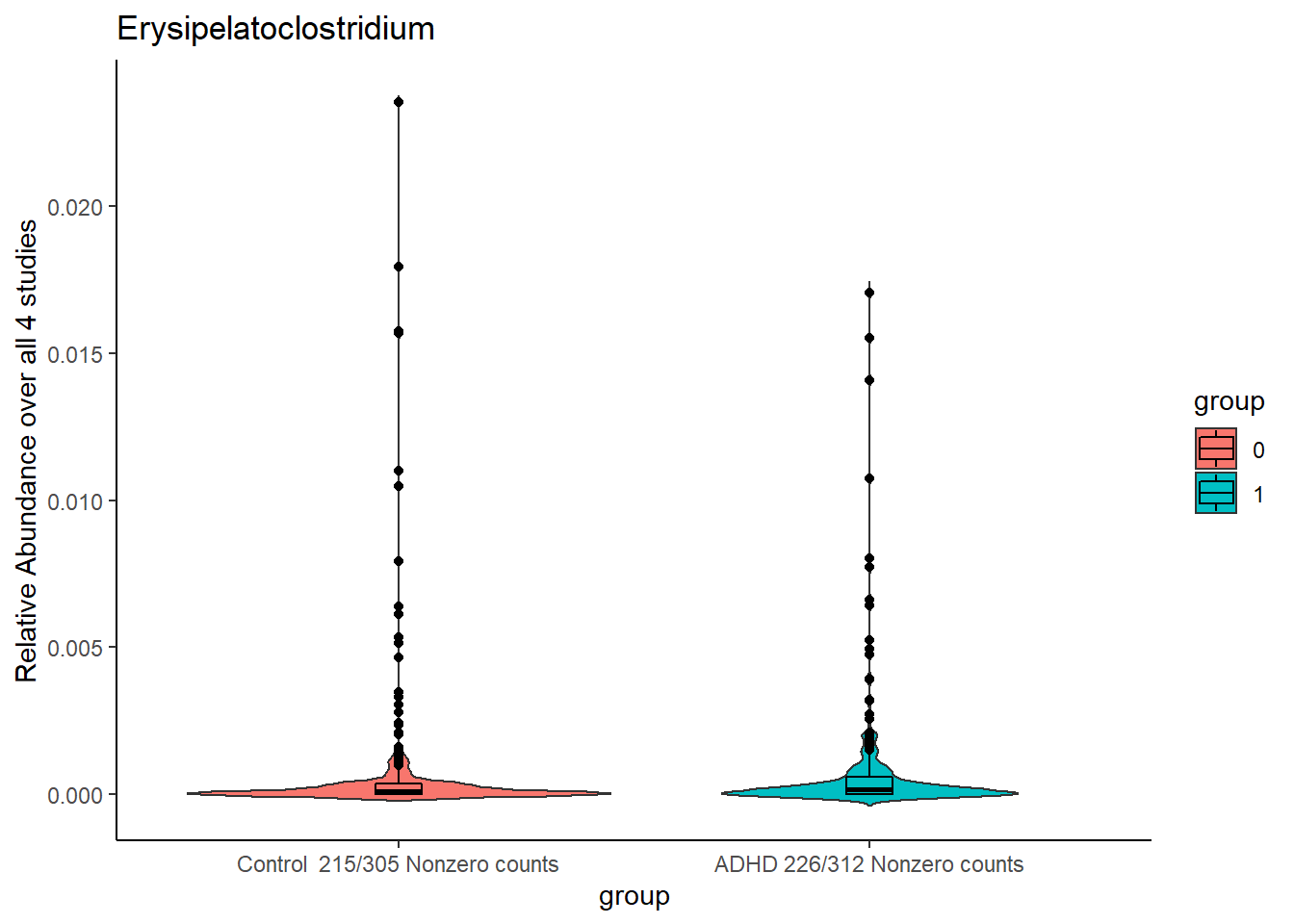

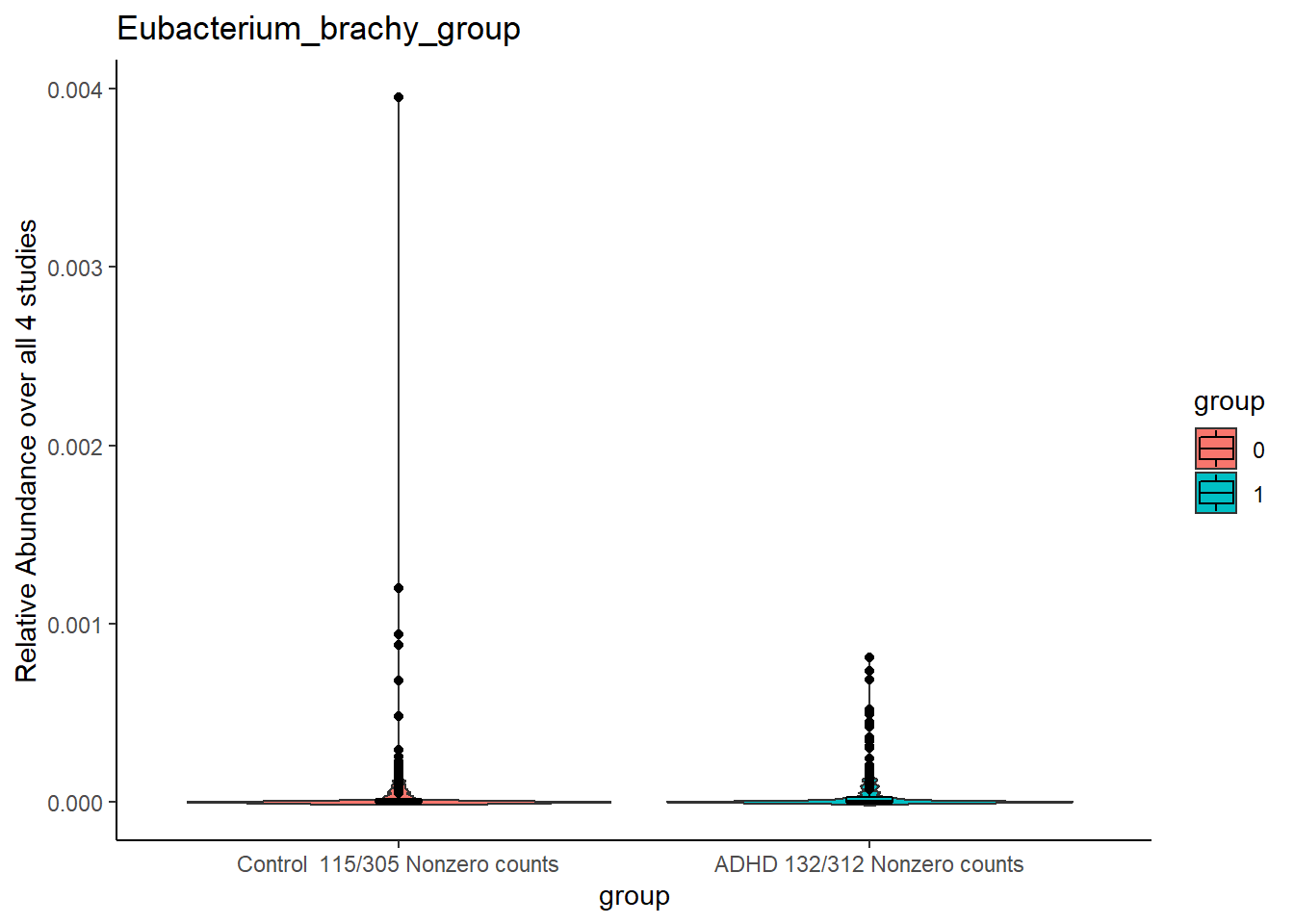

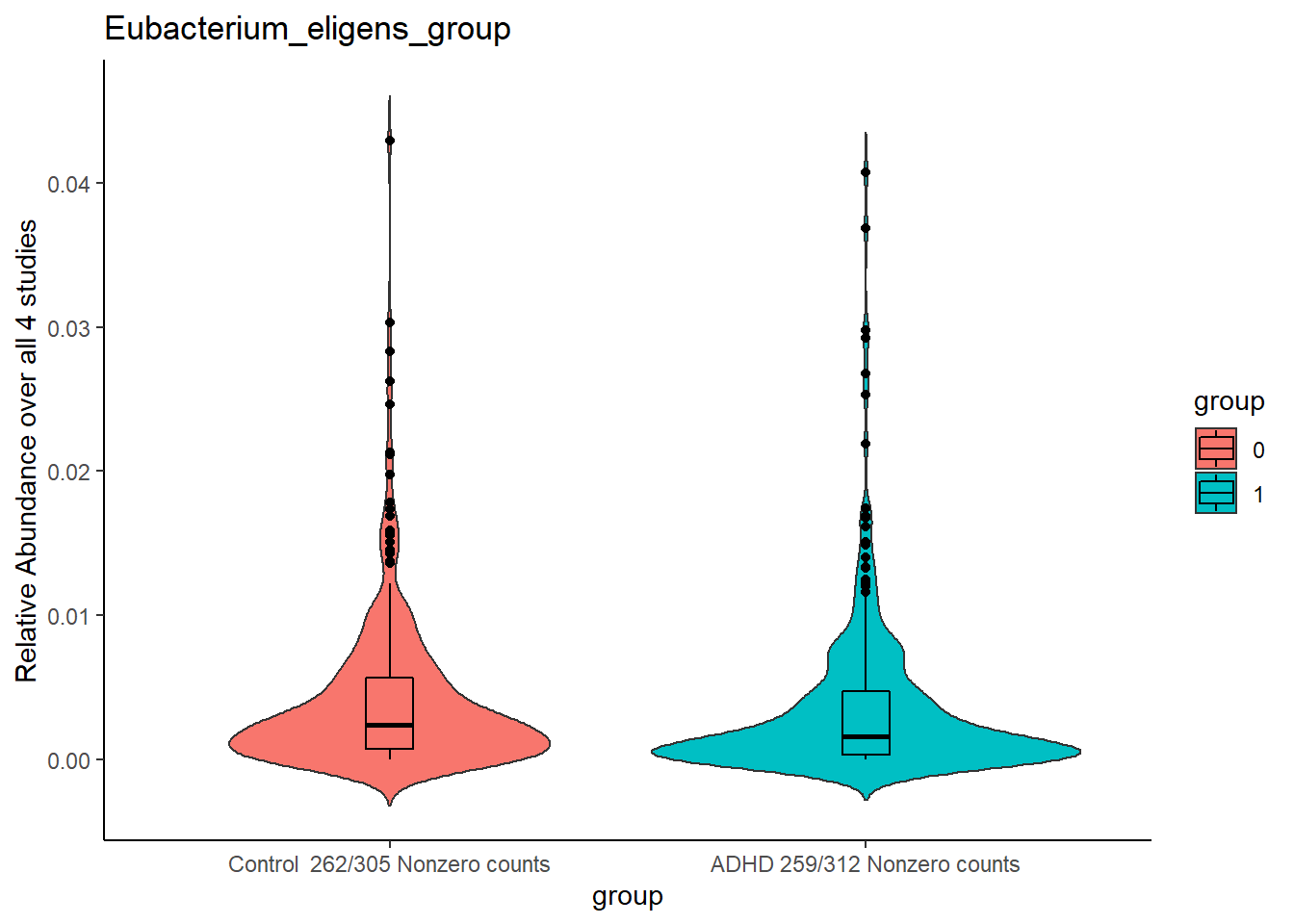

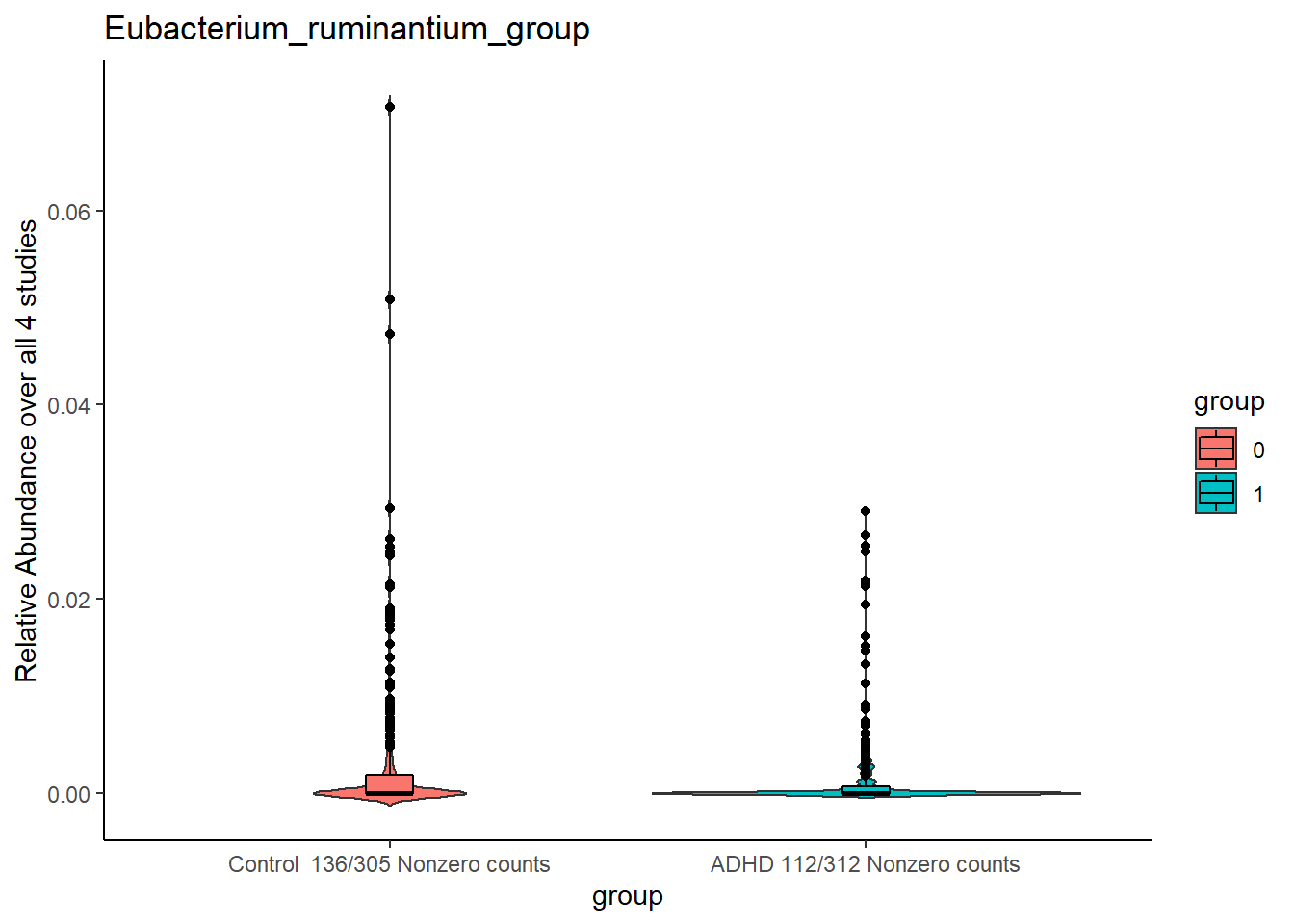

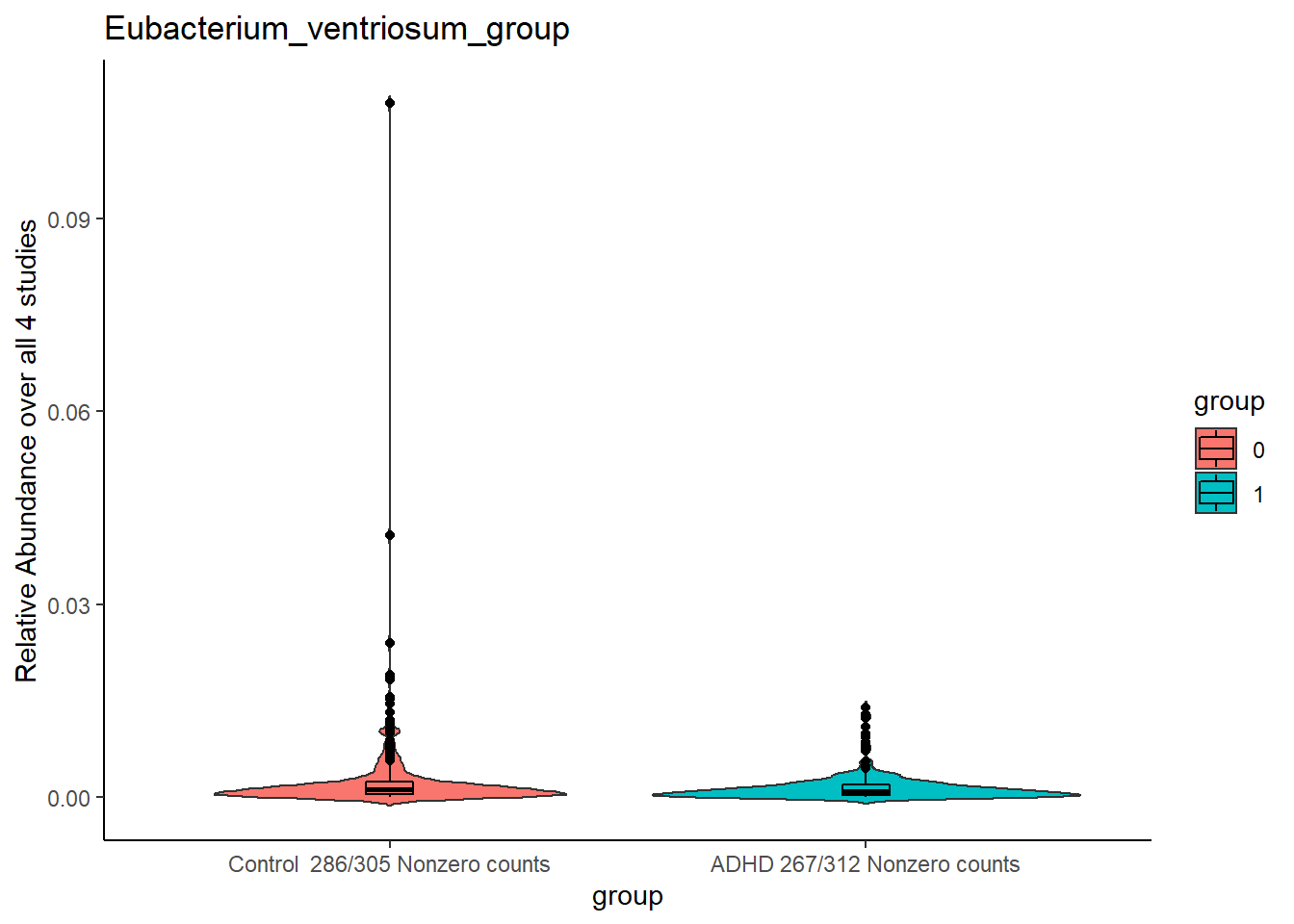

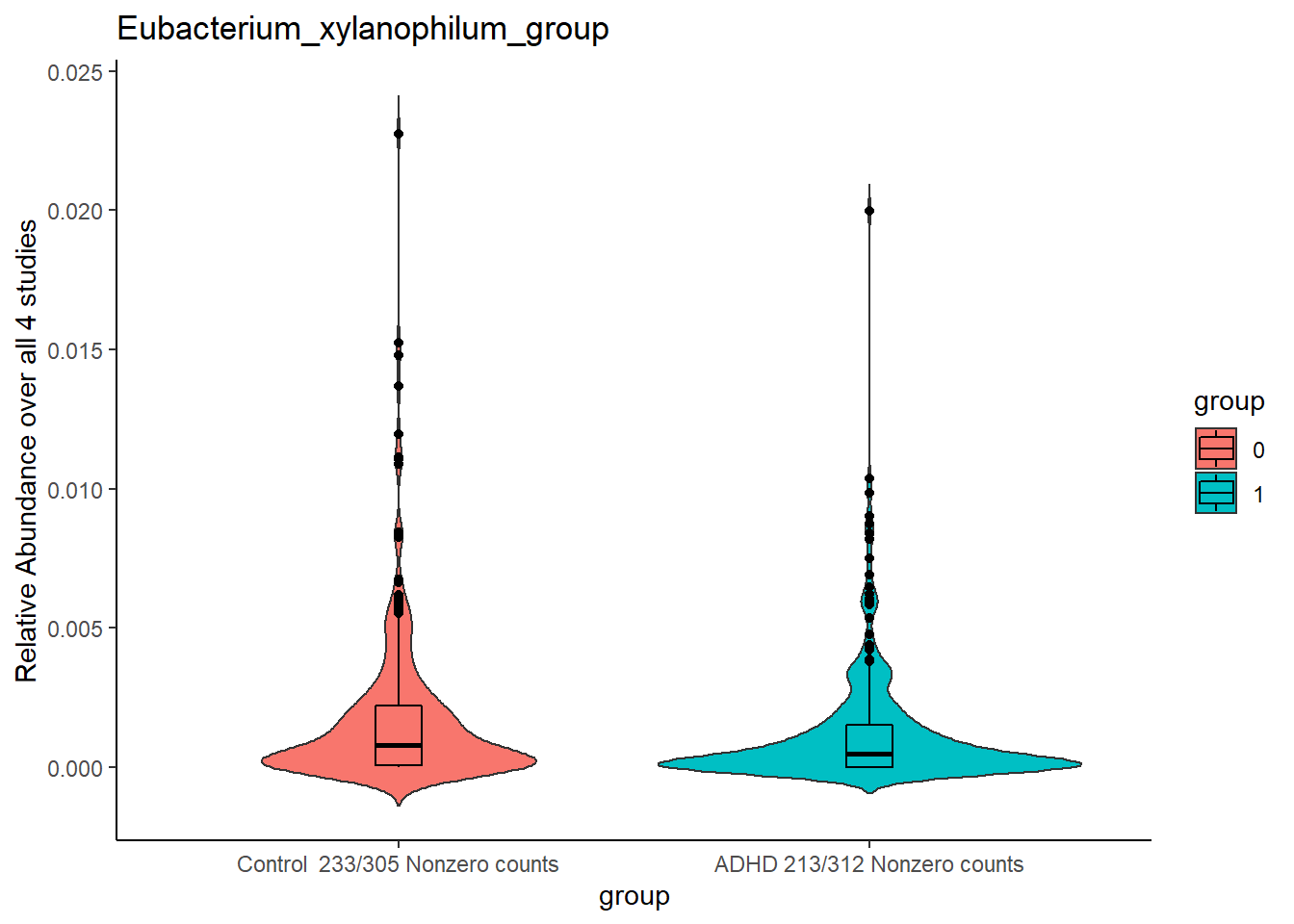

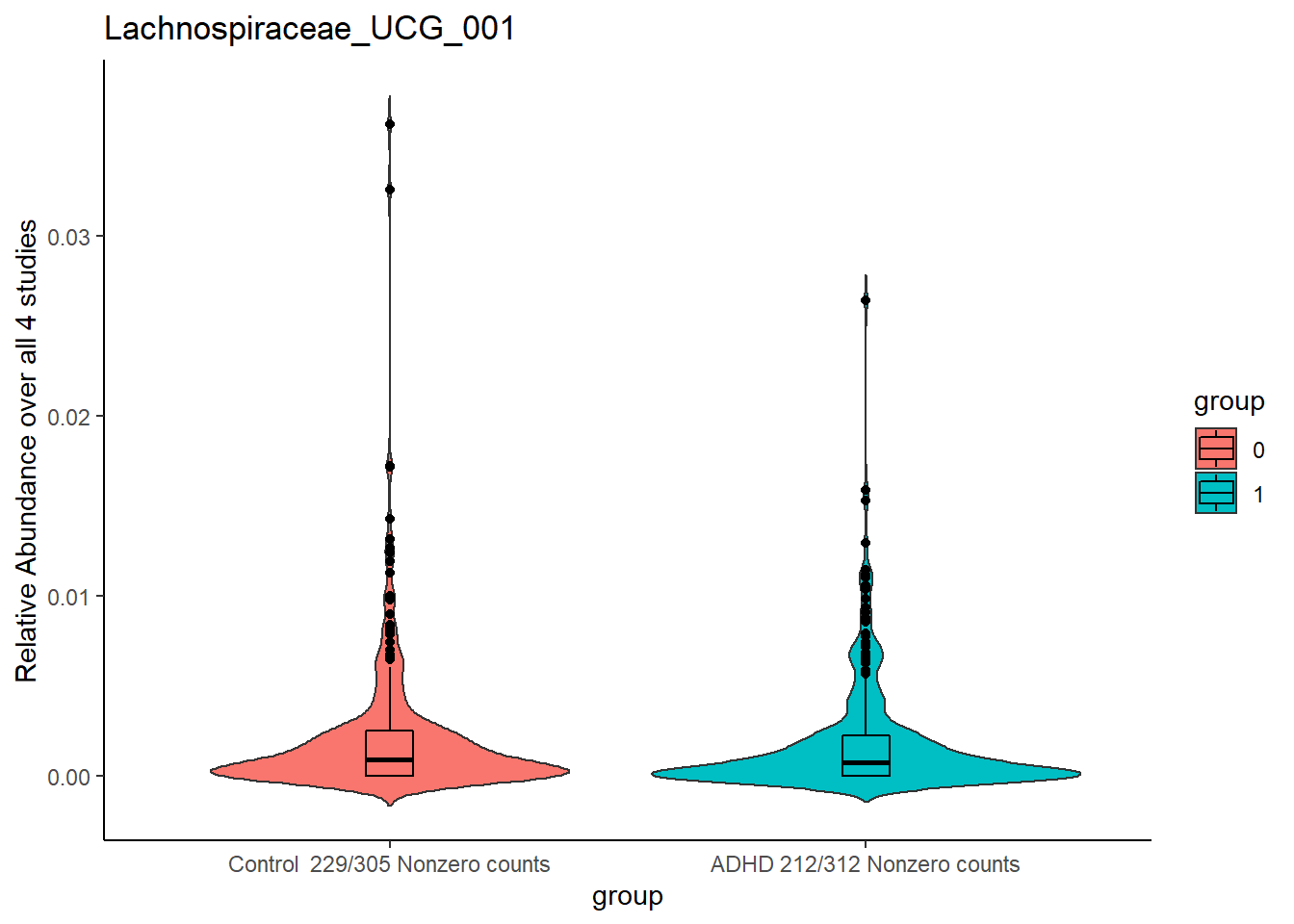

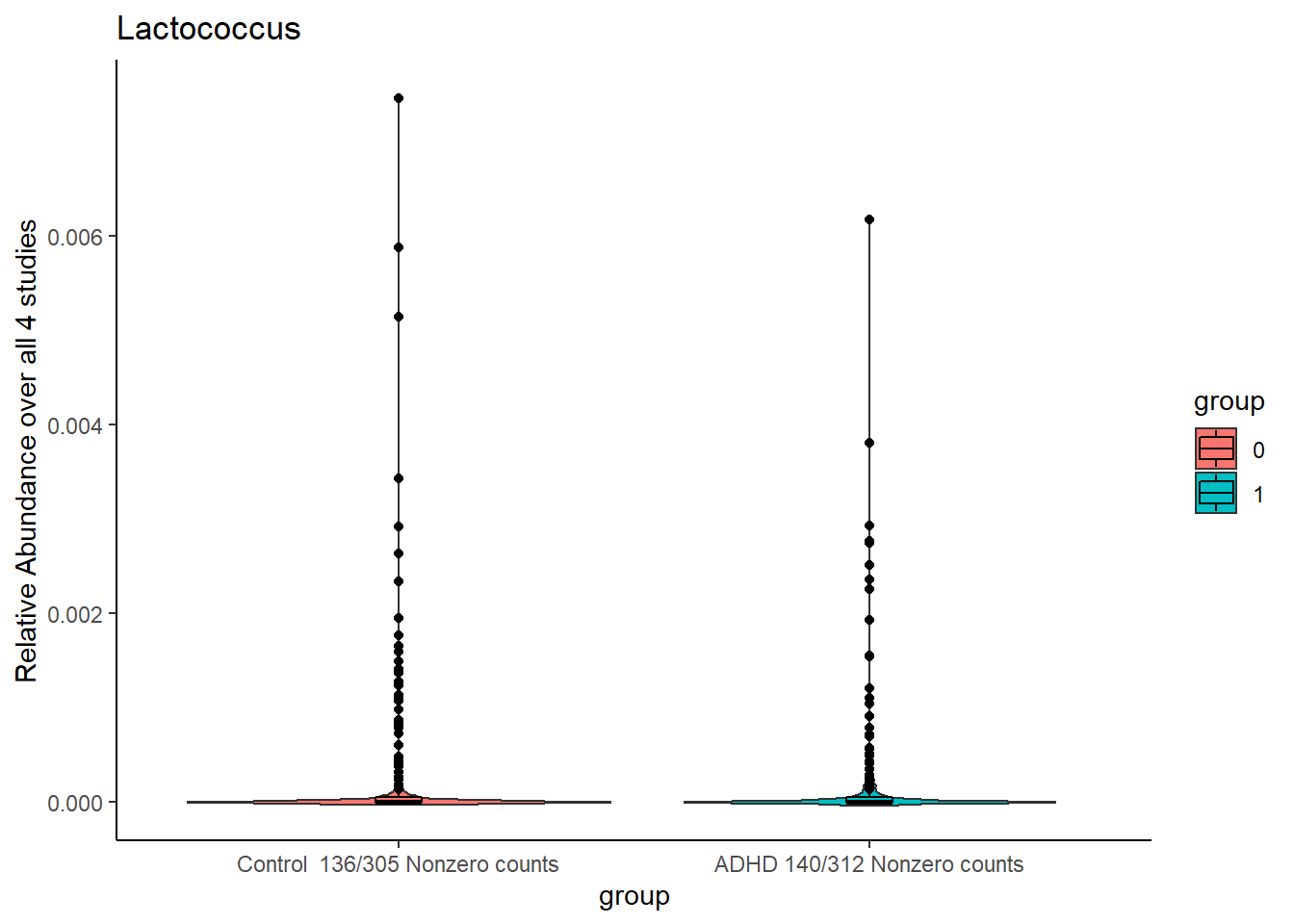

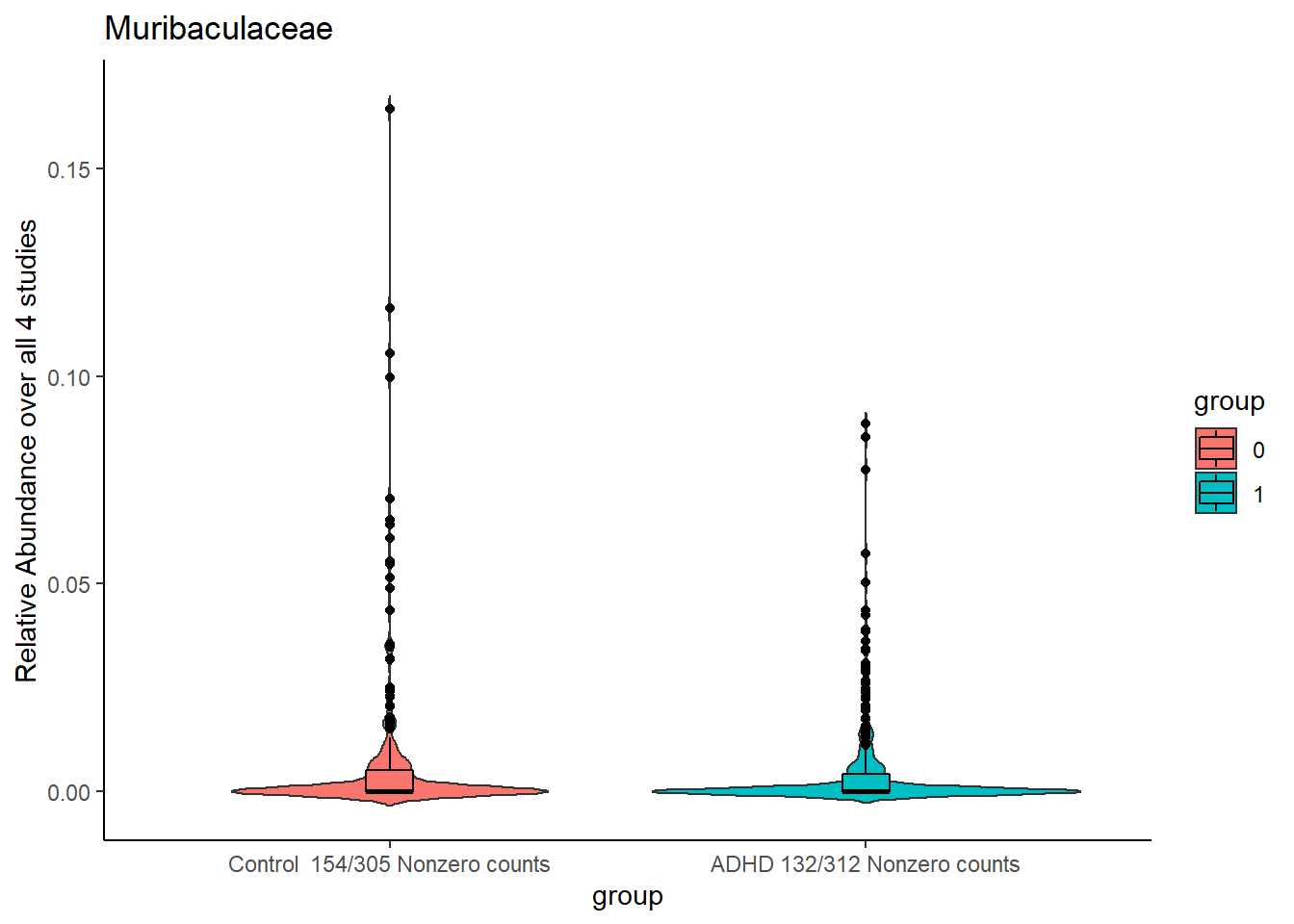

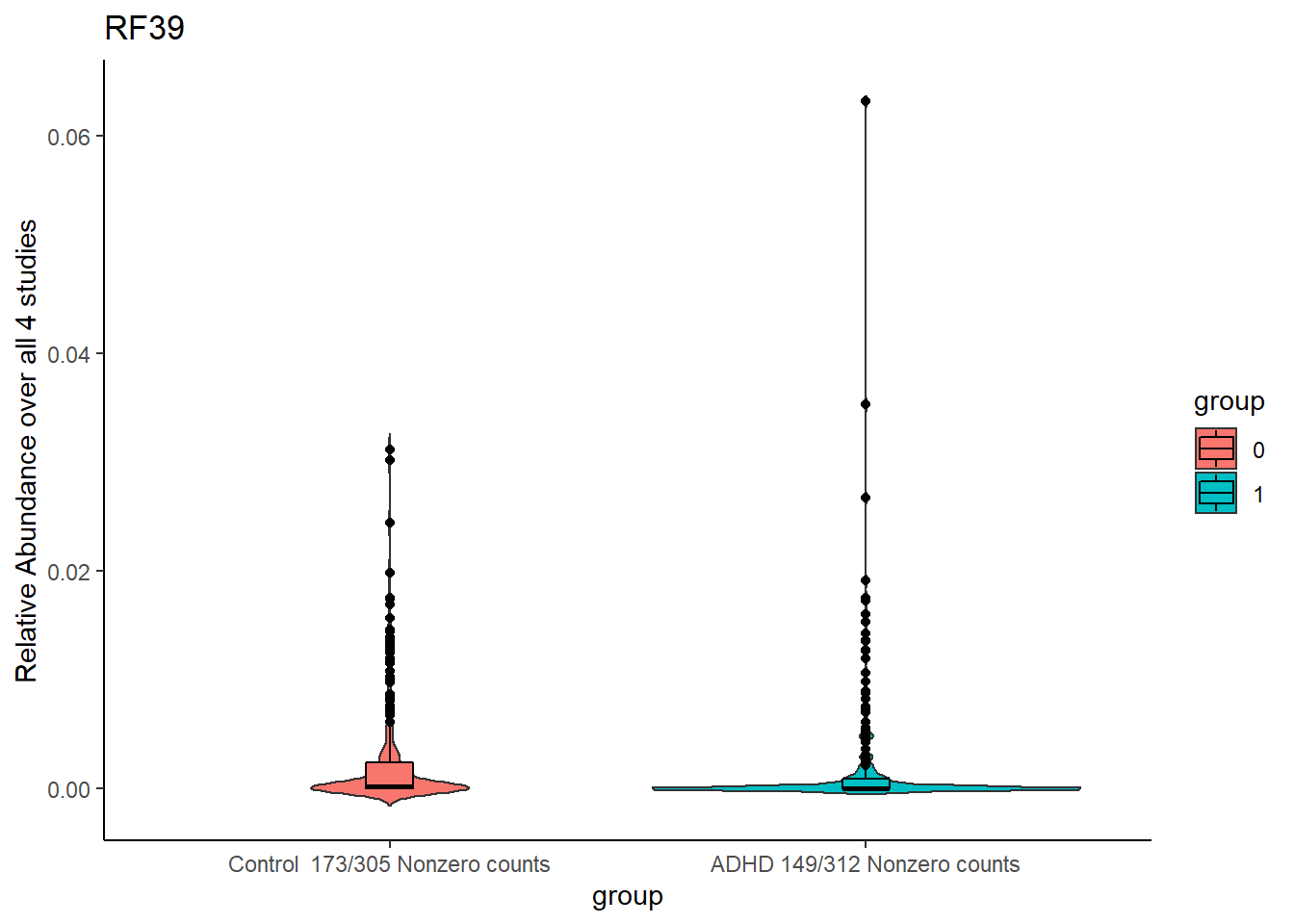

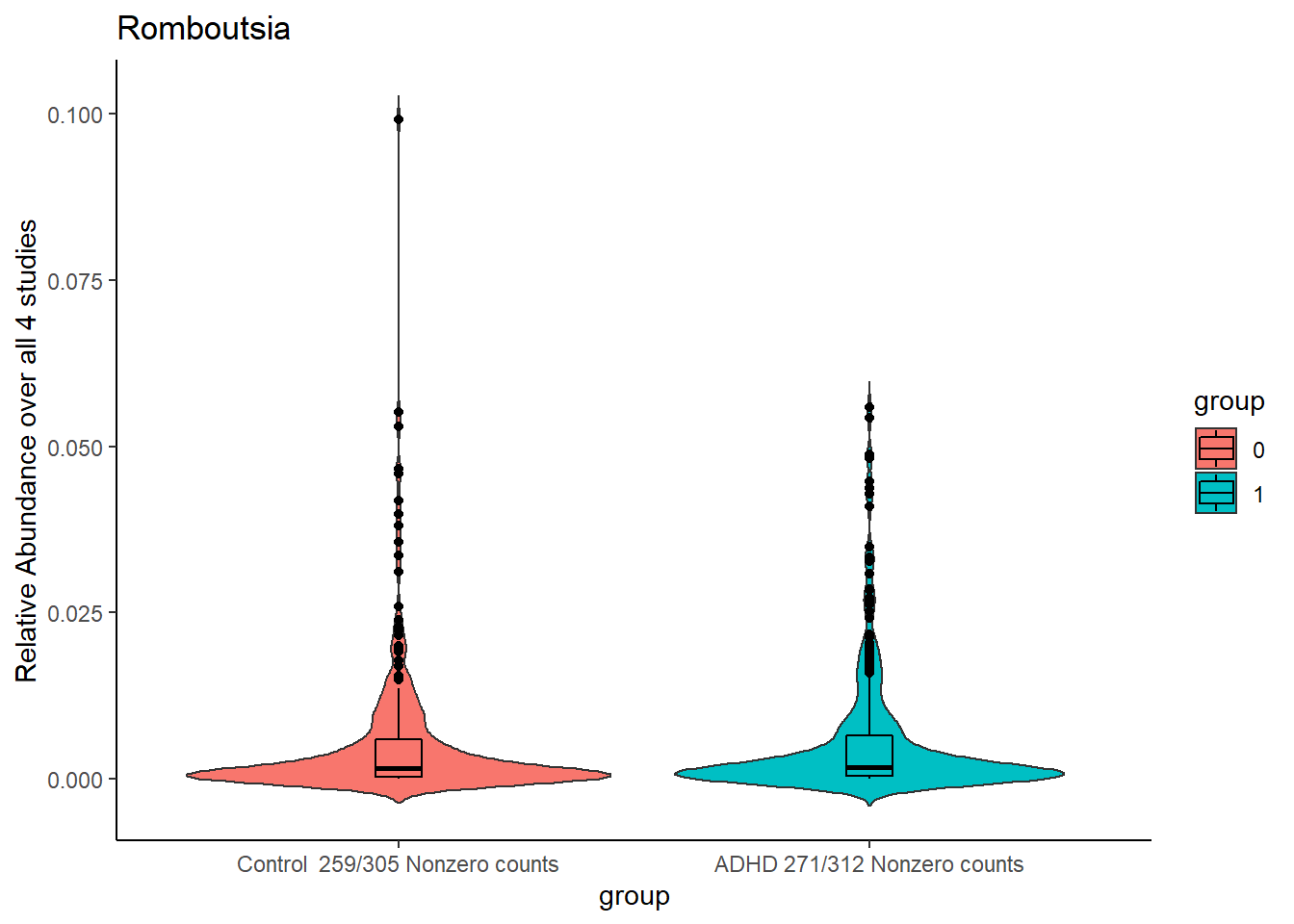

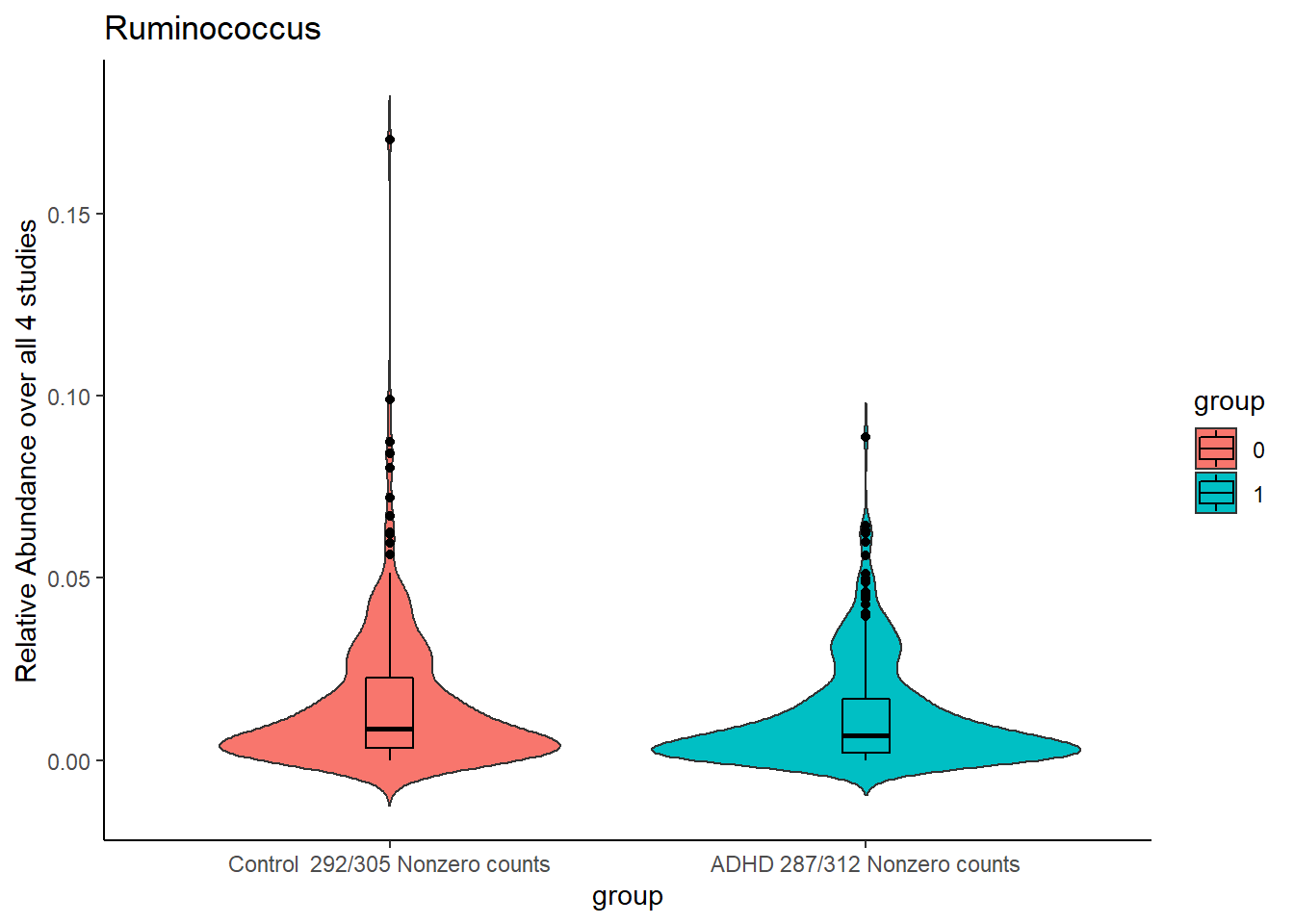

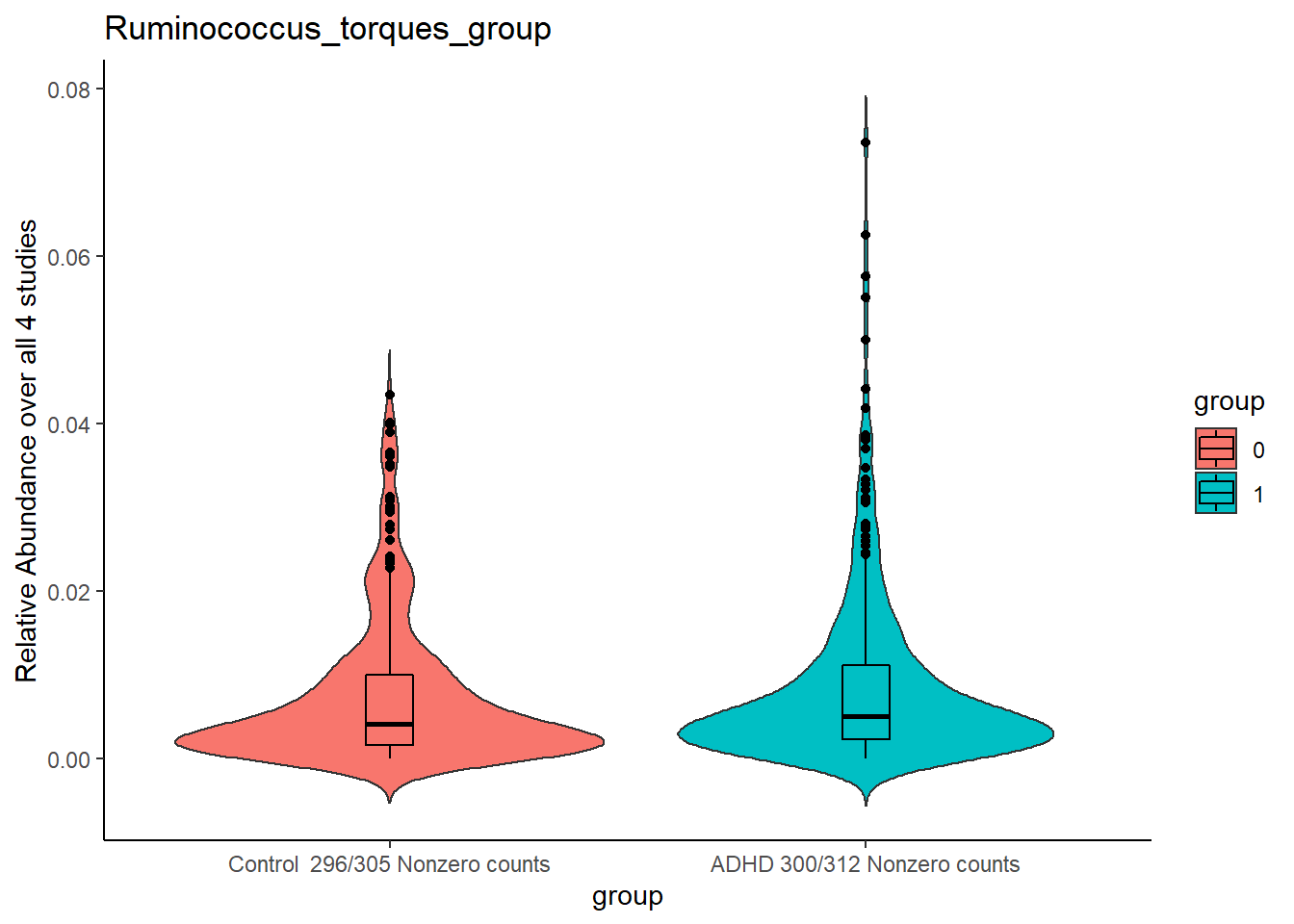

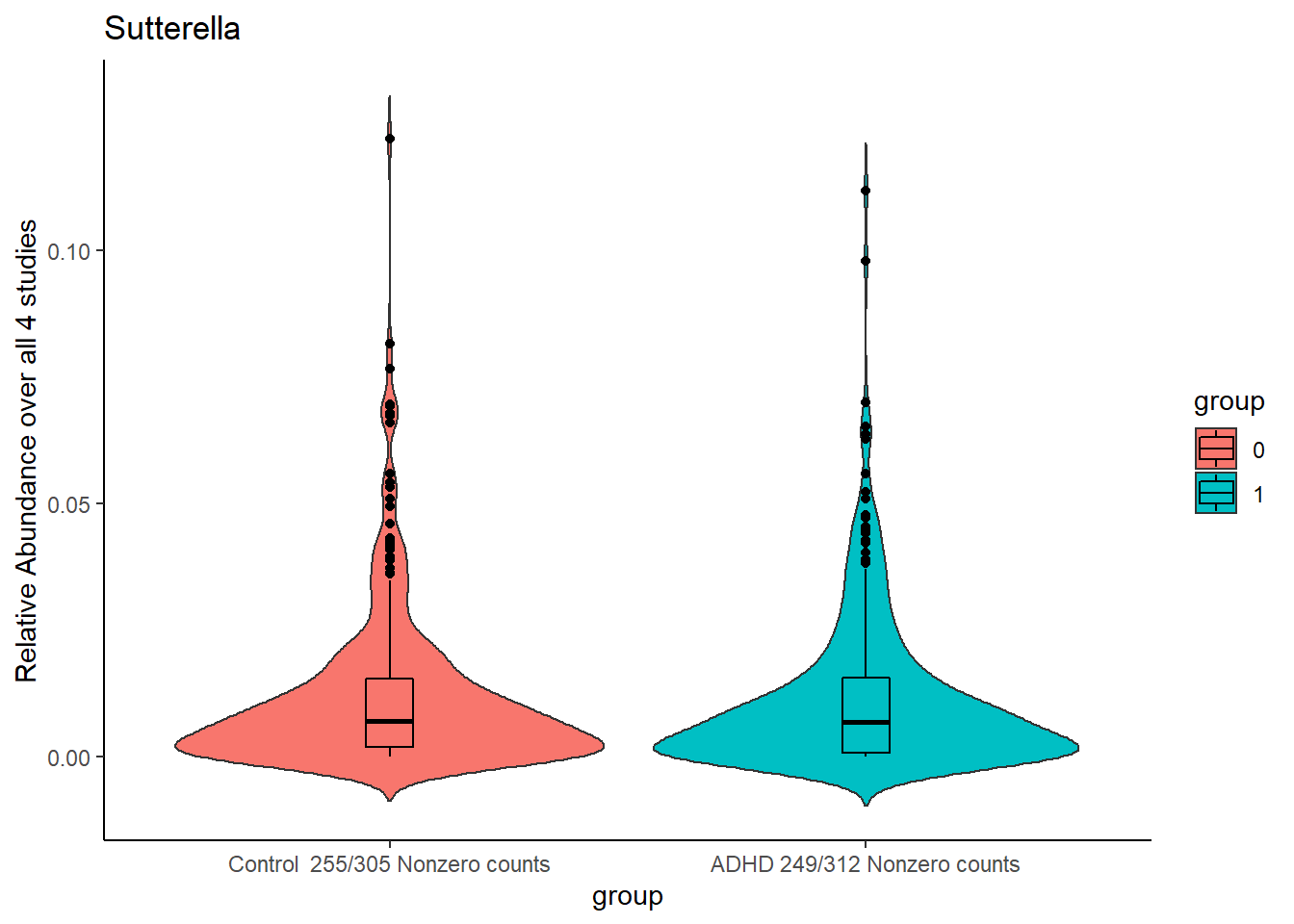

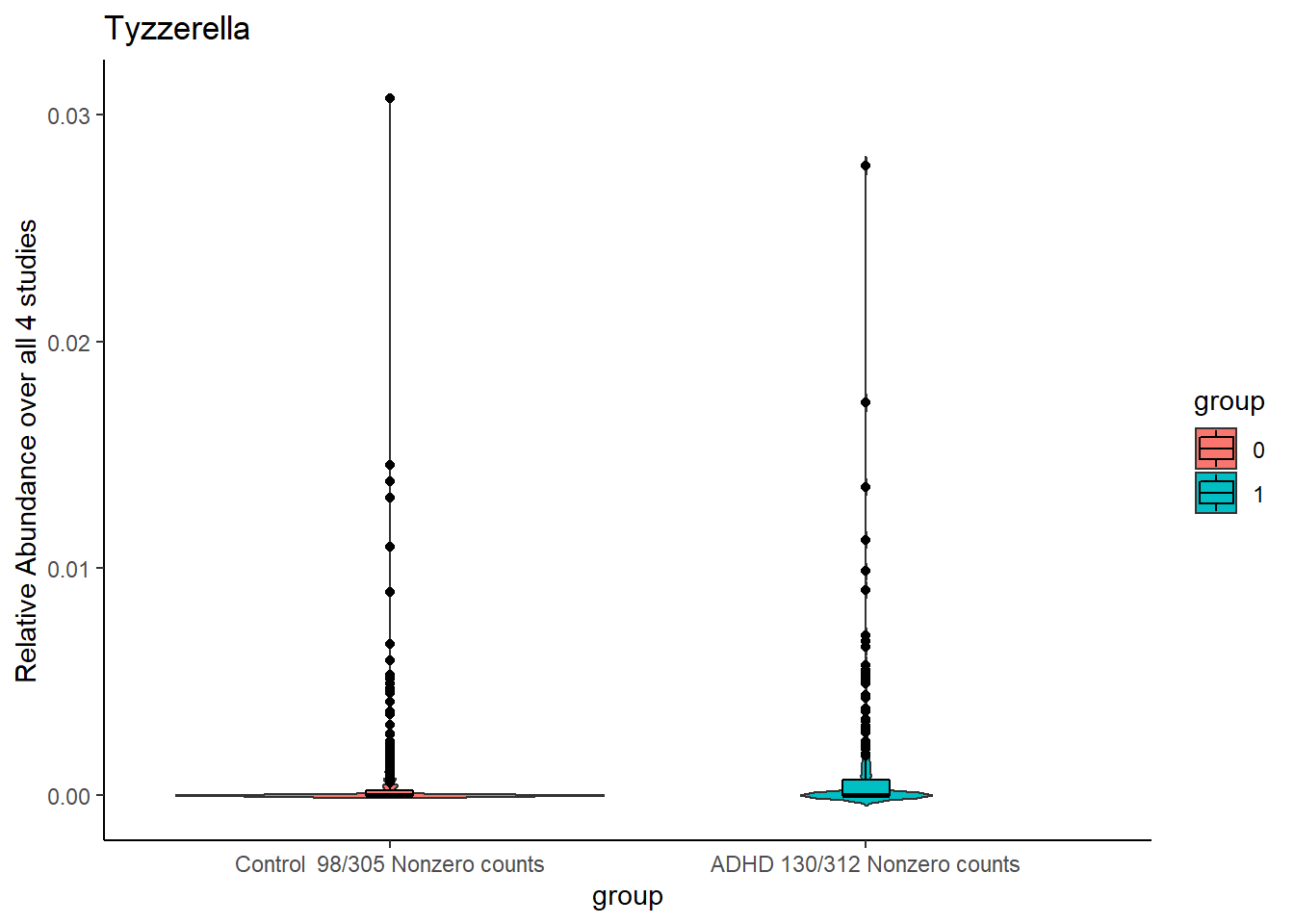

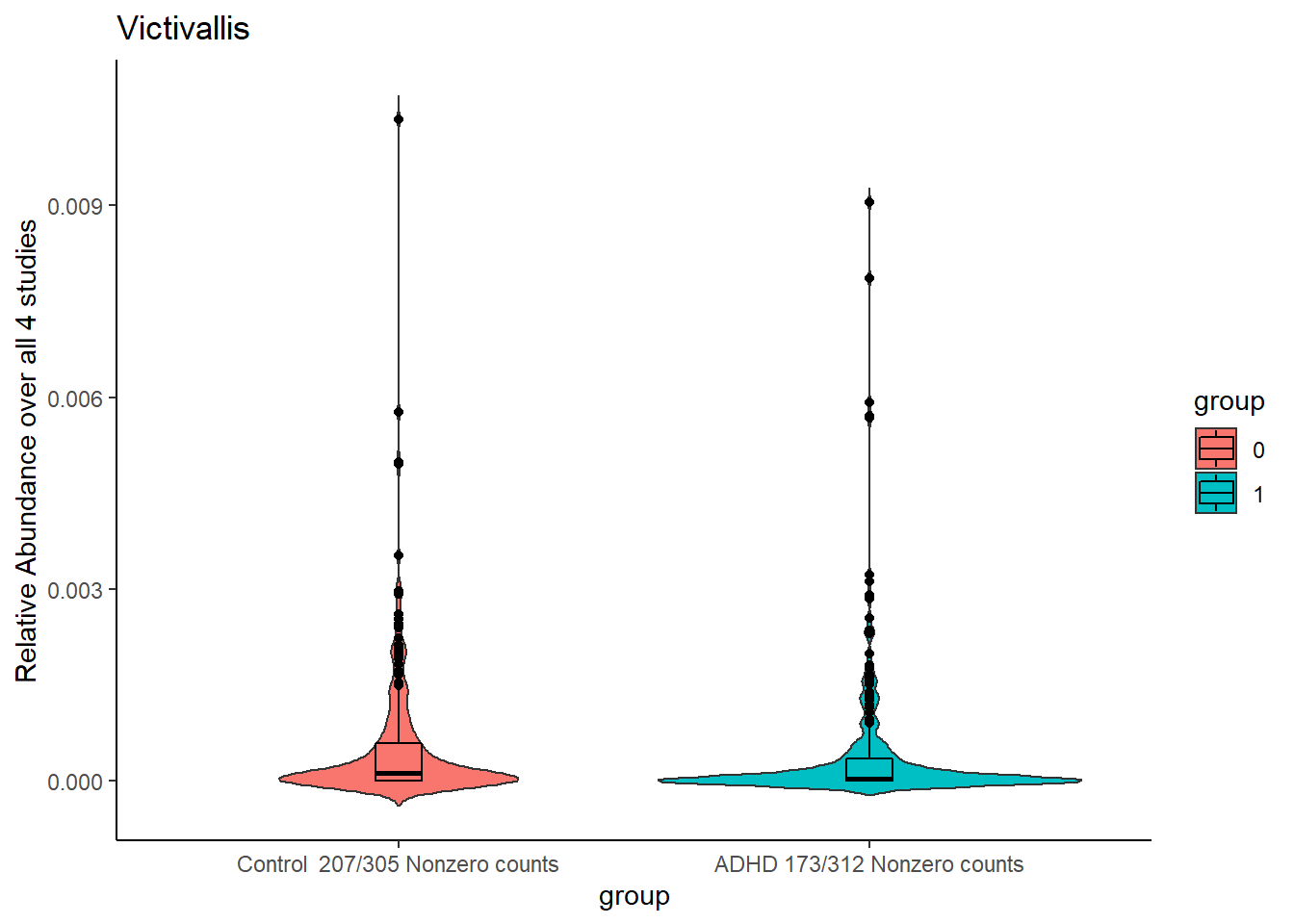

*Supplementary Figure 9. Relative Abundance aggregated over all studies of the 21 prevalent selected genera.*

7. Post-Hoc analyses

7.1 Meta-analyses of symptom associations

Supplementary Table 9. Meta-analyses results on inattention symptoms.

| Genus | pvalue | Std R | SE R | I^2^ | Het. Pvalue | Het. Q | *p*value fdr |
| --- | --- | --- | --- | --- | --- | --- | --- |
| *Eisenbergiella* | 2.90^E^-04 | 0.16 | 0.04 | 0.00 | 6.69^E^-01 | 1.56 | 1.16^E^-03 |
| *Ruminococcus torques group* | 3.30^E^-01 | 0.09 | 0.04 | 0.00 | 7.99^E^-01 | 1.01 | 6.61^E^-02 |
| *Clostridia UCG 014* | 5.96^E^-01 | -0.04 | 0.07 | 53.45 | 9.01^E^-02 | 6.49 | 5.97^E^-01 |
| *Eubacterium xylanophilum group* | 5.96^E^-01 | -0.02 | 0.04 | 0.00 | 4.21^E^-01 | 2.82 | 5.97^E^-01 |

Supplementary Table 10. Meta-analyses results on hyperactivity/impulsivity symptoms.

| Genus | pvalue | Std R | SE R | I^2^ | Het. pvalue | Het. Q | pvalue fdr |
| --- | --- | --- | --- | --- | --- | --- | --- |
| *Eisenbergiella* | 1.07E-04 | 0.14 | 0.04 | 0.00 | 4.37E-01 | 2.72 | 4.27E-03 |
| *Ruminococcus torques group* | 6.18E-03 | 0.13 | 0.05 | 8.00 | 3.96E-01 | 2.97 | 1.24E-02 |
| *Clostridia UCG 014* | 3.78E-01 | -0.05 | 0.04 | 44.89 | 1.41E-01 | 5.46 | 3.78E-01 |
| *Eubacterium xylanophilum group* | 2.09E-01 | -0.05 | 0.06 | 0.06 | 3.78E-01 | 3.08 | 2.79E-01 |

7.2 Post-hoc correction for diet

194 Participants of the MIND-Set and 157 participants of the IMpACT2-NL study additionally filled in a semiquantitative food questionnaire, asking about the frequency of meat, fruit, vegetables, legumes and sweetened beverage consumption (in the scale of never, monthly, weekly or daily). No dietary information was provided for the NeuroIMAGE and Mental-Cat sample. Polychoric correlations between variables were computed, on which exploratory factor analysis (maximum likelihood estimation and varimax rotation) was performed. Parallel analysis suggested two factors in each cohort. The resulting factors were similar in both cohorts. One factor represented the consumption of ‘healthier’ foods (vegetables, legumes and fruit), while the other factor reflected the consumption of less healthy foods (meat and sweetened beverages) (see table ST11 for factor loadings of the individual food items). We computed a diet index  by taking the absolute difference in scores of the first and the second factor. This index was intended to reflect the relative intake of healthier foods compared to less healthy foods. We introduced this diet index as a covariate into the logistic regressions at the individual study level in IMpACT2-NL and MIND-Set for the four significantly associated genera from the meta-analyses (Ruminococcus torques group, Eubacterium xylanophilum group, Eisenbergiella and Clostridia_UCG_014).

Supplementary Table 11. Factor loadings for the diet analyses

| Food Items | MIND-Set | | IMpACT2-NL | |
| --- | --- | --- | --- | --- |
|  | Factor 1 | Factor 2 | Factor 1 | Factor 2 |
| Meat | 0.02 | 0.38 | -0.22 | 0.47 |
| Fruit | 0.47 | -0.38 | 0.64 | -0.37 |
| Vegetables | 0.99 | -0.09 | 0.71 | -0.22 |
| Legumes | 0.43 | -0.04 | 0.55 | 0.07 |
| Sweet beverages | -0.13 | 0.35 | 0.03 | 0.65 |

Notably, on the individual study level, only Eisenbergiella abundance was significantly associated with ADHD diagnosis in IMpACT, the effect remained after correction for diet. All significant associations at the individual study level in MIND-Set remained significant after correction for the diet index. However, diet was significantly associated with ADHD diagnosis across all four associations in MIND-Set and subthreshold (p<.1) associated with ADHD diagnosis in IMpACT2-NL. Supplementary table 12 summarizes the results after correction for the diet index.

Supplementary Table 12. Associations of genus abundance with ADHD diagnosis corrected for diet

| Model  ADHD Diagnosis ~ Genus | | IMpACT2-NL | | | | MIND-Set | | | |
| --- | --- | --- | --- | --- | --- | --- | --- | --- | --- |
|  |  | Estimate | Std Error | t value | *p* value | Estimate | Std Error | t value | *p* value |
| Ruminococcus torques group | Diet | 0.33 | 0.17 | 1.93 | 5.34 E^-2^ | -0.21 | 0.09 | -2.19 | 2.81 E^-2^ |
|  | Genus | -0.05 | 0.17 | -0.28 | 7.77 E^-1^ | 0.32 | 0.13 | 2.55 | 1.09 E^-2^ |
| Eubacterium xylanophilum group | Diet | 0.34 | 0.17 | 1.99 | 4.62 E^-2^ | -0.21 | 0.09 | -2.29 | 2.20 E^-2^ |
|  | Genus | 0.06 | 0.06 | 0.84 | 3.98 E^-1^ | -0.13 | 0.05 | -2.21 | 2.70 E^-2^ |
| Eisenbergiella | Diet | 0.30 | 0.17 | 1.71 | 8.71 E^-2^ | -0.22 | 0.09 | -2.42 | 1.57 E^-2^ |
|  | Genus | 0.17 | 0.08 | 2.07 | 3.89 E^-2^ | 0.16 | 0.08 | 2.02 | 4.36 E^-2^ |
| Clostridia UCG 014 | Diet | 0.34 | 0.17 | 1.95 | 5.11 E^-2^ | -0.21 | 0.09 | -2.25 | 2.47 E^-2^ |
|  | Genus | 0.07 | 0.05 | 1.33 | 1.82 E^-1^ | -0.12 | 0.05 | -2.38 | 1.70 E^-2^ |

7.3 Post-hoc analyses of medication effects

We further investigated the potential effects of medication on the gut-microbiome. Significant associations of *Ruminococcus torques group, Eubacterium xylanophilum group* and *Eisenbergiella* with ADHD diagnosis in the medication naïve individuals of the Mental-Cat as well as the highly medicated sample of the MIND-Set cohort, point towards low sensitivity of these results for ADHD medication. We additionally performed a cases-only analysis of the potential effect of medication on genus abundance in IMpACT2-NL (38 of 78 participants indicated current simulant use) and NeuroIMAGE (12 of 29 participants indicated simulant use). Due to the extreme medication profiles (medication naïve participants in Mental-Cat and highly (and partly multi-)medicated individuals in MIND-Set) we could not perform this analysis in these cohorts. Supplementary Table 13 shows the results of the per study associations of genus-abundance with current use of ADHD medication in participants with ADHD. *Eisenbergiella* abundance was significantly higher in adults with ADHD medication in the NeuroIMAGE, but not the IMpACT2-NL study, while the other results showed no effect of medication. Supplementary Table 14 shows the results of the per study evaluation of associations of genus-abundance with current use of ADHD medication. Eisenbergiella abundance was significantly higher in adults with ADHD medication in the NeuroIMAGE, but not the IMpACT2-NL study.

Supplementary Table 13. Associations of medication with genus abundance

| Genus | IMpACT2-NL | | | | NeuroIMAGE | | | |
| --- | --- | --- | --- | --- | --- | --- | --- | --- |
|  | Estimate | Std Error | t value | *p* value | Estimate | Std Error | t value | *p* value |
| Ruminococcus torques group | -0.32 | 0.19 | -1.71 | 9.1 E^-1^ | 0.17 | 0.39 | 0.42 | 6.7 E^-1^ |
| Eubacterium xylanophilum group | 0.42 | 0.48 | 0.88 | 3.8 E^-1^ | -0.06 | 0.68 | 0.08 | 9.3 E^-1^ |
| Eisenbergiella | 0.25 | 0.36 | 0.70 | 4.8 E^-1^ | 0.56 | 0.25 | 2.23 | 3.5 E^-2^ |
| Clostridia UCG 014 | 0.01 | 0.52 | 0.01 | 9.9 E^-1^ | -0.02 | 0.53 | -0.05 | 9.5 E^-1^ |

8 Supplementary Table 14. STORMS guidelines

| Number | Item | Recommendation | Item Source | Additional Guidance | Yes/No/NA | Comments or location in manuscript |
| --- | --- | --- | --- | --- | --- | --- |
| **Abstract** | | | | | | |
| 1.0 | Structured or Unstructured Abstract | Abstract should include information on background, methods, results, and conclusions in structured or unstructured format. | STORMS |  | Yes |  |
| 1.1 | Study Design | State study design in abstract. | STORMS | See 3.0 for additional information on study design. | Yes |  |
| 1.2 | Sequencing methods | State the strategy used for metagenomic classification. | STORMS | For example, targeted 16S by qPCR or sequencing, shotgun metagenomics, metatranscriptomics, etc. | Yes |  |
| 1.3 | Specimens | Describe body site(s) studied. | STORMS |  | Yes |  |
| **Introduction** | | | | | | |
| 2.0 | Background and Rationale | Summarize the underlying background, scientific evidence, or theory driving the current hypothesis as well as the study objectives. | STORMS |  | Yes |  |
| 2.1 | Hypotheses | State the pre-specified hypothesis. If the study is exploratory, state any pre-specified study objectives. | STORMS |  | Yes | Exploratory |
| **Methods** | | | | | | |
| 3.0 | Study Design | Describe the study design. | STORMS | Observational (Case-Control, Cohort, Cross-sectional survey, etc.) or Experimental (Randomized controlled trial, Non-randomized controlled trial, etc.). For a brief description of common study designs see: DOI: 10.11613/BM.2014.022  If applicable, describe any blinding (e.g. single or double-blinding) used in the course of the study. | Yes |  |
| 3.1 | Participants | State what the population of interest is, and the method by which participants are sampled from that population. Include relevant information on physiological state of the subjects or stage in the life history of disease under study when participants were sampled. | STORMS | Examples of the population of interest could be: adults with no chronic health conditions, adults with type II diabetes, newborns, etc. This is the total population to whom the study is hoped to be generalizable to. The sampling method describes how potential participants were selected from that population.  If the participants are from a substudy of a larger study, provide a brief description of that study and cite that study.  Clearly state how cases and controls are defined.  An example of relevant physiological state might be pre/post menopausal for a vaginal microbiome study; examples of stage in the life history of disease could be whether specimens were collected during active or dormant disease, or before or after treatment. | Yes | Methods and Materials + Supplementary Chapter 1.1 |
| 3.2 | Geographic location | State the geographic region(s) where participants were sampled from. | MIxS: geographic location (country and/or sea,region) | Geographic coordinates can be reported to prevent potential ambiguities if necessary. | Yes | Table 1 |
| 3.3 | Relevant Dates | State the start and end dates for recruitment, follow-up, and data collection. | STORMS | Recruitment is the period in which participants are recruited for the study. In longitudinal studies, follow-up is the date range in which participants are asked to complete a specific assessment. Finally, data collection is the total period in which data is being collected from participants including during initial recruitment through all follow-ups. | Yes | Table 1 |
| 3.4 | Eligibility criteria | List any criteria for inclusion and exclusion of recruited participants. | Modified STROBE | Among potential recruited participants, how were some chosen and others not? This could include criteria such as sex, diet, age, health status, or BMI.  If there is a primary and validation sample, describe inclusion/exclusion criteria for each. | Yes | Supplementary Chapter 1.1 |
| 3.5 | Antibiotics Usage | List what is known about antibiotics usage before or during sample collection. | STORMS | If participants were excluded due to current or recent antibiotics usage, state this here.  Other factors (e.g. proton pump inhibitors, probiotics, etc.) that may influence the microbiome should also be described as well. | Partly | Information with sufficient detail was only acquired in MIND-SET, exclusion criterium in Mental-Cat |
| 3.6 | Analytic sample size | Explain how the final analytic sample size was calculated, including the number of cases and controls if relevant, and reasons for dropout at each stage of the study. This should include the number of individuals in whom microbiome sequencing was attempted and the number in whom microbiome sequencing was successful. | STORMS | Consider use of a flow diagram (see template at https://stormsmicrobiome.org/figures). Also state sample size in abstract.  If power analysis was used to calculate sample size, describe those calculations. | Partly | See Methods section and supplementary chapter 1.1. Note, information of attempted sequencing was not available for Mental-Cat and NeuroIMAGE |
| 3.7 | Longitudinal Studies | For longitudinal studies, state how many follow-ups were conducted, describe sample size at follow-up by group or condition, and discuss any loss to follow-up. | STORMS | If there is loss to follow-up, discuss the likelihood that drop-out is associated with exposures, treatments, or outcomes of interest. | NA |  |
| 3.8 | Matching | For matched studies, give matching criteria. | Modified STROBE | "Matched" refers to matching between comparable study participants as cases and controls or exposed / unexposed.  Indicate whether participants were individual or frequency matched and in what ratio were they matched (e.g. 1 case to 1 control). | NA |  |
| 3.9 | Ethics | State the name of the institutional review board that approved the study and protocols, protocol number and date of approval, and procedures for obtaining informed consent from participants. | STORMS |  | Partly | Note, protocol number and date of approval are not provided |
| 4.0 | Laboratory methods | State the laboratory/center where laboratory work was done. | STORMS | Provide a reference to complete lab protocols if previously published elsewhere such as on protocols.io. Note any modifications of lab protocols and the reason for protocol modifications. | Partly | Information only available for MIND-Set and IMpACT2-NL, Supplementary Chapter 2 |
| 4.1 | Specimen collection | State the body site(s) sampled from and how specimens were collected. | MIxS: sample collection device or method; host body site | Use terms from the Uber-anatomy Ontology (https://www.ebi.ac.uk/ols/ontologies/uberon) to describe body sites in a standardized format. | Yes | Supplementary Chapter 2 |
| 4.2 | Shipping | Describe how samples were stored and shipped to the laboratory. | STORMS | Include length of time from collection to receipt by the lab and if temperature control was used during shipping. | Partly | Supplementary Chapter 2, information only available for IMpACT2-NL and MIND-Set |
| 4.3 | Storage | Describe how the laboratory stored samples, including time between collection and storage and any preservation buffers or refrigeration used. | STORMS | State where each procedure or lot of samples was done if not all in the same place.  Include reagent/lot/catalogue #s for storage buffers. | Partly | Supplementary Chapter 2, information only available for IMpACT2-NL and MIND-Set |
| 4.4 | DNA extraction | Provide DNA extraction method, including kit and version if relevant. | MIxS: nucleic acid extraction | If any DNA quantification methods were used prior to DNA amplification or at the pooling step of library preparation, state so here. | Partly | Supplementary Chapter 2, information only available for IMpACT2-NL and MIND-Set |
| 4.5 | Human DNA sequence depletion or microbial DNA enrichment | Describe whether human DNA sequence depletion or enrichment of microbial or viral DNA was performed. | STORMS |  | NA |  |
| 4.6 | Primer selection | Provide primer selection and DNA amplification methods as well as variable region sequenced (if applicable). | MIxS: pcr primers |  | Yes | Methods section |
| 4.7 | Positive Controls | Describe any positive controls (mock communities) if used. | STORMS | If used, should be deposited under guidance provided in the 8.X items. | Yes | Supplementary Section 1.1 |
| 4.8 | Negative Controls | Describe any negative controls if used. | STORMS | If used, should be deposited under guidance provided in the 8.X items. | Yes | Supplementary Section 1.1 |
| 4.9 | Contaminant mitigation and identification | Provide any laboratory or computational methods used to control for or identify microbiome contamination from the environment, reagents, or laboratory. | STORMS | Includes filtering of reagents and other steps to minimize contamination. It is relevant to state whether the specimens of interest have low microbial load, which makes contamination especially relevant. | NA | Baseclear inclusion of negative water control and sterile lab environment |
| 4.10 | Replication | Describe any biological or technical replicates included in the sequencing, including which steps were replicated between them. | STORMS | Replication may be biological (redundant biological specimens) or technical (aliquots taken at different stages of analysis) and used in extraction, sequencing, preprocessing, and/or data analysis. | NA |  |
| 4.11 | Sequencing strategy | Major divisions of strategy, such as shotgun or amplicon sequencing. | MIxS: sequencing method | For amplicon sequencing (for example, 16S variable region), state the region selected. State the model of sequencer used. | Yes | Methods section |
| 4.12 | Sequencing methods | State whether experimental quantification was used (QMP/cell count based, spike-in based) or whether relative abundance methods were applied. | STORMS | These include read length, sequencing depth per sample (average and minimum), whether reads are paired, and other parameters. | NA |  |
| 4.13 | Batch effects | Detail any blocking or randomization used in study design to avoid confounding of batches with exposures or outcomes. Discuss any likely sources of batch effects, if known. | STORMS | Sources of batch effects include sample collection, storage, library preparation, and sequencing and are commonly unavoidable in all but the smallest of studies. | NA | No randomization, batches are filled with samples according to the date they were received across MIND-Set and IMpACT, mixing control subjects and participants with ADHD |
| 4.14 | Metatranscriptomics | Detail whether any mRNA enrichment was performed and whether/how retrotranscription was performed prior to sequencing. Provide size range of isolated transcripts. Describe whether the sequencing library was stranded or not. Provide details on sequencing methods and platforms. | STORMS | Provide details on any internal standards which may have been used as well as parameters and versions of any software or databases used. | NA |  |
| 4.15 | Metaproteomics | Detail which protease was used for digestion. Provide details on proteomic methods and platforms (e.g. LC-MS/MS, instrument type, column type, mass range, resolution, scan speed, maximum injection time, isolation window, normalised collision energy, and resolution). | STORMS | Provide details on any internal standards which may have been used as well as parameters and versions of any software or databases used. | NA |  |
| 4.16 | Metabolomics | Specify the analytic method used (such as nuclear magnetic resonance spectroscopy or mass spectrometry). For mass spectrometry, detail which fractions were obtained (polar and/or non-polar) and how these were analyzed. Provide details on metabolomics methods and platforms (e.g. derivatization, instrument type, injection type, column type and instrument settings). | STORMS | Provide details on any internal standards which may have been used as well as parameters and versions of any software or databases used. | NA |  |
| 5.0 | Data sources/  measurement | For each non-microbiome variable, including the health condition, intervention, or other variable of interest, state how it was defined, how it was measured or collected, and any transformations applied to the variable prior to analysis. | MIxS: host disease status | State any sources of potential bias in measurements, for example multiple interviewers or measurement instruments, and whether these potential biases were assessed or accounted for in study design.  Use terms from a standardized ontology such as the Experimental Factor Ontology (https://www.ebi.ac.uk/efo/) to describe variables of interest in a standardized format. | Yes | Methods section, Discussion, Supplementary Chapter 1.1 |
| 6.0 | Research design for causal inference | Discuss any potential for confounding by variables that may influence both the outcome and exposure of interest. State any variables controlled for and the rationale for controlling for them. | STORMS | For causal inference, this item refers to describing the assumptions that would be required to draw causal inferences from observational data. See Vujkovic-Cvijin, I., Sklar, J., Jiang, L. et al. Host variables confound gut microbiota studies of human disease. Nature 587, 448–454 (2020). https://doi.org/10.1038/s41586-020-2881-9 for more details on confounding in observational microbiome studies.  For example, hypothesized confounders may be controlled for by multivariable adjustment. Consider using a directed acyclic graph (DAG) to describe your causal model and justify any variables controlled for. DAGs can be made using [www.dagitty.net](http://www.dagitty.net/). | NA |  |
| 6.1 | Selection bias | Discuss potential for selection or survival bias. | STORMS | Selection bias can occur when some members of the target study population are more likely to be included in the study/final analytic sample than others. Some examples include survival bias (where part of the target study population is more likely to die before they can be studied), convenience sampling (where members of the target study population are not selected at random), and loss to follow-up (when probability of dropping out is related to one of the things being studied). | NA | Feature selection was applied, while bias is introduced underestimating low prevalent features, the data-driven selection instead of hypothesis driven might on the other hand reduce bias |
| 7.0 | Bioinformatic and Statistical Methods | Describe any transformations to quantitative variables used in analyses (e.g. use of percentages instead of counts, normalization, rarefaction, categorization). | STORMS | If a variable is analyzed using different transformations, state rationale for the transformation and for each analyses which version of the variable is used.  In case of any complex or multistep transformations, give enumerated instructions for reproducing those transformations. | Yes | Methods section, Supplementary Chapter 2 |
| 7.1 | Quality Control | Describe any methods to identify or filter low quality reads or samples. | MIxS: sequence quality check | If samples were excluded based on quality or read depth, list the criteria used, the number of samples excluded, and the final sample size after quality control. | Yes | Methods section |
| 7.2 | Sequence analysis | Describe any taxonomic, functional profiling, or other sequence analysis performed. | MIxS: feature prediction; similarity search method |  | Yes | Methods section |
| 7.3 | Statistical methods | Describe all statistical methods. | Modified STROBE | Describe any statistical tests used, exploratory data analysis performed, dimension reduction methods/unsupervised analysis, alpha/beta metrics, and/or methods for adjusting for measurement bias.  If multiple statistical methods are possible, discuss why the methods used were selected.  If a multiple hypothesis testing correction method was used, describe the type of correction used.  State which taxonomic levels are analyzed. | Yes | Methods section |
| 7.4 | Longitudinal analysis | If the study is longitudinal, include a section that explicitly states what analysis methods were used (if any) to account for grouping of measurements by individual or patterns over time. | STORMS |  | NA |  |
| 7.5 | Subgroup analysis | Describe any methods used to examine subgroups and interactions. | STROBE |  | NA |  |
| 7.6 | Missing data | Explain how missing data were addressed. | STROBE | "Missing data" refers to participant measurements such as covariates, exposures, outcomes, or time points that should have been collected but were not, not to zeros in taxonomic abundance tables or data points not applicable to that observation. | Yes | missing data was strictly excluded |
| 7.7 | Sensitivity analyses | Describe any sensitivity analyses. | STROBE |  | Yes | Supplementary Chapter 7 |
| 7.8 | Findings | State criteria used to select findings for reporting. | STORMS | For example, false discovery rate with total number of tests, effect size threshold, significance threshold, microbes of interest. | Yes | Convergence across tools, meta-analysis, fdr corrected |
| 7.9 | Software | Cite all software (including read mapping software) and databases (including any used for taxonomic reference or annotating amplicons, if applicable) used. Include version numbers. | Modified STREGA | Installed packages, add-ons or libraries should be stated and cited in addition to the software used.  All parameters employed that differ from the default of that software/version should be provided.  This is in addition to, not a replacement for, publishing of code as outlined in the section Reproducible Research. | Yes | Methods section |
| 8.0 | Reproducible research | Make a statement about whether and how others can reproduce the reported analysis. | STORMS | Any protected information that has been excluded or provided under controlled access should be listed along with any relevant data access procedures. "On request from authors" is not sufficiently detailed; formal data access procedures and conditions should be defined.  If data are unavailable, state so clearly.  Consider using a specialized rubric for reproducible research (such as:<https://mbio.asm.org/content/9/3/e00525-18.short)>.  Consider preregistering the study protocol (such as o[n osf.](http://osf.io/)io or<https://plos.org/open-science/preregistration/).> | NA | We are not owners of the individual study data, raw sequences are available upon request, summary statistics are attached to the supplement |
| 8.1 | Raw data access | State where raw data may be accessed including demultiplexing information. | STORMS | Robust, long-term databases such as those hosted by NCBI and EBI are preferred. If using a private repository, provide rationale. | NA |  |
| 8.2 | Processed data access | State where processed data may be accessed. | STORMS | Unfiltered data should be provided.  Robust, long-term databases such as those hosted by NCBI and EBI-EMBL are preferred. Repositories like zenodo (https://zenodo.org/) or publisso (https://www.publisso.de/en/working-for-you/doi-service/)  can be used to provide a DOI and long-term storage for processed datasets, even those which cannot be published openly. | NA |  |
| 8.3 | Participant data access | State where individual participant data such as demographics and other covariates may be accessed, and how they can be matched to the microbiome data. | STORMS | If re-categorized, transformed, or otherwise derived variables were used in the analysis, these variables or code for deriving them should be provided.  Examples of how participant data can be matched to microbiome data are: using the same set of anonymized identifiers, or using different anonymized identifiers but providing a map.  Provided data should be sufficient to independently replicate the current analysis. | NA |  |
| 8.4 | Source code access | State where code may be accessed. | STORMS | If a standard or formalized workflow was employed, reference it here. | Yes | Methods section |
| 8.5 | Full results | Provide full results of all analyses, in computer-readable format, in supplementary materials. | STORMS | For example, any fold-changes, p-values, or FDR values calculated, provided as a spreadsheet.  Use a machine-readable, plain-text format such as csv or tsv. | Yes | Provided in supplementary materials (word format) |
| **Results** | | | | | | |
| 9.0 | Descriptive data | Give characteristics of study participants (e.g. dietary, demographic, clinical, social) and information on exposures and potential confounders. | STROBE | Typically reported in a table included in the paper or as a supplementary table. Indicate number of participants with missing data for each variable of interest.  This includes environmental and lifestyle factors that may affect the relationship between the microbiome and the condition of interest. Participant diet and medication use should be summarized, if known.  At minimum, age and sex of all participants should be summarized. | Yes | Methods section |
| 10.0 | Microbiome data | Report descriptive findings for microbiome analyses with all applicable outcomes and covariates. | STORMS | This includes measures of diversity as well as relative abundances. These descriptive findings should be reported both for the sample overall and for individual groups. | Yes | Results section, Supplementary Materials |
| 10.1 | Taxonomy | Identify taxonomy using standardized taxon classifications that are sufficient to uniquely identify taxa. | STORMS | If not using full taxonomic hierarchy, make sure it is clear whether names stated are species, genera, family, etc.  Italicize genus/species pairs. Consult journal guidelines or standardized references on taxonomic nomenclature. For instance,<https://wwwnc.cdc.gov/eid/page/scientific-nomenclature> | Yes | Genus only |
| 10.2 | Differential abundance | Report results of differential abundance analysis by the variable of interest and (if applicable) by time, clearly indicating the direction of change and total number of taxa tested. | STORMS | If there are more than two groups, include omnibus (multigroup) test results if applicable to the research question.  If applicable, reported effect sizes should include a measure of uncertainty such as the confidence interval. | Yes | Results, Supplementary chapter 6 |
| 10.3 | Other data types | Report other data analyzed--e.g. metabolic function, functional potential, MAG assembly, and RNAseq. | STORMS |  | NA |  |
| 10.4 | Other statistical analysis | Report any statistical data analysis not covered above. | STORMS | This could include subgroup analysis, sensitivity analyses, and cluster analysis.  Visualizations should be easily interpretable and colorblind-friendly. The caption and/or main text should provide a detailed description of visualizations for visually-impaired readers. | Yes |  |
| **Discussion** | | | | | | |
| 11.0 | Key results | Summarise key results with reference to study objectives | STROBE |  | Yes |  |
| 12.0 | Interpretation | Give a cautious overall interpretation of results considering objectives, limitations, multiplicity of analyses, results from similar studies, and other relevant evidence. | STROBE | Define or clarify any subjective terms such as "dominant," "dysbiosis," and similar words used in interpretation of results.  When interpreting the findings, consider how the interpretation of the findings may be summarized or quoted for the general public such as in press releases or news articles.  If causal language is used in the interpretation (such as "alters," "affects," "results in," "causes," or "impacts"), assumptions made for causal inference should be explicitly stated as part of 6.0 and 13.0.  Distinguish between function potential (ie inferred from metagenomics) and observed activity (ie metatranscriptomic, metabolomic, proteomic) if discussing microbial function. | Yes |  |
| 13.0 | Limitations | Discuss limitations of the study, taking into account sources of potential bias or imprecision. | STROBE | Also consider limitations resulting from the methods (especially novel methods), the study design, and the sample size. | Yes |  |
| 13.1 | Bias | Discuss any potential for bias to influence study findings. | STORMS | May include sampling method, representativeness of study participants, or potential confounding. | Yes |  |
| 13.2 | Generalizability | Discuss the generalisability (external validity) of the study results | STROBE | To what populations or other settings do you expect the conclusions to generalize? | NA |  |
| 14.0 | Ongoing/future work | Describe potential future research or ongoing research based on the study's findings. | STORMS |  | Yes |  |
| **Other information** | | | | | | |
| 15.0 | Funding | Give the source of funding and the role of the funders for the present study and, if applicable, for the original study on which the present article is based | STROBE |  | Partly | Acknowledgements, Where information was provided |
| 15.1 | Acknowledgements | Include acknowledgements of those who contributed to the research but did not meet critera for authorship. | STORMS | For general guidelines on authorship, see [http://www.icmje.org](http://www.icmje.org/) and<https://www.elsevier.com/authors/journal-authors/policies-and-ethics/credit-author-statement> | NA |  |
| 15.2 | Conflicts of Interest | Include a conflicts of interest statement. | STORMS |  | Yes | Disclosure |
| 16.0 | Supplements | Indicate where supplements may be accessed and what materials they contain. | STORMS |  | Yes | In text |
| 17.0 | Supplementary data | Provide supplementary data files of results with for all taxa and all outcome variables analyzed. Indicate the taxonomic level of all taxa. | STORMS | Depending on the analysis performed, examples of the supplemental results included could be mean relative abundance, differential abundance, raw p-value, multiple hypothesis testing-adjusted p-values, and standard error.  All discussed taxa should include the taxonomic level (e.g. class, order, genus). | Yes |  |

9 Supplementary Table 15. PRISMA Checklist

| **Section and Topic** | **Item** | **Checklist item** | **Location where item is reported** |
| --- | --- | --- | --- |
| **TITLE** | | |  |
| Title | 1 | Identify the report as a systematic review. | NA, report is a meta-analysis |
| **ABSTRACT** | | |  |
| Abstract | 2 | See the PRISMA 2020 for Abstracts checklist. | Abstract |
| **INTRODUCTION** | | |  |
| Rationale | 3 | Describe the rationale for the review in the context of existing knowledge. | NA |
| Objectives | 4 | Provide an explicit statement of the objective(s) or question(s) the review addresses. | NA |
| **METHODS** | | |  |
| Eligibility criteria | 5 | Specify the inclusion and exclusion criteria for the review and how studies were grouped for the syntheses. | Supplementary section 1 |
| Information sources | 6 | Specify all databases, registers, websites, organisations, reference lists and other sources searched or consulted to identify studies. Specify the date when each source was last searched or consulted. | Supplementary figure 15 |
| Search strategy | 7 | Present the full search strategies for all databases, registers and websites, including any filters and limits used. | Supplementary figure 15 |
| Selection process | 8 | Specify the methods used to decide whether a study met the inclusion criteria of the review, including how many reviewers screened each record and each report retrieved, whether they worked independently, and if applicable, details of automation tools used in the process. | Supplementary figure 15 |
| Data collection process | 9 | Specify the methods used to collect data from reports, including how many reviewers collected data from each report, whether they worked independently, any processes for obtaining or confirming data from study investigators, and if applicable, details of automation tools used in the process. | Supplementary figure 15 |
| Data items | 10a | List and define all outcomes for which data were sought. Specify whether all results that were compatible with each outcome domain in each study were sought (e.g. for all measures, time points, analyses), and if not, the methods used to decide which results to collect. | Supplementary figure 15, supplementary section 1 |
|  | 10b | List and define all other variables for which data were sought (e.g. participant and intervention characteristics, funding sources). Describe any assumptions made about any missing or unclear information. | NA |
| Study risk of bias assessment | 11 | Specify the methods used to assess risk of bias in the included studies, including details of the tool(s) used, how many reviewers assessed each study and whether they worked independently, and if applicable, details of automation tools used in the process. | NA (not a review, only 2 studies received) |
| Effect measures | 12 | Specify for each outcome the effect measure(s) (e.g. risk ratio, mean difference) used in the synthesis or presentation of results. | Re-analyzed, see Methods section Manuscript |
| Synthesis methods | 13a | Describe the processes used to decide which studies were eligible for each synthesis (e.g. tabulating the study intervention characteristics and comparing against the planned groups for each synthesis (item #5)). | Supplementary figure 15 |
|  | 13b | Describe any methods required to prepare the data for presentation or synthesis, such as handling of missing summary statistics, or data conversions. | Data request, see supplementary section 1 |
|  | 13c | Describe any methods used to tabulate or visually display results of individual studies and syntheses. | NA |
|  | 13d | Describe any methods used to synthesize results and provide a rationale for the choice(s). If meta-analysis was performed, describe the model(s), method(s) to identify the presence and extent of statistical heterogeneity, and software package(s) used. | See Methods section Manuscript |
|  | 13e | Describe any methods used to explore possible causes of heterogeneity among study results (e.g. subgroup analysis, meta-regression). | See Methods section Manuscript |
|  | 13f | Describe any sensitivity analyses conducted to assess robustness of the synthesized results. | See supplementary section 7 |
| Reporting bias assessment | 14 | Describe any methods used to assess risk of bias due to missing results in a synthesis (arising from reporting biases). | Only 2 included studies,assessment skew |
| Certainty assessment | 15 | Describe any methods used to assess certainty (or confidence) in the body of evidence for an outcome. | Convergence, see Methods and Discussion section |
| **RESULTS** | | | |
| Study selection | 16a | Describe the results of the search and selection process, from the number of records identified in the search to the number of studies included in the review, ideally using a flow diagram. | See Methods section Manuscript |
|  | 16b | Cite studies that might appear to meet the inclusion criteria, but which were excluded, and explain why they were excluded. | See Methods section Manuscript |
| Study characteristics | 17 | Cite each included study and present its characteristics. | See supplementary section 1 |
| Risk of bias in studies | 18 | Present assessments of risk of bias for each included study. | NA |
| Results of individual studies | 19 | For all outcomes, present, for each study: (a) summary statistics for each group (where appropriate) and (b) an effect estimate and its precision (e.g. confidence/credible interval), ideally using structured tables or plots. | See supplementary section 3, 4 and 5 |
| Results of syntheses | 20a | For each synthesis, briefly summarise the characteristics and risk of bias among contributing studies. | NA |
|  | 20b | Present results of all statistical syntheses conducted. If meta-analysis was done, present for each the summary estimate and its precision (e.g. confidence/credible interval) and measures of statistical heterogeneity. If comparing groups, describe the direction of the effect. | See Supplementary results and results section |
|  | 20c | Present results of all investigations of possible causes of heterogeneity among study results. | See Supplementary results and results section |
|  | 20d | Present results of all sensitivity analyses conducted to assess the robustness of the synthesized results. | See Supplementary section 7 |
| Reporting biases | 21 | Present assessments of risk of bias due to missing results (arising from reporting biases) for each synthesis assessed. | NA |
| Certainty of evidence | 22 | Present assessments of certainty (or confidence) in the body of evidence for each outcome assessed. | See supplementary section 6 and Results section in Manuscript |
| **DISCUSSION** | | |  |
| Discussion | 23a | Provide a general interpretation of the results in the context of other evidence. | See Discussion in Manuscript |
|  | 23b | Discuss any limitations of the evidence included in the review. | See Discussion in Manuscript |
|  | 23c | Discuss any limitations of the review processes used. | See Discussion in Manuscript |
|  | 23d | Discuss implications of the results for practice, policy, and future research. | See Discussion in Manuscript |
| **OTHER INFORMATION** | | |  |
| Registration and protocol | 24a | Provide registration information for the review, including register name and registration number, or state that the review was not registered. | NA |
|  | 24b | Indicate where the review protocol can be accessed, or state that a protocol was not prepared. | NA |
|  | 24c | Describe and explain any amendments to information provided at registration or in the protocol. | NA |
| Support | 25 | Describe sources of financial or non-financial support for the review, and the role of the funders or sponsors in the review. | See Funding in Manuscript |
| Competing interests | 26 | Declare any competing interests of review authors. | See Financial Disclosures in Manuscript |
| Availability of data, code and other materials | 27 | Report which of the following are publicly available and where they can be found: template data collection forms; data extracted from included studies; data used for all analyses; analytic code; any other materials used in the review. | NA, all data was accessed after motivated request to the data owners |

*From:*  Page MJ, McKenzie JE, Bossuyt PM, Boutron I, Hoffmann TC, Mulrow CD, et al. The PRISMA 2020 statement: an updated guideline for reporting systematic reviews. BMJ 2021;372:n71. doi: 10.1136/bmj.n71

PRISMA FLOWCHART

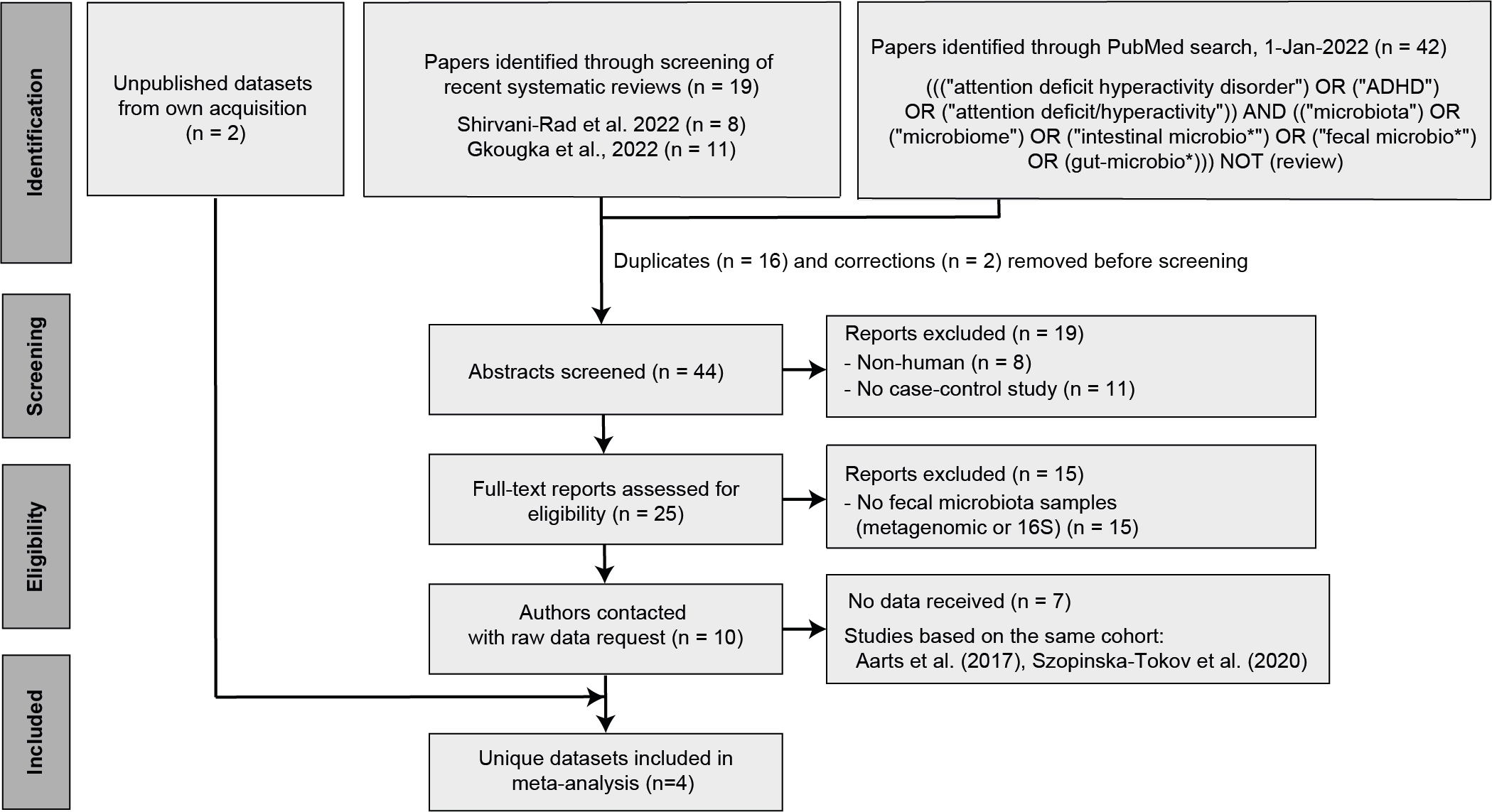

*Supplementary Figure 10. Flow diagram of study selection for the meta-analysis.*
